# A Surgical Planning Pipeline for Human Implantable Brain-Computer Interfaces

**DOI:** 10.64898/2026.02.01.26345325

**Authors:** Peter N. Hadar, Jian Li, Brian Coughlin, William Muñoz, Brian Hsueh, Ziv M. Williams, Seonghwan Yee, Otto Rapalino, David Brandman, Sergey D. Stavisky, Jaimie M. Henderson, Francis R. Willett, Daniel B. Rubin, Leigh R. Hochberg, Sydney S. Cash, Eun Young Choi, Angelique C. Paulk

**Affiliations:** Department of Neurology, Massachusetts General Hospital (MGH), Boston, MA, USA 02114; Athinoula A. Martinos Center for Biomedical Imaging, Department of Radiology, Massachusetts General Hospital, Charlestown, MA, USA 02129; Center for Neurotechnology and Neurorecovery, Department of Neurology, Massachusetts General Hospital, Harvard Medical School, Boston, MA 02114, USA; Department of Neurosurgery, Stanford Medicine, Palo Alto, CA 94304, USA; Department of Neurosurgery, Massachusetts General Hospital (MGH), Boston, MA, USA 02114; Department of Radiology, Massachusetts General Hospital (MGH), Boston, MA, USA 02114; Department of Neurological Surgery, University of California Davis Health, Sacramento, CA 95816, USA; Wu Tsai Neurosciences Institute, Stanford University, Stanford, CA, USA; VA Center for Neurorestoration and Neurotechnology, Office of Research and Development, VA Providence Healthcare System, Providence, RI, USA; Robert J. and Nancy D. Carney Institute for Brain Science, Brown University, Providence, RI, USA; School of Engineering, Brown University, Providence, RI, USA

## Abstract

As implantable brain-computer interfaces (iBCIs) for communication and movement transition from cutting-edge research to clinical practice, a standardized approach will be required to reliably plan neurosurgeries involving complex microelectrode arrays and other neural sensors. Here, through our BrainGate study experiences, we present a replicable methodology, using open-source tools, to create interactive, personalized, 3-dimensional, virtual and physical, functional mapping models to guide iBCI surgical planning and provide intra-operative imaging displays.

## Main Text

Recent advances in brain-computer interfaces (BCIs) have demonstrated the capability to turn intention into action, thereby restoring communication and movement for people with severe functional impairment resulting from neurologic disease or neurotrauma^1–6^. For individuals experiencing the symptoms of neurologic conditions such as stroke, amyotrophic lateral sclerosis (ALS), and spinal cord injury, the potential for meaningful functional recovery with currently available therapies may be limited. However, research into the neural representations of communication and movement has provided potential targets to bridge the gap between intention and action^7,8^. Intracortical BCIs, in which microelectrodes are implanted directly onto the cortical surface, leverage neural signals to decode attempted upper extremity movement to control a computer cursor; interpret attempted handwriting; decipher attempted speech at greater than 60 words a minute; and even restore some conversational communication^9–14^. Ongoing research suggests that this technology has the potential to significantly enhance communication and restore upper extremity movement, enabling greater functional independence and improving quality of life for people with paralysis^1^.

The BrainGate clinical trial (ClinicalTrials.gov ID: NCT00912041) is a multi-center and multi-disciplinary initiative focused on developing intracortical BCIs to restore mobility, communication, and functional independence for people with paralysis caused by neurologic disease or injury. There have been twenty individuals who have enrolled in the BrainGate clinical trial and undergone placement of an intracortical BCI; these participants have made invaluable contributions to our understanding of how the brain encodes movement and communication. The success of these studies is dependent on extensive pre-surgical planning to ensure that BCI sensors are placed in the appropriate brain regions. Targets for sensor placement are generally informed by both the participant’s specific clinical syndrome, their functional goals, and the aims of the research program. Following enrollment, investigators tentatively plan the cortical locations for implantation (e.g., hand motor control areas of precentral gyrus) after acquiring anatomic and functional brain imaging (**Fig. 1**). The participant then undergoes surgical placement of the microelectrode arrays; this procedure involves a craniotomy to expose the cortical surface so that microelectrodes can be implanted into regions of interest. These arrays are connected via wires to percutaneous pedestals that are placed onto the skull and allow the system to connect to external devices that interpret the neural signals (**Fig. 1B,1C, 2**). While this approach has been useful, the ever-expanding array placements, targets, and imaging data expands the number of variables under consideration by the neurosurgeon and clinical research team, especially when the 3-D brain topography, vasculature, neural sensor wiring, and craniotomy size must be taken into account. Taken together, this 3-D planning task extends beyond the typical pre-planning for a functional neurosurgery procedure and some of the commercial packages available for stereotactic surgery. Therefore, we have developed and integrated a pre-surgical planning system and approach that employs open-source imaging processing tools to significantly improve the quality of information available when planning sensor placement, allowing us to standardize iBCI implantation planning, especially in anticipation of scaling up for incorporation into regular clinical practice.

**Figure 1.**
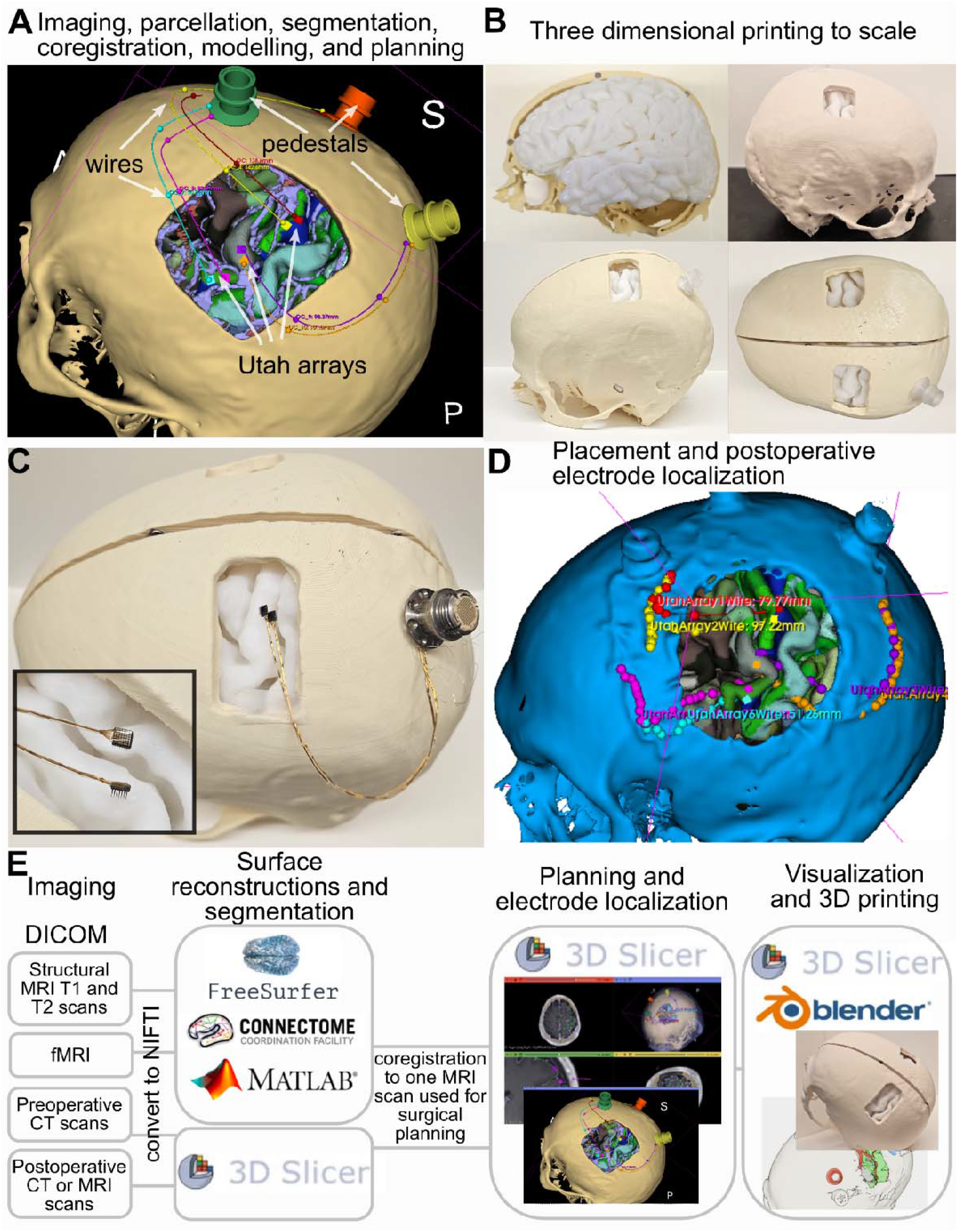
Electrode planning and localization pipeline for Utah array placement. **A)** Preoperative planning after merging imaging, surface generation and segmentation pipelines along with 3D models of the wires, arrays, and pedestals. **B)** Three dimensional printed model of skull, pedestals, and brain surface for planning purposes. **C)** Zoomed in view of Utah arrays alongside the pedestal and wires relative to the 3D printed skull and planned craniotomy. **D)** Postoperative planning after co-registering postoperative scans to the preoperative planning and processed images. **E)** Overall pipeline.

**Figure 2.**
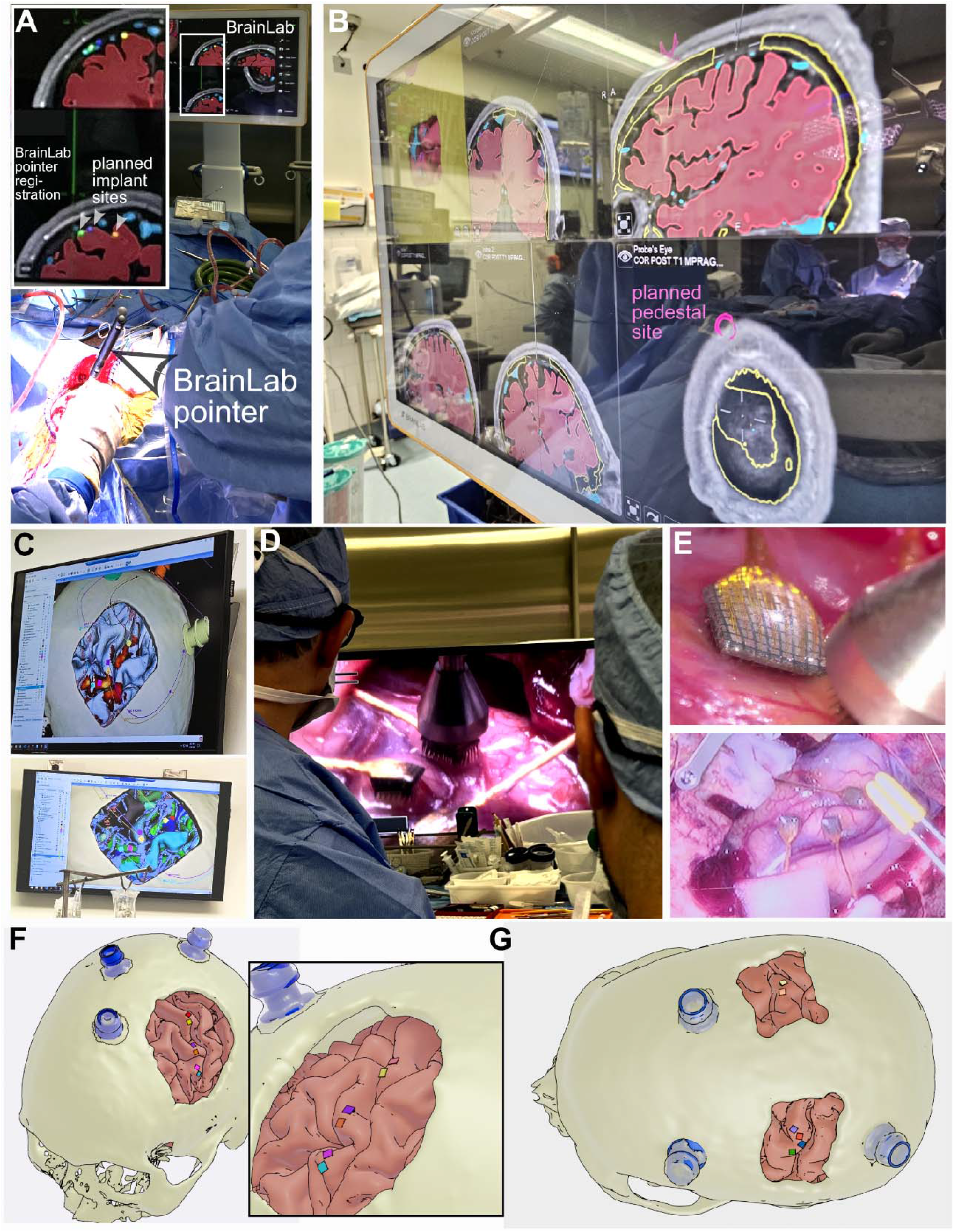
Implanting Utah arrays at the planned locations and craniotomies. **A)** Photograph of the use of the BrainLab pointer used in the surgery and coregistration system of the patient’s MRI and CT coregistered to the brain reconstructions (red) and planned electrode locations (green, blue, and orange dots) as indicated by arrowheads in the inset box outlined in white. **B)** Photograph of the Brainlab screen (with dates and identifying information removed) during the surgery showing the 3D reconstructed brain surface (red) with the planned craniotomy (yellow), vessels (cyan) and planned pedestal locations (pink), with the reflection showing the operation and use of the BrainLab pointer in the brain, skull, and planned craniotomy and array placements. **C)** 3DSlicer view of the planned electrode placements on the monitor on the wall in the operating room during the surgery, with the coloring on the brain reflecting functional MRI activity during a task (top) or the HCP parcellation (bottom). **D)** View in the operating room of the OrbEye camera during placement of the wand inserter and Utah arrays. **E)** Top: close up view of an implanted Utah array; Bottom: view of three placed arrays in the cortex. **F)** 3D representation of the postoperative implants localizations for the first participant of the craniotomy, pedestals, and Utah array placements noted. **G)** 3D representation of the postoperative implants localizations for the second participant depicting the bilateral craniotomies, pedestal, and Utah array placements.

For example, the available implant locations on the cortical surface can be more precisely and confidently identified by examining the 3-D curvature of the brain and skull and device model both in 3-D-rendering software and with physical 3-D printed models (**Fig. 1B,C**). A participant’s cortical vasculature, including small arteries and veins, limits the placement of microelectrode arrays; dedicated vessel imaging can be used to identify these potential obstacles prior to surgery. Finally, the precise overlap and co-registration of anatomical features with functional imaging boundaries of different neural representations of movement can be difficult to visualize in two dimensions and may vary based on a number of non-trivial parameters chosen during the imaging processing pipeline. To address these concerns, we developed a replicable pre-surgical imaging pipeline to assist with functional localization. This pipeline involving standardized imaging acquisition protocols and open-source tools for image processing and 3-D interactive display, the results of which can be displayed intra-operatively on-screen in 3-D (**Fig. 1**,**2**). Further, the output of this software can be printed with the planned craniotomy and 3-D model of the skull and brain for further planning and coordination (**Fig. 1**,**2**).

This methodology was implemented in two participants recently enrolled in the BrainGate clinical trial; one participant, who has a history of advanced ALS, had six microelectrode arrays placed in the speech and hand motor areas in the left hemisphere. The other participant, who has a history of cervical spinal cord injury, underwent placement of microelectrode arrays in the hand motor cortices bilaterally. Following a standardized imaging acquisition protocol (detailed in **Supplementary Material 1 and 2**) involving a head CT and 3T MRI that includes MPRAGE, multi-shell diffusion, post-contrast MRI, vessel imaging, resting state functional MRI (rs-fMRI) with blood oxygen level dependent (BOLD) contrast, and task-based BOLD imaging, the images were preprocessed and directly imported into the free, open-source 3DSlicer software^15,16^, which acted as the central platform for importing and co-registering the different modalities displayed on the participant’s anatomic brain scan.

The structural MR images were processed using the FreeSurfer recon-all pipeline with outputs including the anatomical cortical parcellation and reconstruction of the pial surfaces^17^. Using resting state fMRI (rs-fMRI), we additionally applied the Human Connectome Project (HCP) pipeline, an approach first described in our cohort by Willet et al. (2023), to generate individualized multi-modal cortical parcellation maps based on both structural and functional data^14,18^. The task-based fMRI data were preprocessed using fMRIPrep and the task active regions were localized via a generalized linear modeling (GLM) with respect to the task design using the FreeSurfer Functional Analysis Stream (FsFast)^17,19^. The rs-fMRI data were also preprocessed by fMRIPrep and we identified the spatial maps for the sensorimotor networks, such as hand, foot, and face/tongue area, based on the Nadam-accelerated Scalable and Robust CP Decomposition (NASCAR) network atlases^20–22^. All outputs were projected onto the participant’s native surface space and can be read in MATLAB and Python to produce the pial surfaces and surface maps, which can be co-registered to the other imaging modalities (**Fig. 1E**; https://github.com/Center-For-Neurotechnology/BrainInterface3D).

For the pre-surgical planning conference, the processed structural, functional, and diffusion imaging were imported into the free, open-source 3DSlicer software^15,16^. The head CT was imported to provide the skull model using segmentation, and the post-contrast MRI and MR venogram (MRV) were also imported to incorporate vascular anatomy. These were all co-registered to a specific anatomic T1-weighted scan selected by the neurosurgeon for intraoperative co-registration using ANTs registration^23^. 3D models of the microelectrode arrays and pedestals were imported to 3Dslicer; to assist the clinical team in the surgical planning, 3D-printed models of the brain, skull, arrays, and pedestals were also generated, which allowed for easier in-person discussions on surgical approaches (**Fig. 1A-D**).

During the pre-surgical planning conference, the 3DSlicer results were used to review the anatomic and functional boundaries of the neural representations of motor and language function. The locations of arteries and veins were noted, and locations for each of the planned the microelectrode arrays were proposed. The investigators also used the 3D-printed brain and skull models to help guide decision making. Using the 3DSlicer software, these array models were interactively moved into the intended locations. Following satisfactory placement of the arrays in the 3D model, the boundaries of the craniotomy on the skull were determined following recommended clinical trial limits and the percutaneous pedestals were placed on the model. The wire bundles connecting the microelectrode arrays to the pedestals were also modeled and putative wire trajectories were mapped. By the end of the interactive conference, a 3D model of the participant’s brain and skull, incorporating anatomic, functional, and vascular imaging modalities, was created that included the intended locations of each of the microelectrode arrays, wire bundles, pedestals, and craniotomies.

Following the pre-surgical planning conference, the final models and outputs from the interactive 3DSlicer session were saved into images (MRI) or segmentations (with coded information indicating values for per-slice representations of 3D models) as nifti imaging format and converted to the DICOM imaging format through Karawun^24^. This conversion was necessary to upload the imaging to our image guidance platforms, which at our institution (MGH) included pushing it to the Visage (Visage Version 7.1, Visage Imaging GmbH, Berlin, Germany) picture archiving and communication system (PACS) and then pushing it to BrainLab, a data integration and navigation system used in the operating room^25^. During the operative procedure, these models were displayed on the screen and could be interactively rotated or magnified per the neurosurgeon’s request (**Fig. 2B-D**). Following the implants, post-operative CT imaging of the electrode locations paired with photographs of the implanted electrodes into the cortex were used to confirm the electrode locations. Given the success with these two participants, especially in our ability to minimize the craniotomy size, inform pre-operative surgical planning and intra-operative procedure, and ensure that the pedestal placements are optimized relative to the craniotomies and other patient-specific features (e.g., hairline, wheelchair headrest), elements of this approach are used throughout the BrainGate clinical trial.

## Methods

### Imaging Acquisitions

Both participant MRI scans were conducted on a Siemens Magnetom 3.0T XR Numaris scanner. The complete protocol for participant MRI scans can be found in **Supplementary Material 2**. The protocol was adapted from the HCP protocol with 2 runs of anterior-posterior/posterior-anterior field maps and fMRI scans (resting-state and task-based). For task-based MRI scans, tasks involving opening-closing either hand, reaching-grasping, toe-tapping, lip movement, and language were used to help identify motor and language regions.

Sequence parameters for the Sagittal T1 3D MPRAGE were resolution 1.0x1.0x0.9mm^3^, repetition time (TR) of 2300 ms, echo time (TE) 2.32 ms, flip angle of 8 degrees, parallel imaging using GRAPPA with an acceleration factor of 2, and time of acquisition (TA) of 5:21 minutes. Diffusion sequence parameters involved 3 runs of anterior-posterior encoding and 3 runs of posterior-anterior encoding with 95, 96, and 97 directions, respectively. Post-contrast T1 and T2 scans were completed, and a sagittal 3D phase contrast MR venography was also performed.

### Image Processing

Three image processing pipelines were adopted to provide complementary information (see **Supplementary Figure 1** for the consolidated planning and post-operative image processing workflows). A common analysis step across the pipelines was running FreeSurfer (v7.4 for two of the pipelines and v6.0 for one of the pipelines) recon-all (cite FS) on the preoperative structural images including the T1-weighted scan and the T2-weighted scan when available. The outcomes from this FreeSurfer processing include reconstruction and modeling of the pial and white matter surfaces and anatomical cortical parcellations which serve as the bases for the following analysis.

#### Cortical parcellation

As part of the FreeSurfer outputs, we obtained the anatomical cortical parcellation map based on the DKT atlas^26,27^. We additionally applied the Human Connectome Project (HCP) pipeline (https://github.com/Washington-University/HCPpipelines) to generate individualized cortical parcellation^18,28^ (Glasser et al. 2016). This HCP-based pipeline was implemented on a Linux system with the FMRIB Software Library (FSL) version 6.0, FreeSurfer 6.0, and Connectome Workbench 1.4.2 (https://github.com/Washington-University/HCPpipelines). The minimal preprocessing pipeline (Glasser 2013) was used to preprocess the preoperative T1 and T2 scans, the rs-fMRI scans (as they were of high enough quality) with ICA-fix denoising and MSMAII registration for one participant. For the second participant, we only performed the minimal preprocessing pipeline but without the rs-fMRI processing steps. We took an additional step which was to map the parcellations mapping gathered in a larger cohort of individuals (Glasser et al., 2013, 2016) to the HCP and FreeSurfer 7.4 output (separate outputs) in two different approaches. First, we used an HCP annotation mapped to the FreeSurfer space and a series of FreeSurfer commands and an annotation map projected onto FreeSurfer space (https://figshare.com/articles/dataset/HCP-MMP1_0_projected_on_fsaverage/3498446?file=5528837) to convert the labels into native space which could then be viewed in 3D in and 3DSlicer Blender (https://github.com/Center-For-Neurotechnology/BrainInterface3D) ^29–32^. The second parcellation approach involved applying the Minimal Preprocessing Pipeline (MPP) to the preoperative MRI T1 and T2 scans (Glasser et al. 2013) followed by Connectome Workbench commands to map the HCP parcellation back to native space (https://github.com/Center-For-Neurotechnology/BrainInterface3D). These results were saved as in the CIFTI and GIFTI imaging format and then converted to .ply files (for surfaces) using the Gifti library for MATLAB (https://github.com/gllmflndn/gifti). The postoperative CT was co-registered to the preoperative T1 scans used in the cortical reconstructions using the SlicerANTS extension in 3DSlicer (https://github.com/simonoxen/SlicerANTs).

#### Task localizers

The t-fMRI data were preprocessed using fMRIPrep, including slicing time correction, motion correction, susceptibility distortion correction, co-registration to T1w image^19^. The BOLD signals were sampled onto the FreeSurfer surfaces and converted to the grayordinate space, saved in the CITFI/GIFTI compatible format and converted to .ply files (for surfaces) using the Gifti library for MATLAB (https://github.com/gllmflndn/gifti). The preprocessed t-fMRI data were then fed into the FreeSurfer Functional Analysis Stream (FsFast) for surface-based smoothing with a 4 mm full-width-half-maximum kernel, and a generalized linear modeling (GLM) with respect to the task design^17^. The GLM result were represented as a significance map (-log(p-value)) for each task indicating the key region(s) that was involved in that task.

#### Resting-state network mapping

The rs-fMRI data were also preprocessed by fMRIPrep as the task data. However, in contrast to the task data, we did not apply any additional smoothing to best preserve clear boundaries across different functional regions^33^. To identify spatially overlapped and temporally correlated large-scale brain networks from rs-fMRI data, we used the NASCAR network atlases as our bases and estimated the individual spatial map for each network by regressing the rs-fMRI data against the temporal mode of the NASCAR atlases^20–22,34^. The spatial map for sensorimotor networks, such as hand, foot, and face/tongue area, were successfully identified.

For the 3-D models which could be viewed in 3Dslicer as well as 3-D printed, the FreeSurfer 7.4 pial surface was imported into MATLAB using the freesurferReadSurf function along with the annotations identifying the parcellations. The HCP parcellated surfaces and output were imported using the GIFTI toolbox (https://github.com/gllmflndn/gifti) which read the labels as well as the surfaces. The brain surfaces created with Freesurfer 7.4 and HCP were finally converted into .ply files using MATLAB with the .ply vertices color-coded by parcellation value (https://github.com/Center-For-Neurotechnology/BrainInterface3D) using the surfaceMesh and writeSurfaceMesh functions. This conversion also allows for color-shifting the targeted regions. Any fMRI surface imaging highlighting function (e.g. BOLD signal) was also exported to a color-coded .ply surface using the MATLAB surfaceMesh and writeSurfaceMesh functions (https://github.com/Center-For-Neurotechnology/BrainInterface3D). The head CT was imported to provide the skull model through the Segmentation tool, creating the 3-D skull by thresholding the bright image values. The segmented volume as then converted into a model in 3Dslicer and then exported as an .obj file to be printed in 3D. The post-contrast T1 MPRAGE and MRV head scans were used to help delineate the vascular anatomy. The vasculature was then also segmented to create 3D volumes of vessels. To increase the contrast of the vessels, the pre-contrast T1 MPRAGE was subtracted from the post-contrast T1 MPRAGE through volume subtraction in 3Dslicer and then the vessels were segmented. Further manual Segmentation editing was performed to remove vasculature from the skin and dura above the surface of the brain.

### Pre-Operative Planning and Display

The results of the imaging preprocessing pipelines (Freesurfer 7.4 and HCP), brain and skull models, original images as NIFTI files, and 3D models of the microelectrode arrays and pedestals were imported into 3DSlicer to create a consolidated 3D brain and skull model for pre-surgical planning (MNI model with components are provided in **Supplementary Material 3**). 3DSlicer is interactive, and during the pre-surgical planning conference, investigators could request to view separate modalities, such as functional imaging or vascular imaging, to allow for a better delineation of the target region. Based on the pre-surgical planning conference, the location of the microelectrode array placements, pedestals, and wire were inputted and saved in the 3D model. Using the Segmentation Editor tool, the skull model was edited to represent the extent of the craniotomy and was saved as a model. The 3-D skull with the craniotomy was then exported as an .obj file to be then printed in 3-D which allowed enough placement of the 3-D brain hemispheres to be inserted and viewed together in person. The 3D MNI model files are provided in **Supplementary Material 3** to create a similar model.

### Intra-Operative Display

Once the final pre-surgical decisions were made, the volume files (MRI or CT imaging, segmentation volumes of the 3-D models, etc.) were converted from the nifti imaging format to the DICOM imaging format with the correct headers for use in the clinical system through Karawun. This conversion was necessary to allow for the modified files to be displayed in the operating room within the clinical space. The DICOM files were then uploaded to the Visage PACS and were pushed to the BrainLab machine used in the operating room. These 3D models could then be displayed on a screen in the operating room and, in an interactive fashion, could be rotated or magnified during the case per the neurosurgeon’s request. Further, the 3DSlicer view could be moved to display the same view as the open craniotomy as pictured using the overhead OR camera or the OrbEye^35^. A key aspect of this work was that the participant’s head and face were co-registered to the BrainLab display such that the planned placement and craniotomies could be directly mapped to the skull and head in real-time.

## Supporting information

Supplementary Data Part 1

Supplementary Data Part 2

Supplementary Data Part 3

Supplementary Data Part 4

Supplementary Material: BrainGate Imaging Protocol

## Data Availability

The code for reproducing the figures is made available at https://github.com/Center-For-Neurotechnology/BrainInterface3D. The data required to reproduce the visualizations in this study are publicly available on FigShare. The dataset contains neuroimaging data recorded from participants, including 3D models of the brain and skull.

## Acknowledgements and Funding

The investigators would like to thank the participants, their families, and care partners for their extraordinary dedication to this research. The investigators would also like to thank the collaborators on the scientific research teams at Massachusetts General Hospital, Brown University, Providence VA Medical Center, Stanford, UC Davis, and Emory.

## CAUTION

Investigational Device. Limited by Federal Law to Investigational Use. The content is solely the responsibility of the authors and does not necessarily represent the official views of the National Institutes of Health, or the Department of Veterans Affairs, or the United States Government.

The funders had no role in the study design, data collection and interpretation, or the decision to submit the work for publication. The work described in this manuscript was supported by: NIH-National Institute on Deafness and Other Communication Disorders (NIDCD) from U01DC017844 (LRH), R01DC014034 (JMH), and NIH-National Institute of Neurologic Disorders and Stroke (NINDS) from UH2NS095548 (LRH) and U01NS098968 (SSC) and R01NS134410 (ACP), Searle Scholars (SDS), Burroughs Wellcome Fund Career Award at the Scientific Interface (SDS), CDMRP ALS Pilot Clinical Trial Award (AL220043) from the Office of the Assistant Secretary of Defense for Health Affairs (SDS), the Wu Tsai Neurosciences Institute at Stanford (JMH), and Larry and Pamela Garlick (JMH).

---

**Supplementary Figure 1:**
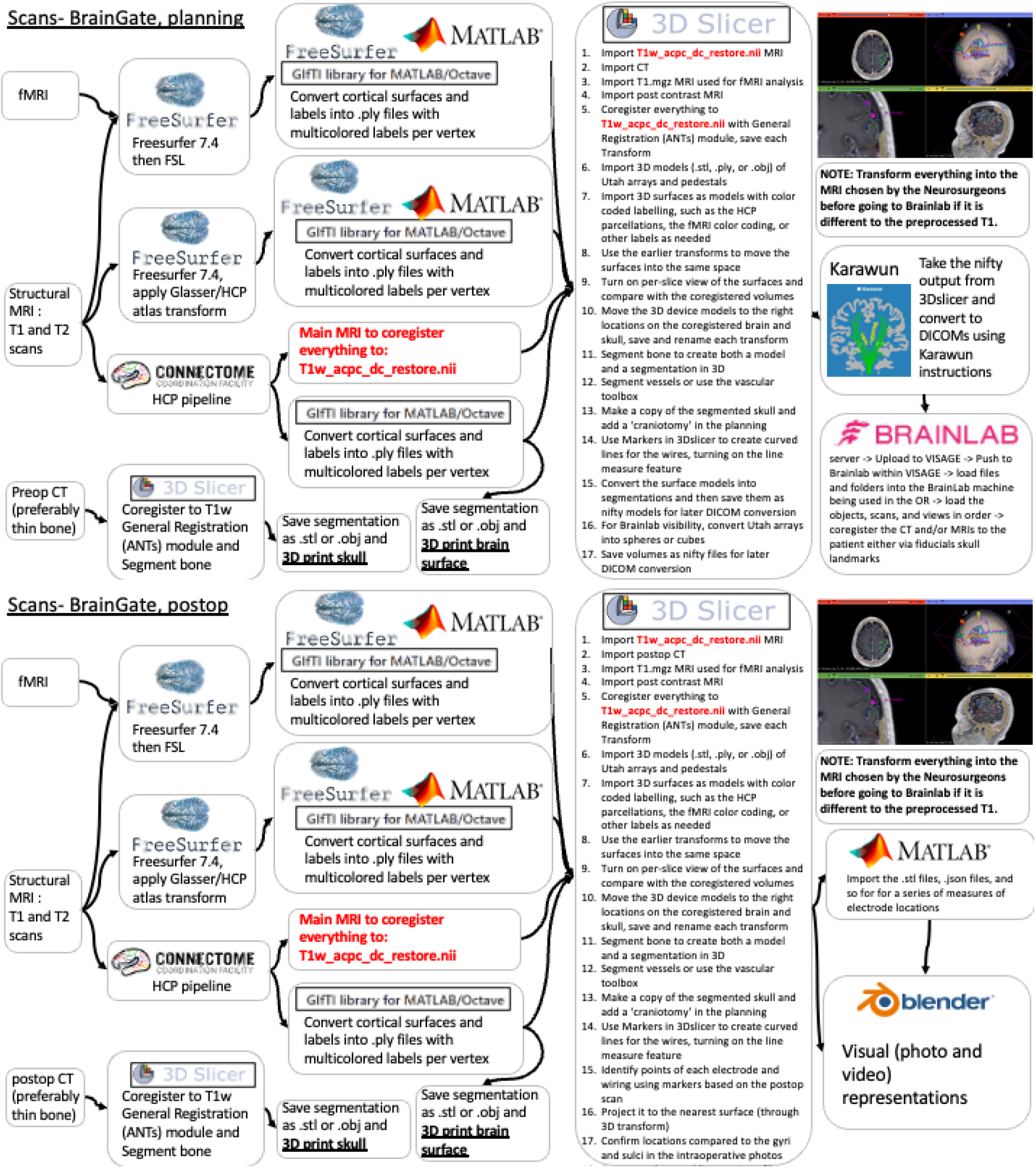
Surgical Planning Imaging Pipeline Overview. A) A schematic of the image processing workflow is shown for the pre-operative phase. Structural MRI scans (T1- and T2-weighted scans), functional MRI scans, and a head CT are used for reconstruction and to model the pial and white matter surfaces and anatomical cortical parcellations; functional network mapping and vascular segmentation was also performed. 3D Slicer was used for visualization, and the results of these overlays were converted back into DICOM format through Karawun and uploaded to BrainLab for intra-operative use. B) A schematic of the image processing workflow is shown for the post-operative phase. There is an initial common analysis pipeline that now incorporates a post-operative head CT scan, and 3D Slicer is then used to help identify the extent of the craniotomy and electrode placements for visualization; Blender is also used to help with the visual representations.

