## Supplementary figures and images for "A Surgical Planning Pipeline for Human Implantable Brain-Computer Interfaces"

### MNIDemo-Scene.png

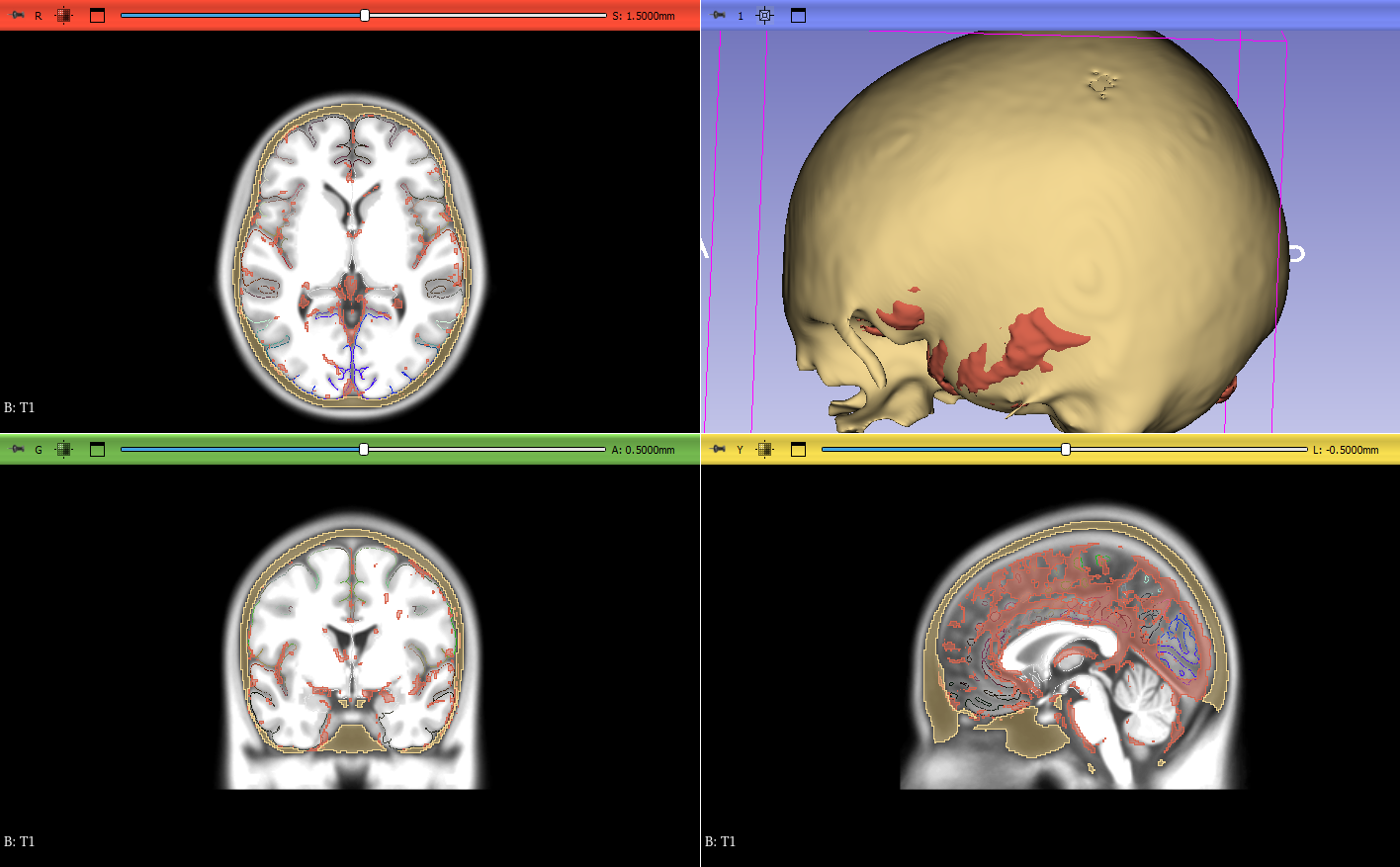
