## Supplementary Material: BrainGate Imaging Protocol for "A Surgical Planning Pipeline for Human Implantable Brain-Computer Interfaces"

### Table of contents

\\PHYSICS

XA30 RESEARCH PROT

PARCELLATION

[redacted]

Localizer
AAHScout
Localizer_aligned
Sag_T1_3D_Mprage
Sag_T2_3D_Space_Wave
SpinEchoFieldMap_1_AP
SpinEchoFieldMap_1_PA
rfMRI_REST_1_AP
rfMRI_REST_1_PA
SpinEchoFieldMap_2_AP
SpinEchoFieldMap_2_PA
rfMRI_REST_2_AP
rfMRI_REST_2_PA
OPEN-CLOSE RT HAND_FMRI
OPEN-CLOSE LT HAND_FMRI
LIPS_FMRI
TOE TAPPING_ RT FOOT FMRI
REACH-GRASP RT HAND_FMRI
REACH-GRASP LT HAND_FMRI
Ax_Flair
Sag_Flair_3D_Space_Wave
Ax_DWI
Sag 3D PC MRV Head
Ax_SWI_Wave
dMRI_dir95_AP
dMRI_dir95_PA
dMRI_dir96_AP
dMRI_dir96_PA
dMRI_dir97_AP
dMRI_dir97_PA
Post_Sag_T2_3D_Space_Wave
Post Sag_T1_3D_Mprage

**\\PHYSICS\XA30 RESEARCH PROT\PARCELLATION\BRAINGATE2 08-26-2024\Localizer**

TA: 9 sec Coil Selection: Auto Voxel Size: 1.2×1.2×5.0 mm³ Acc:: None Rel. SNR: 1.00

### Properties

| | |
|---|---|
| Start measurement without further preparation | On |
| Wait for User to Start | On |
| Start measurements | Single Measurement |
| Prio Recon | On |
| Auto Open Inline Display | Off |
| Auto Close Inline Display | Off |
| Load Images to MR View&GO | Off |
| Auto Store Images | On |
| Load Images to Stamp Segments | On |
| Load Images to Graphic Segments | On |
| Graphic segment | All Segments |
| Inline Movie | Off |

### Routine

| | |
|---|---|
| Slice Group | 1 |
| Slices | 1 |
| Distance Factor | 20 % |
| Position | L0.0 A20.0 H0.0 mm |
| Orientation | Transversal |
| Phase Encoding Dir. | A >> P |
| Slice Group | 2 |
| Slices | 1 |
| Distance Factor | 20 % |
| Position | L0.0 A20.0 H0.0 mm |
| Orientation | Transversal |
| Phase Encoding Dir. | A >> P |
| Slice Group | 3 |
| Slices | 1 |
| Distance Factor | 20 % |
| Position | L0.0 A20.0 H0.0 mm |
| Orientation | Transversal |
| Phase Encoding Dir. | A >> P |
| Phase Oversampling | 0 % |
| FoV Read | 300 mm |
| FoV Phase | 100.0 % |
| Slice Thickness | 5.0 mm |
| TR | 40.0 ms |
| TE | 3.00 ms |
| Averages | 1 |
| Concatenations | 1 |
| AutoAlign | --- |

### Contrast - Common

| | |
|---|---|
| TR | 40.0 ms |
| TE | 3.00 ms |
| MTC | Off |
| Magn. Preparation | None |
| Flip Angle | 15 deg |
| Fat-Water Contrast | Standard |
| Dark Blood | Off |
| Contrasts | 1 |
| SWI | Off |
| Reconstruction | Magnitude |

### Contrast - Dynamic

| | |
|---|---|
| Dynamic Mode | Standard |
| Measurements | 1 |
| Multiple Series | Off |

### Resolution - Common

| | |
|---|---|
| FoV Read | 300 mm |
| FoV Phase | 100.0 % |
| Slice Thickness | 5.0 mm |
| Base Resolution | 256 |
| Phase Resolution | 75 % |
| Interpolation | Off |

### Resolution - Acceleration

| | |
|---|---|
| Acceleration mode | None |
| Advanced Reconstruction | Off |
| Phase Partial Fourier | Off |
| Asymmetric Echo | Allowed |

### Resolution - Filter

| | |
|---|---|
| Raw Filter | Off |
| Elliptical Filter | On |
| Distortion Correction | 2D |
| Normalize | Prescan |
| Image Filter | Off |

### Geometry - Common

| | |
|---|---|
| Slice Group | 1 |
| Slices | 1 |
| Distance Factor | 20 % |
| Position | L0.0 A20.0 H0.0 mm |
| Orientation | Transversal |
| Phase Encoding Dir. | A >> P |
| Slice Group | 2 |
| Slices | 1 |
| Distance Factor | 20 % |
| Position | L0.0 A20.0 H0.0 mm |
| Orientation | Transversal |
| Phase Encoding Dir. | A >> P |
| Slice Group | 3 |
| Slices | 1 |
| Distance Factor | 20 % |
| Position | L0.0 A20.0 H0.0 mm |
| Orientation | Transversal |
| Phase Encoding Dir. | A >> P |
| Phase Oversampling | 0 % |
| FoV Read | 300 mm |
| FoV Phase | 100.0 % |
| Slice Thickness | 5.0 mm |
| TR | 40.0 ms |
| Multi-Slice Mode | Interleaved |
| Series | Interleaved |
| Concatenations | 1 |

### Geometry - AutoAlign

| | |
|---|---|
| Slice Group | 1 |
| Position | L0.0 A20.0 H0.0 mm |
| Orientation | Transversal |
| Phase Encoding Dir. | A >> P |
| Slice Group | 2 |
| Position | L0.0 A20.0 H0.0 mm |
| Orientation | Transversal |
| Phase Encoding Dir. | A >> P |
| Slice Group | 3 |
| Position | L0.0 A20.0 H0.0 mm |
| Orientation | Transversal |

### Geometry - AutoAlign

| | |
|---|---|
| Phase Encoding Dir. | A >> P |
| AutoAlign | --- |
| Initial Position | L0.0 A20.0 H0.0 |
| L | 0.0 mm |
| A | 20.0 mm |
| H | 0.0 mm |
| Initial Orientation | Transversal |
| Initial Rotation | 0.00 deg |

### Geometry - Saturation

| | |
|---|---|
| Saturation Mode | Standard |
| Special Saturation | None |

### Geometry - Tim Planning Suite

| | |
|---|---|
| Set-n-Go Protocol | Off |
| Table Position | 0 mm |
| Table Position | H |
| Inline Composing | Off |

### System - Miscellaneous

| | |
|---|---|
| Coil Selection | ACS All but spine |
| MSMA | S - C - T |
| Sagittal | R >> L |
| Coronal | A >> P |
| Transversal | F >> H |
| Coil Combination | Sum of Squares |
| Matrix Optimization | Off |
| Coil Focus | Flat |

### System - Adjustments

| | |
|---|---|
| Adjustment Strategy | Standard |
| B0 Shim | Tune up |
| B1 Shim | TrueForm |
| CoilShim | Off |
| Adjustment Tolerance | Auto |
| Adjust with Body Coil | Off |
| Confirm Frequency | Never |
| Assume Silicone | Off |

### System - Adjust Volume

| | |
|---|---|
| Position | Isocenter |
| Orientation | Transversal |
| Rotation | 0.00 deg |
| A >> P | 263 mm |
| R >> L | 350 mm |
| F >> H | 350 mm |
| Reset | Off |

### System - pTx

| | |
|---|---|
| B1 Shim | TrueForm |
| Excitation | Slice-sel. |
| LR Balancing | Off |

### System - Tx/Rx

| | |
|---|---|
| Frequency 1H | 123.257916 MHz |
| ? Ref. Amplitude 1H | 0.000 V |
| Reset | Off |
| Image Scaling | 1.000 |

### Physio - Signal

| | |
|---|---|
| 1st Signal/Mode | None |
| TR | 40.0 ms |
| Segments | 1 |

### Physio - Signal

| | |
|---|---|
| Concatenations | 1 |

### Physio - Cardiac

| | |
|---|---|
| Tagging | None |
| Fat-Water Contrast | Standard |
| Magn. Preparation | None |
| Dark Blood | Off |
| FoV Read | 300 mm |
| FoV Phase | 100.0 % |
| Phase Resolution | 75 % |
| Dynamic Mode | Standard |

### Physio - PACE

| | |
|---|---|
| Resp. Control | Off |
| Concatenations | 1 |

### Inline - Liver

| | |
|---|---|
| Liver Registration | Off |
| Save Original Images | On |

### Inline - Subtraction

| | |
|---|---|
| Subtract | Off |
| Measurements | 1 |
| StdDev | Off |
| Save Original Images | On |

### Inline - Cardiac

| | |
|---|---|
| Magn. Preparation | None |
| Save Original Images | On |
| Contrasts | 1 |
| TE | 3.00 ms |
| TR | 40.0 ms |

### Inline - MIP

| | |
|---|---|
| MIP Sag | Off |
| MIP Cor | Off |
| MIP Tra | Off |
| MIP Time | Off |
| Radial MIP | Off |
| Save Original Images | On |
| MPR Sag | Off |
| MPR Cor | Off |
| MPR Tra | Off |

### Inline - Soft Tissue

| | |
|---|---|
| Wash-in | Off |
| Wash-out | Off |
| TTP | Off |
| PEI | Off |
| MIP Time | Off |
| Measurements | 1 |

### Inline - Composing

| | |
|---|---|
| Inline Composing | Off |

### Inline - MapIt

| | |
|---|---|
| MapIt | None |
| Flip Angle | 15 deg |
| Measurements | 1 |
| Contrasts | 1 |
| TE | 3.00 ms |
| TR | 40.0 ms |
| Save Original Images | On |

### Sequence - Part 1

| | |
|---|---|
| Sequence Name | fl |
| Dimension | 2D |
| Excitation | Slice-sel. |
| RF Pulse Type | Normal |
| Gradient Mode | Fast |
| Flow Compensation | None |
| Bandwidth | 260 Hz/Px |
| Asymmetric Echo | Allowed |
| Segments | 1 |

### Sequence - Part 2

| | |
|---|---|
| Introduction | On |
| RF Spoiling | On |
| Acoustic noise reduction | Off |

### Sequence - Assistant

| | |
|---|---|
| SAR Assistant | Off |
| Allowed Delay | 0 s |

**\\PHYSICS\XA30 RESEARCH PROT\PARCELLATION\BRAINGATE2 08-26-2024\AAHScout**

TA: 14 sec Coil Selection: Auto Voxel Size: 1.6×1.6×1.6 mm³ Acc:: 3 Rel. SNR: 1.00

### Properties

| | |
|---|---|
| Start measurement without further preparation | Off |
| Wait for User to Start | Off |
| Start measurements | Single Measurement |
| Prio Recon | On |
| Auto Open Inline Display | Off |
| Auto Close Inline Display | Off |
| Load Images to MR View&GO | On |
| Auto Store Images | On |
| Load Images to Stamp Segments | Off |
| Load Images to Graphic Segments | Off |
| Graphic segment | Default |
| Inline Movie | Off |

### Routine

| | |
|---|---|
| Slab Group | 1 |
| Slabs | 1 |
| Distance Factor | 20 % |
| Position | L0.0 A20.0 H0.0 mm |
| Orientation | Sagittal |
| Phase Encoding Dir. | A >> P |
| Slices per Slab | 128 |
| Phase Oversampling | 0 % |
| Slice Oversampling | 0.0 % |
| FoV Read | 260 mm |
| FoV Phase | 100.0 % |
| Slice Thickness | 1.6 mm |
| TR | 3.2 ms |
| TE | 1.37 ms |
| Averages | 1 |
| Concatenations | 1 |
| AutoAlign | Head |

### Contrast - Common

| | |
|---|---|
| TR | 3.2 ms |
| TE | 1.37 ms |
| Flip Angle | 8 deg |
| Fat-Water Contrast | Standard |
| Contrasts | 1 |
| Reconstruction | Magnitude |

### Contrast - Dynamic

| | |
|---|---|
| Dynamic Mode | Standard |
| Measurements | 1 |
| Time to Center | 6.2 s |

### Resolution - Common

| | |
|---|---|
| FoV Read | 260 mm |
| FoV Phase | 100.0 % |
| Slice Thickness | 1.6 mm |
| Base Resolution | 160 |
| Phase Resolution | 100 % |
| Slice Resolution | 69 % |
| Trajectory | Cartesian |

### Resolution - Acceleration

| | |
|---|---|
| Acceleration mode | GRAPPA |
| Reference Scans | Integrated |
| Acceleration Factor PE | 3 |
| Reference Lines PE | 24 |

### Resolution - Acceleration

| | |
|---|---|
| Acceleration Factor 3D | 1 |
| Phase Partial Fourier | 6/8 |
| Slice Partial Fourier | 6/8 |
| Asymmetric Echo | Weak |

### Resolution - Filter

| | |
|---|---|
| Raw Filter | Off |
| Elliptical Filter | Off |
| Distortion Correction | 3D |
| Normalize | Prescan |
| Noise Masking | Off |
| Image Filter | Off |

### Geometry - Common

| | |
|---|---|
| Slab Group | 1 |
| Slabs | 1 |
| Distance Factor | 20 % |
| Position | L0.0 A20.0 H0.0 mm |
| Orientation | Sagittal |
| Phase Encoding Dir. | A >> P |
| Slices per Slab | 128 |
| Phase Oversampling | 0 % |
| Slice Oversampling | 0.0 % |
| FoV Read | 260 mm |
| FoV Phase | 100.0 % |
| Slice Thickness | 1.6 mm |
| TR | 3.2 ms |
| Multi-Slice Mode | Sequential |
| Series | Ascending |
| Concatenations | 1 |

### Geometry - AutoAlign

| | |
|---|---|
| Slab Group | 1 |
| Position | L0.0 A20.0 H0.0 mm |
| Orientation | Sagittal |
| Phase Encoding Dir. | A >> P |
| AutoAlign | Head |
| Initial Position | Isocenter |
| L | 0.0 mm |
| P | 0.0 mm |
| H | 0.0 mm |
| Initial Orientation | Transversal |
| Initial Rotation | 0.00 deg |

### Geometry - Tim Planning Suite

| | |
|---|---|
| Set-n-Go Protocol | Off |
| Table Position | 0 mm |
| Table Position | H |
| Inline Composing | Off |

### System - Miscellaneous

| | |
|---|---|
| Coil Selection | ACS All but spine |
| MSMA | S - C - T |
| Sagittal | R >> L |
| Coronal | A >> P |
| Transversal | F >> H |
| Coil Combination | Adaptive Combine |
| Matrix Optimization | Off |
| Coil Focus | Flat |

### System - Adjustments

| | |
|---|---|
| Adjustment Strategy | Standard |
| B0 Shim | Tune up |
| B1 Shim | TrueForm |
| CoilShim | Off |
| Adjustment Tolerance | Auto |
| Adjust with Body Coil | Off |
| Confirm Frequency | Never |
| Assume Silicone | Off |

### System - Adjust Volume

| | |
|---|---|
| Position | Isocenter |
| Orientation | Transversal |
| Rotation | 0.00 deg |
| A >> P | 263 mm |
| R >> L | 350 mm |
| F >> H | 350 mm |
| Reset | Off |

### System - pTx

| | |
|---|---|
| B1 Shim | TrueForm |
| Excitation | Non-sel. |

### System - Tx/Rx

| | |
|---|---|
| Frequency 1H | 123.257916 MHz |
| ? Ref. Amplitude 1H | 0.000 V |
| Reset | Off |
| Image Scaling | 1.000 |

### Physio - PACE

| | |
|---|---|
| Resp. Control | Off |
| Concatenations | 1 |

### Inline - Dynamic

| | |
|---|---|
| Dynamic Mode | Standard |
| Flip Angle | 8 deg |
| Measurements | 1 |
| Time to Center | 6.2 s |

### Inline - Subtraction

| | |
|---|---|
| Subtract | Off |
| Measurements | 1 |
| StdDev | Off |
| Save Original Images | On |

### Inline - Cardiac

| | |
|---|---|
| Save Original Images | On |
| Contrasts | 1 |
| TE | 1.37 ms |
| TR | 3.2 ms |

### Inline - MIP

| | |
|---|---|
| MIP Sag | Off |
| MIP Cor | Off |
| MIP Tra | Off |
| MIP Time | Off |
| Radial MIP | Off |
| Save Original Images | On |
| MPR Sag | Off |
| MPR Cor | Off |
| MPR Tra | Off |

### Inline - Composing

| | |
|---|---|
| Inline Composing | Off |

### Inline - MapIt

| | |
|---|---|
| MapIt | None |
| Flip Angle | 8 deg |
| Measurements | 1 |
| Contrasts | 1 |
| TE | 1.37 ms |
| TR | 3.2 ms |
| Save Original Images | On |

### Sequence - Part 1

| | |
|---|---|
| Sequence Name | fl |
| Dimension | 3D |
| Excitation | Non-sel. |
| RF Pulse Type | Fast |
| Gradient Mode | Normal |
| Bandwidth | 540 Hz/Px |
| Asymmetric Echo | Weak |

### Sequence - Part 2

| | |
|---|---|
| Introduction | On |
| RF Spoiling | On |
| Breast Application | Off |

### Sequence - Assistant

| | |
|---|---|
| SAR Assistant | Off |

**\\PHYSICS\XA30 RESEARCH PROT\PARCELLATION\BRAINGATE2 08-26-2024\Localizer_aligned**

TA: 21 sec Coil Selection: Auto Voxel Size: 1.2×1.2×5.0 mm³ Acc:: None Rel. SNR: 1.00

### Properties

| | |
|---|---|
| Start measurement without further preparation | Off |
| Wait for User to Start | Off |
| Start measurements | Single Measurement |
| Prio Recon | On |
| Auto Open Inline Display | Off |
| Auto Close Inline Display | Off |
| Load Images to MR View&GO | On |
| Auto Store Images | On |
| Load Images to Stamp Segments | On |
| Load Images to Graphic Segments | On |
| Graphic segment | All Segments |
| Inline Movie | Off |

### Routine

| | |
|---|---|
| Slice Group | 1 |
| Slices | 1 |
| Distance Factor | 20 % |
| Position | R3.0 A40.6 H27.5 mm |
| Orientation | T > C-4.2 > S3.7 |
| Phase Encoding Dir. | A >> P |
| Slice Group | 2 |
| Slices | 1 |
| Distance Factor | 20 % |
| Position | R3.0 A40.6 H27.5 mm |
| Orientation | T > C-4.2 > S3.7 |
| Phase Encoding Dir. | A >> P |
| Slice Group | 3 |
| Slices | 1 |
| Distance Factor | 20 % |
| Position | R3.0 A40.6 H27.5 mm |
| Orientation | T > C-4.2 > S3.7 |
| Phase Encoding Dir. | A >> P |
| Phase Oversampling | 0 % |
| FoV Read | 300 mm |
| FoV Phase | 100.0 % |
| Slice Thickness | 5.0 mm |
| TR | 104.0 ms |
| TE | 3.00 ms |
| Averages | 1 |
| Concatenations | 1 |
| AutoAlign | Head > Brain |

### Contrast - Common

| | |
|---|---|
| TR | 104.0 ms |
| TE | 3.00 ms |
| MTC | Off |
| Magn. Preparation | None |
| Flip Angle | 15 deg |
| Fat-Water Contrast | Standard |
| Dark Blood | Off |
| Contrasts | 1 |
| SWI | Off |
| Reconstruction | Magnitude |

### Contrast - Dynamic

| | |
|---|---|
| Dynamic Mode | Standard |
| Measurements | 1 |
| Multiple Series | Off |

### Resolution - Common

| | |
|---|---|
| FoV Read | 300 mm |
| FoV Phase | 100.0 % |
| Slice Thickness | 5.0 mm |
| Base Resolution | 256 |
| Phase Resolution | 75 % |
| Interpolation | Off |

### Resolution - Acceleration

| | |
|---|---|
| Acceleration mode | None |
| Advanced Reconstruction | Off |
| Phase Partial Fourier | Off |
| Asymmetric Echo | Allowed |

### Resolution - Filter

| | |
|---|---|
| Raw Filter | Off |
| Elliptical Filter | On |
| Distortion Correction | 2D |
| Normalize | Prescan |
| Image Filter | Off |

### Geometry - Common

| | |
|---|---|
| Slice Group | 1 |
| Slices | 1 |
| Distance Factor | 20 % |
| Position | R3.0 A40.6 H27.5 mm |
| Orientation | T > C-4.2 > S3.7 |
| Phase Encoding Dir. | A >> P |
| Slice Group | 2 |
| Slices | 1 |
| Distance Factor | 20 % |
| Position | R3.0 A40.6 H27.5 mm |
| Orientation | T > C-4.2 > S3.7 |
| Phase Encoding Dir. | A >> P |
| Slice Group | 3 |
| Slices | 1 |
| Distance Factor | 20 % |
| Position | R3.0 A40.6 H27.5 mm |
| Orientation | T > C-4.2 > S3.7 |
| Phase Encoding Dir. | A >> P |
| Phase Oversampling | 0 % |
| FoV Read | 300 mm |
| FoV Phase | 100.0 % |
| Slice Thickness | 5.0 mm |
| TR | 104.0 ms |
| Multi-Slice Mode | Interleaved |
| Series | Interleaved |
| Concatenations | 1 |

### Geometry - AutoAlign

| | |
|---|---|
| Slice Group | 1 |
| Position | R3.0 A40.6 H27.5 mm |
| Orientation | T > C-4.2 > S3.7 |
| Phase Encoding Dir. | A >> P |
| Slice Group | 2 |
| Position | R3.0 A40.6 H27.5 mm |
| Orientation | T > C-4.2 > S3.7 |
| Phase Encoding Dir. | A >> P |
| Slice Group | 3 |
| Position | R3.0 A40.6 H27.5 mm |
| Orientation | T > C-4.2 > S3.7 |

### Geometry - AutoAlign

| | |
|---|---|
| Phase Encoding Dir. | A >> P |
| AutoAlign | Head > Brain |
| Initial Position | L0.0 A26.5 H10.7 |
| L | 0.0 mm |
| A | 26.5 mm |
| H | 10.7 mm |
| Initial Orientation | Transversal |
| Initial Rotation | 0.00 deg |

### Geometry - Saturation

| | |
|---|---|
| Saturation Mode | Standard |
| Special Saturation | None |

### Geometry - Tim Planning Suite

| | |
|---|---|
| Set-n-Go Protocol | Off |
| Table Position | 0 mm |
| Table Position | H |
| Inline Composing | Off |

### System - Miscellaneous

| | |
|---|---|
| Coil Selection | ACS All but spine |
| MSMA | S - C - T |
| Sagittal | R >> L |
| Coronal | A >> P |
| Transversal | F >> H |
| Coil Combination | Sum of Squares |
| Matrix Optimization | Off |
| Coil Focus | Flat |

### System - Adjustments

| | |
|---|---|
| Adjustment Strategy | Standard |
| B0 Shim | Tune up |
| B1 Shim | TrueForm |
| CoilShim | Off |
| Adjustment Tolerance | Auto |
| Adjust with Body Coil | Off |
| Confirm Frequency | Never |
| Assume Silicone | Off |

### System - Adjust Volume

| | |
|---|---|
| Position | Isocenter |
| Orientation | Transversal |
| Rotation | 0.00 deg |
| A >> P | 263 mm |
| R >> L | 350 mm |
| F >> H | 350 mm |
| Reset | Off |

### System - pTx

| | |
|---|---|
| B1 Shim | TrueForm |
| Excitation | Slice-sel. |
| LR Balancing | Off |

### System - Tx/Rx

| | |
|---|---|
| Frequency 1H | 123.257916 MHz |
| ? Ref. Amplitude 1H | 0.000 V |
| Reset | Off |
| Image Scaling | 1.000 |

### Physio - Signal

| | |
|---|---|
| 1st Signal/Mode | None |
| TR | 104.0 ms |
| Segments | 1 |

### Physio - Signal

| | |
|---|---|
| Concatenations | 1 |

### Physio - Cardiac

| | |
|---|---|
| Tagging | None |
| Fat-Water Contrast | Standard |
| Magn. Preparation | None |
| Dark Blood | Off |
| FoV Read | 300 mm |
| FoV Phase | 100.0 % |
| Phase Resolution | 75 % |
| Dynamic Mode | Standard |

### Physio - PACE

| | |
|---|---|
| Resp. Control | Off |
| Concatenations | 1 |

### Inline - Liver

| | |
|---|---|
| Liver Registration | Off |
| Save Original Images | On |

### Inline - Subtraction

| | |
|---|---|
| Subtract | Off |
| Measurements | 1 |
| StdDev | Off |
| Save Original Images | On |

### Inline - Cardiac

| | |
|---|---|
| Magn. Preparation | None |
| Save Original Images | On |
| Contrasts | 1 |
| TE | 3.00 ms |
| TR | 104.0 ms |

### Inline - MIP

| | |
|---|---|
| MIP Sag | Off |
| MIP Cor | Off |
| MIP Tra | Off |
| MIP Time | Off |
| Radial MIP | Off |
| Save Original Images | On |
| MPR Sag | Off |
| MPR Cor | Off |
| MPR Tra | Off |

### Inline - Soft Tissue

| | |
|---|---|
| Wash-in | Off |
| Wash-out | Off |
| TTP | Off |
| PEI | Off |
| MIP Time | Off |
| Measurements | 1 |

### Inline - Composing

| | |
|---|---|
| Inline Composing | Off |

### Inline - MapIt

| | |
|---|---|
| MapIt | None |
| Flip Angle | 15 deg |
| Measurements | 1 |
| Contrasts | 1 |
| TE | 3.00 ms |
| TR | 104.0 ms |
| Save Original Images | On |

### Sequence - Part 1

| | |
|---|---|
| Sequence Name | fl |
| Dimension | 2D |
| Excitation | Slice-sel. |
| RF Pulse Type | Normal |
| Gradient Mode | Fast |
| Flow Compensation | None |
| Bandwidth | 260 Hz/Px |
| Asymmetric Echo | Allowed |
| Segments | 1 |

### Sequence - Part 2

| | |
|---|---|
| Introduction | On |
| RF Spoiling | On |
| Acoustic noise reduction | Off |

### Sequence - Assistant

| | |
|---|---|
| SAR Assistant | Off |
| Allowed Delay | 0 s |

| \\PHYSICS\XA30 RESEARCH PROT\PARCELLATION\BRAINGATE2 08-26-2024\Sag_T1_3D_Mprage |
|---|
| TA: 5:21 min Coil Selection: Auto Voxel Size: 1.0×1.0×0.9 mm³ Acc:: 2 Rel. SNR: 1.00 |

### Properties

| | |
|---|---|
| Start measurement without further preparation | On |
| Wait for User to Start | Off |
| Start measurements | Single Measurement |
| Prio Recon | Off |
| Auto Open Inline Display | Off |
| Auto Close Inline Display | Off |
| Load Images to MR View&GO | On |
| Auto Store Images | On |
| Load Images to Stamp Segments | Off |
| Load Images to Graphic Segments | Off |
| Graphic segment | Default |
| Inline Movie | Off |

### Routine

| | |
|---|---|
| Slab Group | 1 |
| Slabs | 1 |
| Distance Factor | 50 % |
| Position | R1.6 A29.4 F10.9 mm |
| Orientation | S > T-2.8 > C-0.8 |
| Phase Encoding Dir. | A >> P |
| Slices per Slab | 192 |
| Phase Oversampling | 0 % |
| Slice Oversampling | 16.7 % |
| FoV Read | 260 mm |
| FoV Phase | 100.0 % |
| Slice Thickness | 0.9 mm |
| TR | 2300.0 ms |
| TE | 2.32 ms |
| Averages | 1 |
| Concatenations | 1 |
| AutoAlign | Head > Basis |

### Contrast - Common

| | |
|---|---|
| TR | 2300.0 ms |
| TE | 2.32 ms |
| Magn. Preparation | Non-sel. IR |
| TI | 900 ms |
| Flip Angle | 8 deg |
| Fat-Water Contrast | Standard |
| Dark Blood | Off |
| Reconstruction | Magnitude |

### Contrast - Dynamic

| | |
|---|---|
| Dynamic Mode | Standard |
| Measurements | 1 |
| Multiple Series | Each Measurement |
| Reordering | Linear |

### Resolution - Common

| | |
|---|---|
| FoV Read | 260 mm |
| FoV Phase | 100.0 % |
| Slice Thickness | 0.9 mm |
| Base Resolution | 256 |
| Phase Resolution | 100 % |
| Slice Resolution | 100 % |
| Interpolation | Off |

### Resolution - Acceleration

| | |
|---|---|
| Acceleration mode | GRAPPA |

### Resolution - Acceleration

| | |
|---|---|
| Reference Scans | Integrated |
| Acceleration Factor PE | 2 |
| Reference Lines PE | 24 |
| Acceleration Factor 3D | 1 |
| Phase Partial Fourier | Off |
| Slice Partial Fourier | Off |
| Asymmetric Echo | Allowed |
| Elliptical Scanning | Off |

### Resolution - Filter

| | |
|---|---|
| Raw Filter | Off |
| Elliptical Filter | Off |
| Distortion Correction | 2D |
| Normalize | Prescan |
| Image Filter | Off |

### Geometry - Common

| | |
|---|---|
| Slab Group | 1 |
| Slabs | 1 |
| Distance Factor | 50 % |
| Position | R1.6 A29.4 F10.9 mm |
| Orientation | S > T-2.8 > C-0.8 |
| Phase Encoding Dir. | A >> P |
| Slices per Slab | 192 |
| Phase Oversampling | 0 % |
| Slice Oversampling | 16.7 % |
| FoV Read | 260 mm |
| FoV Phase | 100.0 % |
| Slice Thickness | 0.9 mm |
| TR | 2300.0 ms |
| Multi-Slice Mode | Single Shot |
| Series | Ascending |
| Concatenations | 1 |

### Geometry - AutoAlign

| | |
|---|---|
| Slab Group | 1 |
| Position | R1.6 A29.4 F10.9 mm |
| Orientation | S > T-2.8 > C-0.8 |
| Phase Encoding Dir. | A >> P |
| AutoAlign | Head > Basis |
| Initial Position | L0.4 P0.4 H3.5 |
| L | 0.4 mm |
| P | 0.4 mm |
| H | 3.5 mm |
| Initial Orientation | S > T |
| S > T | 0.80 |
| > C | 0.10 |
| Initial Rotation | 6.57 deg |

### Geometry - Navigator

### Geometry - Tim Planning Suite

| | |
|---|---|
| Set-n-Go Protocol | Off |
| Table Position | 0 mm |
| Table Position | H |
| Inline Composing | Off |

### System - Miscellaneous

| | |
|---|---|
| Coil Selection | ACS All but spine |
| MSMA | S - C - T |

### System - Miscellaneous

| | |
|---|---|
| Sagittal | R >> L |
| Coronal | A >> P |
| Transversal | F >> H |
| Coil Combination | Adaptive Combine |
| Matrix Optimization | Off |
| Coil Focus | Flat |

### System - Adjustments

| | |
|---|---|
| Adjustment Strategy | Standard |
| B0 Shim | Tune up |
| B1 Shim | TrueForm |
| CoilShim | Off |
| Adjustment Tolerance | Auto |
| Adjust with Body Coil | Off |
| Confirm Frequency | Never |
| Assume Silicone | Off |

### System - Adjust Volume

| | |
|---|---|
| Position | Isocenter |
| Orientation | Transversal |
| Rotation | 0.00 deg |
| A >> P | 263 mm |
| R >> L | 350 mm |
| F >> H | 350 mm |
| Reset | Off |

### System - pTx

| | |
|---|---|
| B1 Shim | TrueForm |
| Excitation | Non-sel. |

### System - Tx/Rx

| | |
|---|---|
| Frequency 1H | 123.257916 MHz |
| ? Ref. Amplitude 1H | 0.000 V |
| Reset | Off |
| Image Scaling | 1.000 |

### Physio - Signal

| | |
|---|---|
| 1st Signal/Mode | None |
| TR | 2300.0 ms |
| Concatenations | 1 |

### Physio - Cardiac

| | |
|---|---|
| Fat-Water Contrast | Standard |
| Magn. Preparation | Non-sel. IR |
| TI | 900 ms |
| Dark Blood | Off |
| FoV Read | 260 mm |
| FoV Phase | 100.0 % |
| Phase Resolution | 100 % |
| Dynamic Mode | Standard |

### Physio - PACE

| | |
|---|---|
| Resp. Control | Off |
| Concatenations | 1 |

### Inline - Subtraction

| | |
|---|---|
| Subtract | Off |
| Measurements | 1 |
| StdDev | Off |
| Save Original Images | On |

### Inline - Cardiac

| | |
|---|---|
| Magn. Preparation | Non-sel. IR |
| Save Original Images | On |
| TE | 2.32 ms |
| TR | 2300.0 ms |

### Inline - MIP

| | |
|---|---|
| MIP Sag | Off |
| MIP Cor | Off |
| MIP Tra | Off |
| MIP Time | Off |
| Radial MIP | Off |
| Save Original Images | On |
| MPR Sag | Off |
| MPR Cor | Off |
| MPR Tra | Off |

### Inline - Composing

| | |
|---|---|
| Inline Composing | Off |

### Inline - MapIt

| | |
|---|---|
| MapIt | None |
| Flip Angle | 8 deg |
| Measurements | 1 |
| TE | 2.32 ms |
| TR | 2300.0 ms |
| Save Original Images | On |

### Sequence - Part 1

| | |
|---|---|
| Sequence Name | tfl |
| Dimension | 3D |
| Excitation | Non-sel. |
| RF Pulse Type | Normal |
| Gradient Mode | Normal |
| Flow Compensation | None |
| Reordering | Linear |
| Bandwidth | 200 Hz/Px |
| Echo Spacing | 6.86 ms |
| Asymmetric Echo | Allowed |
| Turbo Factor | 224 |

### Sequence - Part 2

| | |
|---|---|
| Introduction | On |
| RF Spoiling | On |
| Incr. Gradient Spoiling | Off |

### Sequence - Assistant

| | |
|---|---|
| SAR Assistant | Off |

### \\PHYSICS\XA30 RESEARCH PROT\PARCELLATION\BRAINGATE2 08-26-2024\Sag_T2_3D_Space_Wave

TA: 1:38 min Coil Selection: Auto Voxel Size: 1.0×1.0×1.0 mm³ Acc:: 6 Rel. SNR: 1.00

#### Properties

| | |
|---|---|
| Start measurement without further preparation | On |
| Wait for User to Start | Off |
| Start measurements | Single Measurement |
| Prio Recon | Off |
| Auto Open Inline Display | Off |
| Auto Close Inline Display | Off |
| Load Images to MR View&GO | On |
| Auto Store Images | On |
| Load Images to Stamp Segments | Off |
| Load Images to Graphic Segments | Off |
| Graphic segment | Default |
| Inline Movie | Off |

#### Routine

| | |
|---|---|
| Slab Group | 1 |
| Slabs | 1 |
| Position | R1.6 A29.4 F10.9 mm |
| Orientation | S > T-2.8 > C-0.8 |
| Phase Encoding Dir. | A >> P |
| Slices per Slab | 224 |
| Phase Oversampling | 0 % |
| Slice Oversampling | 0.0 % |
| FoV Read | 256 mm |
| FoV Phase | 100.0 % |
| Slice Thickness | 1.00 mm |
| TR | 3200.0 ms |
| TE | 407.00 ms |
| Averages | 1.0 |
| Concatenations | 1 |
| AutoAlign | Head > Basis |

#### Contrast - Common

| | |
|---|---|
| TR | 3200.0 ms |
| TE | 407.00 ms |
| MTC | Off |
| Magn. Preparation | None |
| Flip Angle Mode | T2 Var |
| Fat-Water Contrast | Standard |
| Dark Blood | Off |
| Blood Suppression | Off |
| Wrap-up Magn. | Restore |
| Reconstruction | Magnitude |

#### Contrast - Dynamic

| | |
|---|---|
| Dynamic Mode | Standard |
| Measurements | 1 |
| Multiple Series | Each Measurement |
| Reordering | Linear |

#### Resolution - Common

| | |
|---|---|
| FoV Read | 256 mm |
| FoV Phase | 100.0 % |
| Slice Thickness | 1.00 mm |
| Base Resolution | 256 |
| Phase Resolution | 100 % |
| Slice Resolution | 100 % |
| Interpolation | Off |

#### Resolution - Acceleration

| | |
|---|---|
| Acceleration mode | CAIPIRINHA |
| Total Factor | 6 |
| Reference Scans | GRE/Separate |
| Acceleration Factor PE | 3 |
| Reference Lines PE | 24 |
| Acceleration Factor 3D | 2 |
| Reference Lines 3D | 48 |
| Reordering Shift 3D | 0 |
| Phase Partial Fourier | Allowed |
| Slice Partial Fourier | Off |
| Elliptical Scanning | On |

#### Resolution - Filter

| | |
|---|---|
| Distortion Correction | 3D |
| Normalize | Prescan |
| Image Filter | Off |

#### Geometry - Common

| | |
|---|---|
| Slab Group | 1 |
| Slabs | 1 |
| Position | R1.6 A29.4 F10.9 mm |
| Orientation | S > T-2.8 > C-0.8 |
| Phase Encoding Dir. | A >> P |
| Slices per Slab | 224 |
| Phase Oversampling | 0 % |
| Slice Oversampling | 0.0 % |
| FoV Read | 256 mm |
| FoV Phase | 100.0 % |
| Slice Thickness | 1.00 mm |
| TR | 3200.0 ms |
| Concatenations | 1 |

#### Geometry - AutoAlign

| | |
|---|---|
| Slab Group | 1 |
| Position | R1.6 A29.4 F10.9 mm |
| Orientation | S > T-2.8 > C-0.8 |
| Phase Encoding Dir. | A >> P |
| AutoAlign | Head > Basis |
| Initial Position | L0.4 P0.4 H3.5 |
| L | 0.4 mm |
| P | 0.4 mm |
| H | 3.5 mm |
| Initial Orientation | S > T |
| S > T | 0.80 |
| > C | 0.10 |
| Initial Rotation | 6.56 deg |

#### Geometry - Navigator

#### Geometry - Saturation

| | |
|---|---|
| Special Saturation | None |

#### Geometry - Tim Planning Suite

| | |
|---|---|
| Set-n-Go Protocol | Off |
| Table Position | 0 mm |
| Table Position | H |
| Inline Composing | Off |

### System - Miscellaneous

| | |
|---|---|
| Coil Selection | Auto Coil Select |
| MSMA | S - C - T |
| Sagittal | R >> L |
| Coronal | A >> P |
| Transversal | F >> H |
| Coil Combination | Sum of Squares |
| Matrix Optimization | Performance |
| Coil Focus | Flat |

### System - Adjustments

| | |
|---|---|
| Adjustment Strategy | Standard |
| B0 Shim | Tune up |
| B1 Shim | TrueForm |
| CoilShim | Off |
| Adjustment Tolerance | Auto |
| Adjust with Body Coil | Off |
| Confirm Frequency | Never |
| Assume Silicone | Off |

### System - Adjust Volume

| | |
|---|---|
| Position | Isocenter |
| Orientation | Transversal |
| Rotation | 0.00 deg |
| A >> P | 263 mm |
| R >> L | 350 mm |
| F >> H | 350 mm |
| Reset | Off |

### System - pTx

| | |
|---|---|
| B1 Shim | TrueForm |
| Excitation | Non-sel. |

### System - Tx/Rx

| | |
|---|---|
| Frequency 1H | 123.257916 MHz |
| ? Ref. Amplitude 1H | 0.000 V |
| Reset | Off |
| Image Scaling | 1.000 |

### Physio - Signal

| | |
|---|---|
| 1st Signal/Mode | None |
| Trigger Delay | 0 ms |
| TR | 3200.0 ms |
| Concatenations | 1 |

### Physio - Cardiac

| | |
|---|---|
| Fat-Water Contrast | Standard |
| Magn. Preparation | None |
| Dark Blood | Off |
| FoV Read | 256 mm |
| FoV Phase | 100.0 % |
| Phase Resolution | 100 % |
| Dynamic Mode | Standard |

### Physio - PACE

| | |
|---|---|
| Resp. Control | Off |
| Concatenations | 1 |

### Inline - Subtraction

| | |
|---|---|
| Subtract | Off |
| Measurements | 1 |
| StdDev | Off |
| Save Original Images | On |

### Inline - Cardiac

| | |
|---|---|
| Magn. Preparation | None |
| Save Original Images | On |
| TE | 407.00 ms |
| TR | 3200.0 ms |

### Inline - MIP

| | |
|---|---|
| MIP Sag | Off |
| MIP Cor | Off |
| MIP Tra | Off |
| MIP Time | Off |
| Radial MIP | Off |
| Save Original Images | On |
| MPR Sag | Off |
| MPR Cor | Off |
| MPR Tra | Off |

### Inline - Composing

| | |
|---|---|
| Inline Composing | Off |

### Sequence - Part 1

| | |
|---|---|
| Sequence Name | WIP_spcR |
| Dimension | 3D |
| Excitation | Non-sel. |
| RF Pulse Type | Normal |
| Gradient Mode | Fast |
| Flow Compensation | None |
| Reordering | Linear |
| Bandwidth | 651 Hz/Px |
| Echo Spacing | 3.84 ms |
| Turbo Factor | 282 |
| Echo Train Duration | 952 ms |

### Sequence - Part 2

| | |
|---|---|
| Introduction | On |

### Sequence - Special

| | |
|---|---|
| Play Blip | Yes |
| Wave | Both |
| Coil Mixing | On |
| ROFilt Pre | 10 perc |
| ROFilt Post | 0 perc |
| LINFilt Pre | 0 perc |
| LINFilt Post | 0 perc |
| PARFilt Pre | 10 perc |
| PARFilt Post | 10 perc |
| Reg. Beta | 0.20 |
| Reg. Lambda | 0.20 |
| MAX Slew wave | 160.0 mT/m/ms |
| MAX Ampl wave | 20.0 mT/m |
| NumOfCycles Wave | 5.0 |

### Sequence - Assistant

| | |
|---|---|
| SAR Assistant | Off |
| Allowed Delay | 0 s |

**\\PHYSICS\XA30 RESEARCH PROT\PARCELLATION\BRAINGATE2 08-26-2024\SpinEchoFieldMap_1_AP**

TA: 8 sec Coil Selection: Auto Voxel Size: 2.0×2.0×2.0 mm³ Acc:: None Rel. SNR: 1.00

### Properties

| | |
|---|---|
| Start measurement without further preparation | On |
| Wait for User to Start | On |
| Start measurements | Single Measurement |
| Prio Recon | Off |
| Auto Open Inline Display | Off |
| Auto Close Inline Display | Off |
| Load Images to MR View&GO | On |
| Auto Store Images | On |
| Load Images to Stamp Segments | Off |
| Load Images to Graphic Segments | Off |
| Graphic segment | Default |
| Inline Movie | Off |

### Routine

| | |
|---|---|
| Slice Group | 1 |
| Slices | 72 |
| Distance Factor | 0 % |
| Position | R4.9 A16.5 H29.7 mm |
| Orientation | T > C-4.3 > S3.3 |
| Phase Encoding Dir. | A >> P |
| Phase Oversampling | 0 % |
| FoV Read | 208 mm |
| FoV Phase | 100.0 % |
| Slice Thickness | 2.0 mm |
| TR | 8000.0 ms |
| TE | 66.00 ms |
| Averages | 1 |
| Multi-band accel. factor | 1 |
| AutoAlign | Head > Brain |

### Contrast - Common

| | |
|---|---|
| TR | 8000.0 ms |
| TE | 66.00 ms |
| MTC | Off |
| Magn. Preparation | None |
| Flip Angle | 90 deg |
| Refocus flip angle | 180 deg |
| Fat-Water Contrast | Fat Saturation |
| Grad. rev. fat suppr. | Disabled |
| Contrasts | 1 |
| Reconstruction | Magnitude |

### Contrast - Dynamic

| | |
|---|---|
| Dynamic Mode | Standard |
| Measurements | 1 |
| Delay in TR | 0.00 ms |

### Resolution - Common

| | |
|---|---|
| FoV Read | 208 mm |
| FoV Phase | 100.0 % |
| Slice Thickness | 2.0 mm |
| Base Resolution | 104 |
| Phase Resolution | 100 % |
| Interpolation | Off |

### Resolution - Acceleration

| | |
|---|---|
| Acceleration mode | None |
| Advanced Reconstruction | Off |

### Resolution - Acceleration

| | |
|---|---|
| Phase Partial Fourier | Off |

### Resolution - Filter

| | |
|---|---|
| Raw Filter | Off |
| Elliptical Filter | Off |
| Hamming | Off |
| Distortion Correction | Off |
| Normalize | Off |

### Geometry - Common

| | |
|---|---|
| Slice Group | 1 |
| Slices | 72 |
| Distance Factor | 0 % |
| Position | R4.9 A16.5 H29.7 mm |
| Orientation | T > C-4.3 > S3.3 |
| Phase Encoding Dir. | A >> P |
| Phase Oversampling | 0 % |
| FoV Read | 208 mm |
| FoV Phase | 100.0 % |
| Slice Thickness | 2.0 mm |
| TR | 8000.0 ms |
| Multi-Slice Mode | Interleaved |
| Series | Interleaved |
| Multi-band accel. factor | 1 |

### Geometry - AutoAlign

| | |
|---|---|
| Slice Group | 1 |
| Position | R4.9 A16.5 H29.7 mm |
| Orientation | T > C-4.3 > S3.3 |
| Phase Encoding Dir. | A >> P |
| AutoAlign | Head > Brain |
| Initial Position | R1.4 A2.6 H14.8 |
| R | 1.4 mm |
| A | 2.6 mm |
| H | 14.8 mm |
| Initial Orientation | T > S |
| T > S | -0.40 |
| > C | -0.10 |
| Initial Rotation | 0.14 deg |

### Geometry - Saturation

| | |
|---|---|
| Special Saturation | None |

### Geometry - Tim Planning Suite

| | |
|---|---|
| Set-n-Go Protocol | Off |
| Table Position | 0 mm |
| Table Position | H |
| Inline Composing | Off |

### System - Miscellaneous

| | |
|---|---|
| Coil Selection | ACS All but spine |
| MSMA | S - C - T |
| Sagittal | R >> L |
| Coronal | A >> P |
| Transversal | F >> H |
| Coil Combination | Sum of Squares |
| Matrix Optimization | Off |
| Coil Focus | Flat |

### System - Adjustments

| | |
|---|---|
| Adjustment Strategy | Standard |
| B0 Shim | Standard |
| B1 Shim | TrueForm |
| CoilShim | Off |
| Adjustment Tolerance | Auto |
| Adjust with Body Coil | Off |
| Confirm Frequency | Never |
| Assume Silicone | Off |

### System - Adjust Volume

| | |
|---|---|
| Position | R4.9 A16.5 H29.7 mm |
| Orientation | T > C-4.3 > S3.3 |
| Rotation | -0.44 deg |
| A >> P | 208 mm |
| R >> L | 208 mm |
| F >> H | 144 mm |
| Reset | Off |

### System - pTx

| | |
|---|---|
| B1 Shim | TrueForm |
| Excitation | Standard |

### System - Tx/Rx

| | |
|---|---|
| Frequency 1H | 123.257916 MHz |
| ? Ref. Amplitude 1H | 0.000 V |
| Reset | Off |
| Image Scaling | 1.000 |

### Physio - Signal

| | |
|---|---|
| 1st Signal/Mode | None |
| TR | 8000.0 ms |
| Multi-band accel. factor | 1 |

### BOLD

| | |
|---|---|
| GLM Statistics | Off |
| Ignore Meas. at Start | 0 |
| Ignore After Transition | 0 |
| Model Transition States | Off |
| Temp. Highpass Filter | Off |
| Threshold | 4.00 |
| Paradigm Size | 3 |
| Meas[1] | Active |
| Meas[2] | Active |
| Meas[3] | Ignore |
| Motion Correction | Off |
| Spatial Filter | Off |
| Measurements | 1 |
| Delay in TR | 0.00 ms |

### Sequence - Part 1

| | |
|---|---|
| Sequence Name | epse |
| Dimension | 2D |
| Excitation | Standard |
| RF Pulse Type | Normal |
| Gradient Mode | Performance |
| Bandwidth | 2290 Hz/Px |
| Echo Spacing | 0.58 ms |
| Free Echo Spacing | On |
| EPI Factor | 104 |

### Sequence - Part 2

| | |
|---|---|
| Introduction | Off |

### Sequence - Special

| | |
|---|---|
| Min. prep scans | 0 |
| Delay before PC scans | 0 us |
| SENSE1 coil combine | Off |
| Invert RO/PE polarity | Off |
| Disable freq. update | Off |
| Suppress 16-bit DICOM | Off |
| Force equal slice timing | Off |
| FFT scale factor | 1.00 |
| Fat saturation FA | 110.00 deg |
| Fat sat. offset | 0.00 Hz |
| Physio recording | Off |
| Triggering scheme | Standard |

### Sequence - Assistant

| | |
|---|---|
| SAR Assistant | Off |

**\\PHYSICS\XA30 RESEARCH PROT\PARCELLATION\BRAINGATE2 08-26-2024\SpinEchoFieldMap_1_PA**

TA: 8 sec Coil Selection: Auto Voxel Size: 2.0×2.0×2.0 mm³ Acc:: None Rel. SNR: 1.00

### Properties

| | |
|---|---|
| Start measurement without further preparation | On |
| Wait for User to Start | Off |
| Start measurements | Single Measurement |
| Prio Recon | Off |
| Auto Open Inline Display | Off |
| Auto Close Inline Display | Off |
| Load Images to MR View&GO | On |
| Auto Store Images | On |
| Load Images to Stamp Segments | Off |
| Load Images to Graphic Segments | Off |
| Graphic segment | Default |
| Inline Movie | Off |

### Routine

| | |
|---|---|
| Slice Group | 1 |
| Slices | 72 |
| Distance Factor | 0 % |
| Position | R4.9 A16.5 H29.7 mm |
| Orientation | T > C-4.3 > S3.3 |
| Phase Encoding Dir. | P >> A |
| Phase Oversampling | 0 % |
| FoV Read | 208 mm |
| FoV Phase | 100.0 % |
| Slice Thickness | 2.0 mm |
| TR | 8000.0 ms |
| TE | 66.00 ms |
| Averages | 1 |
| Multi-band accel. factor | 1 |
| AutoAlign | Head > Brain |

### Contrast - Common

| | |
|---|---|
| TR | 8000.0 ms |
| TE | 66.00 ms |
| MTC | Off |
| Magn. Preparation | None |
| Flip Angle | 90 deg |
| Refocus flip angle | 180 deg |
| Fat-Water Contrast | Fat Saturation |
| Grad. rev. fat suppr. | Disabled |
| Contrasts | 1 |
| Reconstruction | Magnitude |

### Contrast - Dynamic

| | |
|---|---|
| Dynamic Mode | Standard |
| Measurements | 1 |
| Delay in TR | 0.00 ms |

### Resolution - Common

| | |
|---|---|
| FoV Read | 208 mm |
| FoV Phase | 100.0 % |
| Slice Thickness | 2.0 mm |
| Base Resolution | 104 |
| Phase Resolution | 100 % |
| Interpolation | Off |

### Resolution - Acceleration

| | |
|---|---|
| Acceleration mode | None |
| Advanced Reconstruction | Off |

### Resolution - Acceleration

| | |
|---|---|
| Phase Partial Fourier | Off |

### Resolution - Filter

| | |
|---|---|
| Raw Filter | Off |
| Elliptical Filter | Off |
| Hamming | Off |
| Distortion Correction | Off |
| Normalize | Off |

### Geometry - Common

| | |
|---|---|
| Slice Group | 1 |
| Slices | 72 |
| Distance Factor | 0 % |
| Position | R4.9 A16.5 H29.7 mm |
| Orientation | T > C-4.3 > S3.3 |
| Phase Encoding Dir. | P >> A |
| Phase Oversampling | 0 % |
| FoV Read | 208 mm |
| FoV Phase | 100.0 % |
| Slice Thickness | 2.0 mm |
| TR | 8000.0 ms |
| Multi-Slice Mode | Interleaved |
| Series | Interleaved |
| Multi-band accel. factor | 1 |

### Geometry - AutoAlign

| | |
|---|---|
| Slice Group | 1 |
| Position | R4.9 A16.5 H29.7 mm |
| Orientation | T > C-4.3 > S3.3 |
| Phase Encoding Dir. | P >> A |
| AutoAlign | Head > Brain |
| Initial Position | R1.4 A2.6 H14.8 |
| R | 1.4 mm |
| A | 2.6 mm |
| H | 14.8 mm |
| Initial Orientation | T > S |
| T > S | -0.40 |
| > C | -0.10 |
| Initial Rotation | -180.00 deg |

### Geometry - Saturation

| | |
|---|---|
| Special Saturation | None |

### Geometry - Tim Planning Suite

| | |
|---|---|
| Set-n-Go Protocol | Off |
| Table Position | 0 mm |
| Table Position | H |
| Inline Composing | Off |

### System - Miscellaneous

| | |
|---|---|
| Coil Selection | ACS All but spine |
| MSMA | S - C - T |
| Sagittal | R >> L |
| Coronal | A >> P |
| Transversal | F >> H |
| Coil Combination | Sum of Squares |
| Matrix Optimization | Off |
| Coil Focus | Flat |

### System - Adjustments

| | |
|---|---|
| Adjustment Strategy | Standard |
| B0 Shim | Standard |
| B1 Shim | TrueForm |
| CoilShim | Off |
| Adjustment Tolerance | Auto |
| Adjust with Body Coil | Off |
| Confirm Frequency | Never |
| Assume Silicone | Off |

### System - Adjust Volume

| | |
|---|---|
| Position | R4.9 A16.5 H29.7 mm |
| Orientation | T > C-4.3 > S3.3 |
| Rotation | 179.42 deg |
| A >> P | 208 mm |
| R >> L | 208 mm |
| F >> H | 144 mm |
| Reset | Off |

### System - pTx

| | |
|---|---|
| B1 Shim | TrueForm |
| Excitation | Standard |

### System - Tx/Rx

| | |
|---|---|
| Frequency 1H | 123.257916 MHz |
| ? Ref. Amplitude 1H | 0.000 V |
| Reset | Off |
| Image Scaling | 1.000 |

### Physio - Signal

| | |
|---|---|
| 1st Signal/Mode | None |
| TR | 8000.0 ms |
| Multi-band accel. factor | 1 |

### BOLD

| | |
|---|---|
| GLM Statistics | Off |
| Ignore Meas. at Start | 0 |
| Ignore After Transition | 0 |
| Model Transition States | Off |
| Temp. Highpass Filter | Off |
| Threshold | 4.00 |
| Paradigm Size | 3 |
| Meas[1] | Active |
| Meas[2] | Active |
| Meas[3] | Ignore |
| Motion Correction | Off |
| Spatial Filter | Off |
| Measurements | 1 |
| Delay in TR | 0.00 ms |

### Sequence - Part 1

| | |
|---|---|
| Sequence Name | epse |
| Dimension | 2D |
| Excitation | Standard |
| RF Pulse Type | Normal |
| Gradient Mode | Performance |
| Bandwidth | 2290 Hz/Px |
| Echo Spacing | 0.58 ms |
| Free Echo Spacing | On |
| EPI Factor | 104 |

### Sequence - Part 2

| | |
|---|---|
| Introduction | Off |

### Sequence - Special

| | |
|---|---|
| Min. prep scans | 0 |
| Delay before PC scans | 0 us |
| SENSE1 coil combine | Off |
| Invert RO/PE polarity | On |
| Disable freq. update | Off |
| Suppress 16-bit DICOM | Off |
| Force equal slice timing | Off |
| FFT scale factor | 1.00 |
| Fat saturation FA | 110.00 deg |
| Fat sat. offset | 0.00 Hz |
| Physio recording | Off |
| Triggering scheme | Standard |

### Sequence - Assistant

| | |
|---|---|
| SAR Assistant | Off |

| \\PHYSICS\XA30 RESEARCH PROT\PARCELLATION\BRAINGATE2 08-26-2024\rfMRI_REST_1_AP |
|---|
| TA: 6:40 min Coil Selection: Auto Voxel Size: 2.0×2.0×2.0 mm³ Acc:: None Rel. SNR: 1.00 |

### Properties

| | |
|---|---|
| Start measurement without further preparation | On |
| Wait for User to Start | Off |
| Start measurements | Single Measurement |
| Prio Recon | Off |
| Auto Open Inline Display | Off |
| Auto Close Inline Display | Off |
| Load Images to MR View&GO | On |
| Auto Store Images | On |
| Load Images to Stamp Segments | Off |
| Load Images to Graphic Segments | Off |
| Graphic segment | Default |
| Inline Movie | Off |

### Routine

| | |
|---|---|
| Slice Group | 1 |
| Slices | 72 |
| Distance Factor | 0 % |
| Position | R4.9 A16.5 H29.7 mm |
| Orientation | T > C-4.3 > S3.3 |
| Phase Encoding Dir. | A >> P |
| Phase Oversampling | 0 % |
| FoV Read | 208 mm |
| FoV Phase | 100.0 % |
| Slice Thickness | 2.0 mm |
| TR | 800.0 ms |
| TE | 37.00 ms |
| Averages | 1 |
| Multi-band accel. factor | 8 |
| AutoAlign | Head > Brain |

### Contrast - Common

| | |
|---|---|
| TR | 800.0 ms |
| TE | 37.00 ms |
| MTC | Off |
| Magn. Preparation | None |
| Flip Angle | 52 deg |
| Fat-Water Contrast | Fat Saturation |
| Contrasts | 1 |
| Reconstruction | Magnitude |

### Contrast - Dynamic

| | |
|---|---|
| Dynamic Mode | Standard |
| Measurements | 488 |
| Delay in TR | 0.00 ms |

### Resolution - Common

| | |
|---|---|
| FoV Read | 208 mm |
| FoV Phase | 100.0 % |
| Slice Thickness | 2.0 mm |
| Base Resolution | 104 |
| Phase Resolution | 100 % |
| Interpolation | Off |

### Resolution - Acceleration

| | |
|---|---|
| Acceleration mode | None |
| Advanced Reconstruction | Off |
| Phase Partial Fourier | Off |

### Resolution - Filter

| | |
|---|---|
| Raw Filter | Off |
| Elliptical Filter | Off |
| Hamming | Off |
| Distortion Correction | 2D |
| Normalize | Off |

### Geometry - Common

| | |
|---|---|
| Slice Group | 1 |
| Slices | 72 |
| Distance Factor | 0 % |
| Position | R4.9 A16.5 H29.7 mm |
| Orientation | T > C-4.3 > S3.3 |
| Phase Encoding Dir. | A >> P |
| Phase Oversampling | 0 % |
| FoV Read | 208 mm |
| FoV Phase | 100.0 % |
| Slice Thickness | 2.0 mm |
| TR | 800.0 ms |
| Multi-Slice Mode | Interleaved |
| Series | Interleaved |
| Multi-band accel. factor | 8 |

### Geometry - AutoAlign

| | |
|---|---|
| Slice Group | 1 |
| Position | R4.9 A16.5 H29.7 mm |
| Orientation | T > C-4.3 > S3.3 |
| Phase Encoding Dir. | A >> P |
| AutoAlign | Head > Brain |
| Initial Position | R1.4 A2.6 H14.8 |
| R | 1.4 mm |
| A | 2.6 mm |
| H | 14.8 mm |
| Initial Orientation | T > S |
| T > S | -0.40 |
| > C | -0.10 |
| Initial Rotation | 0.14 deg |

### Geometry - Saturation

| | |
|---|---|
| Special Saturation | None |

### Geometry - Tim Planning Suite

| | |
|---|---|
| Set-n-Go Protocol | Off |
| Table Position | 0 mm |
| Table Position | H |
| Inline Composing | Off |

### System - Miscellaneous

| | |
|---|---|
| Coil Selection | Auto Coil Select |
| MSMA | S - C - T |
| Sagittal | R >> L |
| Coronal | A >> P |
| Transversal | F >> H |
| Coil Combination | Sum of Squares |
| Matrix Optimization | Off |
| Coil Focus | Flat |

### System - Adjustments

| | |
|---|---|
| Adjustment Strategy | Standard |
| B0 Shim | Standard |
| B1 Shim | TrueForm |

### System - Adjustments

| | |
|---|---|
| CoilShim | Off |
| Adjustment Tolerance | Auto |
| Adjust with Body Coil | Off |
| Confirm Frequency | Never |
| Assume Silicone | Off |

### System - Adjust Volume

| | |
|---|---|
| Position | R4.9 A16.5 H29.7 mm |
| Orientation | T > C-4.3 > S3.3 |
| Rotation | -0.44 deg |
| A >> P | 208 mm |
| R >> L | 208 mm |
| F >> H | 144 mm |
| Reset | Off |

### System - pTx

| | |
|---|---|
| B1 Shim | TrueForm |
| Excitation | Standard |

### System - Tx/Rx

| | |
|---|---|
| Frequency 1H | 123.257916 MHz |
| ? Ref. Amplitude 1H | 0.000 V |
| Reset | Off |
| Image Scaling | 1.000 |

### Physio - Signal

| | |
|---|---|
| 1st Signal/Mode | None |
| TR | 800.0 ms |
| Multi-band accel. factor | 8 |

### BOLD

| | |
|---|---|
| GLM Statistics | Off |
| Ignore Meas. at Start | 0 |
| Ignore After Transition | 0 |
| Model Transition States | On |
| Temp. Highpass Filter | On |
| Threshold | 4.00 |
| Paradigm Size | 3 |
| Meas[1] | Active |
| Meas[2] | Active |
| Meas[3] | Ignore |
| Motion Correction | Off |
| Spatial Filter | Off |
| Measurements | 488 |
| Delay in TR | 0.00 ms |

### Sequence - Part 1

| | |
|---|---|
| Sequence Name | epfid |
| Dimension | 2D |
| Excitation | Standard |
| Gradient Mode | Performance |
| Flow Compensation | None |
| Bandwidth | 2290 Hz/Px |
| Echo Spacing | 0.58 ms |
| Free Echo Spacing | On |
| EPI Factor | 104 |

### Sequence - Part 2

| | |
|---|---|
| Introduction | Off |
| RF Spoiling | Off |

### Sequence - Special

| | |
|---|---|
| Excite pulse duration | 6600 us |
| Min. prep scans | 0 |
| Min. prep scans SB | 0 |
| Delay before PC scans | 0 us |
| Single-band images | Off |
| MB LeakBlock kernel | On |
| MB dual kernel | Off |
| MB RF phase scramble | Off |
| Opt. MB RF pulse BW | Off |
| SENSE1 coil combine | Off |
| Invert RO/PE polarity | Off |
| Disable freq. update | Off |
| Suppress 16-bit DICOM | Off |
| Force equal slice timing | Off |
| Online multi-band recon. | Online |
| FFT scale factor | 1.00 |
| Fat saturation FA | 110.00 deg |
| Fat sat. offset | 0.00 Hz |
| Sinc exc. pulse BWTP | 5.20 |
| Physio recording | Off |
| Triggering scheme | Standard |

### Sequence - Assistant

| | |
|---|---|
| SAR Assistant | Off |

**\\PHYSICS\XA30 RESEARCH PROT\PARCELLATION\BRAINGATE2 08-26-2024\rfMRI_REST_1_PA**

TA: 6:40 min Coil Selection: Auto Voxel Size: 2.0×2.0×2.0 mm³ Acc:: None Rel. SNR: 1.00

### Properties

| | |
|---|---|
| Start measurement without further preparation | On |
| Wait for User to Start | Off |
| Start measurements | Single Measurement |
| Prio Recon | Off |
| Auto Open Inline Display | Off |
| Auto Close Inline Display | Off |
| Load Images to MR View&GO | On |
| Auto Store Images | On |
| Load Images to Stamp Segments | Off |
| Load Images to Graphic Segments | Off |
| Graphic segment | Default |
| Inline Movie | Off |

### Routine

| | |
|---|---|
| Slice Group | 1 |
| Slices | 72 |
| Distance Factor | 0 % |
| Position | R4.9 A16.5 H29.7 mm |
| Orientation | T > C-4.3 > S3.3 |
| Phase Encoding Dir. | P >> A |
| Phase Oversampling | 0 % |
| FoV Read | 208 mm |
| FoV Phase | 100.0 % |
| Slice Thickness | 2.0 mm |
| TR | 800.0 ms |
| TE | 37.00 ms |
| Averages | 1 |
| Multi-band accel. factor | 8 |
| AutoAlign | Head > Brain |

### Contrast - Common

| | |
|---|---|
| TR | 800.0 ms |
| TE | 37.00 ms |
| MTC | Off |
| Magn. Preparation | None |
| Flip Angle | 52 deg |
| Fat-Water Contrast | Fat Saturation |
| Contrasts | 1 |
| Reconstruction | Magnitude |

### Contrast - Dynamic

| | |
|---|---|
| Dynamic Mode | Standard |
| Measurements | 488 |
| Delay in TR | 0.00 ms |

### Resolution - Common

| | |
|---|---|
| FoV Read | 208 mm |
| FoV Phase | 100.0 % |
| Slice Thickness | 2.0 mm |
| Base Resolution | 104 |
| Phase Resolution | 100 % |
| Interpolation | Off |

### Resolution - Acceleration

| | |
|---|---|
| Acceleration mode | None |
| Advanced Reconstruction | Off |
| Phase Partial Fourier | Off |

### Resolution - Filter

| | |
|---|---|
| Raw Filter | Off |
| Elliptical Filter | Off |
| Hamming | Off |
| Distortion Correction | 2D |
| Normalize | Off |

### Geometry - Common

| | |
|---|---|
| Slice Group | 1 |
| Slices | 72 |
| Distance Factor | 0 % |
| Position | R4.9 A16.5 H29.7 mm |
| Orientation | T > C-4.3 > S3.3 |
| Phase Encoding Dir. | P >> A |
| Phase Oversampling | 0 % |
| FoV Read | 208 mm |
| FoV Phase | 100.0 % |
| Slice Thickness | 2.0 mm |
| TR | 800.0 ms |
| Multi-Slice Mode | Interleaved |
| Series | Interleaved |
| Multi-band accel. factor | 8 |

### Geometry - AutoAlign

| | |
|---|---|
| Slice Group | 1 |
| Position | R4.9 A16.5 H29.7 mm |
| Orientation | T > C-4.3 > S3.3 |
| Phase Encoding Dir. | P >> A |
| AutoAlign | Head > Brain |
| Initial Position | R1.4 A2.6 H14.8 |
| R | 1.4 mm |
| A | 2.6 mm |
| H | 14.8 mm |
| Initial Orientation | T > S |
| T > S | -0.40 |
| > C | -0.10 |
| Initial Rotation | -180.00 deg |

### Geometry - Saturation

| | |
|---|---|
| Special Saturation | None |

### Geometry - Tim Planning Suite

| | |
|---|---|
| Set-n-Go Protocol | Off |
| Table Position | 0 mm |
| Table Position | H |
| Inline Composing | Off |

### System - Miscellaneous

| | |
|---|---|
| Coil Selection | Auto Coil Select |
| MSMA | S - C - T |
| Sagittal | R >> L |
| Coronal | A >> P |
| Transversal | F >> H |
| Coil Combination | Sum of Squares |
| Matrix Optimization | Off |
| Coil Focus | Flat |

### System - Adjustments

| | |
|---|---|
| Adjustment Strategy | Standard |
| B0 Shim | Standard |
| B1 Shim | TrueForm |

### System - Adjustments

| | |
|---|---|
| CoilShim | Off |
| Adjustment Tolerance | Auto |
| Adjust with Body Coil | Off |
| Confirm Frequency | Never |
| Assume Silicone | Off |

### System - Adjust Volume

| | |
|---|---|
| Position | R4.9 A16.5 H29.7 mm |
| Orientation | T > C-4.3 > S3.3 |
| Rotation | 179.42 deg |
| A >> P | 208 mm |
| R >> L | 208 mm |
| F >> H | 144 mm |
| Reset | Off |

### System - pTx

| | |
|---|---|
| B1 Shim | TrueForm |
| Excitation | Standard |

### System - Tx/Rx

| | |
|---|---|
| Frequency 1H | 123.257916 MHz |
| ? Ref. Amplitude 1H | 0.000 V |
| Reset | Off |
| Image Scaling | 1.000 |

### Physio - Signal

| | |
|---|---|
| 1st Signal/Mode | None |
| TR | 800.0 ms |
| Multi-band accel. factor | 8 |

### BOLD

| | |
|---|---|
| GLM Statistics | Off |
| Ignore Meas. at Start | 0 |
| Ignore After Transition | 0 |
| Model Transition States | On |
| Temp. Highpass Filter | On |
| Threshold | 4.00 |
| Paradigm Size | 3 |
| Meas[1] | Active |
| Meas[2] | Active |
| Meas[3] | Ignore |
| Motion Correction | Off |
| Spatial Filter | Off |
| Measurements | 488 |
| Delay in TR | 0.00 ms |

### Sequence - Part 1

| | |
|---|---|
| Sequence Name | epfid |
| Dimension | 2D |
| Excitation | Standard |
| Gradient Mode | Performance |
| Flow Compensation | None |
| Bandwidth | 2290 Hz/Px |
| Echo Spacing | 0.58 ms |
| Free Echo Spacing | On |
| EPI Factor | 104 |

### Sequence - Part 2

| | |
|---|---|
| Introduction | Off |
| RF Spoiling | Off |

### Sequence - Special

| | |
|---|---|
| Excite pulse duration | 6600 us |
| Min. prep scans | 0 |
| Min. prep scans SB | 0 |
| Delay before PC scans | 0 us |
| Single-band images | Off |
| MB LeakBlock kernel | On |
| MB dual kernel | Off |
| MB RF phase scramble | Off |
| Opt. MB RF pulse BW | Off |
| SENSE1 coil combine | Off |
| Invert RO/PE polarity | Off |
| Disable freq. update | Off |
| Suppress 16-bit DICOM | Off |
| Force equal slice timing | Off |
| Online multi-band recon. | Online |
| FFT scale factor | 1.00 |
| Fat saturation FA | 110.00 deg |
| Fat sat. offset | 0.00 Hz |
| Sinc exc. pulse BWTP | 5.20 |
| Physio recording | Off |
| Triggering scheme | Standard |

### Sequence - Assistant

| | |
|---|---|
| SAR Assistant | Off |

**\\PHYSICS\XA30 RESEARCH PROT\PARCELLATION\BRAINGATE2 08-26-2024\SpinEchoFieldMap_2_AP**

TA: 8 sec Coil Selection: Auto Voxel Size: 2.0×2.0×2.0 mm³ Acc:: None Rel. SNR: 1.00

### Properties

| | |
|---|---|
| Start measurement without further preparation | On |
| Wait for User to Start | On |
| Start measurements | Single Measurement |
| Prio Recon | Off |
| Auto Open Inline Display | Off |
| Auto Close Inline Display | Off |
| Load Images to MR View&GO | On |
| Auto Store Images | On |
| Load Images to Stamp Segments | Off |
| Load Images to Graphic Segments | Off |
| Graphic segment | Default |
| Inline Movie | Off |

### Routine

| | |
|---|---|
| Slice Group | 1 |
| Slices | 72 |
| Distance Factor | 0 % |
| Position | R4.9 A16.5 H29.7 mm |
| Orientation | T > C-4.3 > S3.3 |
| Phase Encoding Dir. | A >> P |
| Phase Oversampling | 0 % |
| FoV Read | 208 mm |
| FoV Phase | 100.0 % |
| Slice Thickness | 2.0 mm |
| TR | 8000.0 ms |
| TE | 66.00 ms |
| Averages | 1 |
| Multi-band accel. factor | 1 |
| AutoAlign | Head > Brain |

### Contrast - Common

| | |
|---|---|
| TR | 8000.0 ms |
| TE | 66.00 ms |
| MTC | Off |
| Magn. Preparation | None |
| Flip Angle | 90 deg |
| Refocus flip angle | 180 deg |
| Fat-Water Contrast | Fat Saturation |
| Grad. rev. fat suppr. | Disabled |
| Contrasts | 1 |
| Reconstruction | Magnitude |

### Contrast - Dynamic

| | |
|---|---|
| Dynamic Mode | Standard |
| Measurements | 1 |
| Delay in TR | 0.00 ms |

### Resolution - Common

| | |
|---|---|
| FoV Read | 208 mm |
| FoV Phase | 100.0 % |
| Slice Thickness | 2.0 mm |
| Base Resolution | 104 |
| Phase Resolution | 100 % |
| Interpolation | Off |

### Resolution - Acceleration

| | |
|---|---|
| Acceleration mode | None |
| Advanced Reconstruction | Off |

### Resolution - Acceleration

| | |
|---|---|
| Phase Partial Fourier | Off |

### Resolution - Filter

| | |
|---|---|
| Raw Filter | Off |
| Elliptical Filter | Off |
| Hamming | Off |
| Distortion Correction | Off |
| Normalize | Off |

### Geometry - Common

| | |
|---|---|
| Slice Group | 1 |
| Slices | 72 |
| Distance Factor | 0 % |
| Position | R4.9 A16.5 H29.7 mm |
| Orientation | T > C-4.3 > S3.3 |
| Phase Encoding Dir. | A >> P |
| Phase Oversampling | 0 % |
| FoV Read | 208 mm |
| FoV Phase | 100.0 % |
| Slice Thickness | 2.0 mm |
| TR | 8000.0 ms |
| Multi-Slice Mode | Interleaved |
| Series | Interleaved |
| Multi-band accel. factor | 1 |

### Geometry - AutoAlign

| | |
|---|---|
| Slice Group | 1 |
| Position | R4.9 A16.5 H29.7 mm |
| Orientation | T > C-4.3 > S3.3 |
| Phase Encoding Dir. | A >> P |
| AutoAlign | Head > Brain |
| Initial Position | R1.4 A2.6 H14.8 |
| R | 1.4 mm |
| A | 2.6 mm |
| H | 14.8 mm |
| Initial Orientation | T > S |
| T > S | -0.40 |
| > C | -0.10 |
| Initial Rotation | 0.14 deg |

### Geometry - Saturation

| | |
|---|---|
| Special Saturation | None |

### Geometry - Tim Planning Suite

| | |
|---|---|
| Set-n-Go Protocol | Off |
| Table Position | 0 mm |
| Table Position | H |
| Inline Composing | Off |

### System - Miscellaneous

| | |
|---|---|
| Coil Selection | ACS All but spine |
| MSMA | S - C - T |
| Sagittal | R >> L |
| Coronal | A >> P |
| Transversal | F >> H |
| Coil Combination | Sum of Squares |
| Matrix Optimization | Off |
| Coil Focus | Flat |

### System - Adjustments

| | |
|---|---|
| Adjustment Strategy | Standard |
| B0 Shim | Standard |
| B1 Shim | TrueForm |
| CoilShim | Off |
| Adjustment Tolerance | Auto |
| Adjust with Body Coil | Off |
| Confirm Frequency | Never |
| Assume Silicone | Off |

### System - Adjust Volume

| | |
|---|---|
| Position | R4.9 A16.5 H29.7 mm |
| Orientation | T > C-4.3 > S3.3 |
| Rotation | -0.44 deg |
| A >> P | 208 mm |
| R >> L | 208 mm |
| F >> H | 144 mm |
| Reset | Off |

### System - pTx

| | |
|---|---|
| B1 Shim | TrueForm |
| Excitation | Standard |

### System - Tx/Rx

| | |
|---|---|
| Frequency 1H | 123.257916 MHz |
| ? Ref. Amplitude 1H | 0.000 V |
| Reset | Off |
| Image Scaling | 1.000 |

### Physio - Signal

| | |
|---|---|
| 1st Signal/Mode | None |
| TR | 8000.0 ms |
| Multi-band accel. factor | 1 |

### BOLD

| | |
|---|---|
| GLM Statistics | Off |
| Ignore Meas. at Start | 0 |
| Ignore After Transition | 0 |
| Model Transition States | Off |
| Temp. Highpass Filter | Off |
| Threshold | 4.00 |
| Paradigm Size | 3 |
| Meas[1] | Active |
| Meas[2] | Active |
| Meas[3] | Ignore |
| Motion Correction | Off |
| Spatial Filter | Off |
| Measurements | 1 |
| Delay in TR | 0.00 ms |

### Sequence - Part 1

| | |
|---|---|
| Sequence Name | epse |
| Dimension | 2D |
| Excitation | Standard |
| RF Pulse Type | Normal |
| Gradient Mode | Performance |
| Bandwidth | 2290 Hz/Px |
| Echo Spacing | 0.58 ms |
| Free Echo Spacing | On |
| EPI Factor | 104 |

### Sequence - Part 2

| | |
|---|---|
| Introduction | Off |

### Sequence - Special

| | |
|---|---|
| Min. prep scans | 0 |
| Delay before PC scans | 0 us |
| SENSE1 coil combine | Off |
| Invert RO/PE polarity | Off |
| Disable freq. update | Off |
| Suppress 16-bit DICOM | Off |
| Force equal slice timing | Off |
| FFT scale factor | 1.00 |
| Fat saturation FA | 110.00 deg |
| Fat sat. offset | 0.00 Hz |
| Physio recording | Off |
| Triggering scheme | Standard |

### Sequence - Assistant

| | |
|---|---|
| SAR Assistant | Off |

**\\PHYSICS\XA30 RESEARCH PROT\PARCELLATION\BRAINGATE2 08-26-2024\SpinEchoFieldMap_2_PA**

TA: 8 sec Coil Selection: Auto Voxel Size: 2.0×2.0×2.0 mm³ Acc:: None Rel. SNR: 1.00

### Properties

| | |
|---|---|
| Start measurement without further preparation | On |
| Wait for User to Start | Off |
| Start measurements | Single Measurement |
| Prio Recon | Off |
| Auto Open Inline Display | Off |
| Auto Close Inline Display | Off |
| Load Images to MR View&GO | On |
| Auto Store Images | On |
| Load Images to Stamp Segments | Off |
| Load Images to Graphic Segments | Off |
| Graphic segment | Default |
| Inline Movie | Off |

### Routine

| | |
|---|---|
| Slice Group | 1 |
| Slices | 72 |
| Distance Factor | 0 % |
| Position | R4.9 A16.5 H29.7 mm |
| Orientation | T > C-4.3 > S3.3 |
| Phase Encoding Dir. | P >> A |
| Phase Oversampling | 0 % |
| FoV Read | 208 mm |
| FoV Phase | 100.0 % |
| Slice Thickness | 2.0 mm |
| TR | 8000.0 ms |
| TE | 66.00 ms |
| Averages | 1 |
| Multi-band accel. factor | 1 |
| AutoAlign | Head > Brain |

### Contrast - Common

| | |
|---|---|
| TR | 8000.0 ms |
| TE | 66.00 ms |
| MTC | Off |
| Magn. Preparation | None |
| Flip Angle | 90 deg |
| Refocus flip angle | 180 deg |
| Fat-Water Contrast | Fat Saturation |
| Grad. rev. fat suppr. | Disabled |
| Contrasts | 1 |
| Reconstruction | Magnitude |

### Contrast - Dynamic

| | |
|---|---|
| Dynamic Mode | Standard |
| Measurements | 1 |
| Delay in TR | 0.00 ms |

### Resolution - Common

| | |
|---|---|
| FoV Read | 208 mm |
| FoV Phase | 100.0 % |
| Slice Thickness | 2.0 mm |
| Base Resolution | 104 |
| Phase Resolution | 100 % |
| Interpolation | Off |

### Resolution - Acceleration

| | |
|---|---|
| Acceleration mode | None |
| Advanced Reconstruction | Off |

### Resolution - Acceleration

| | |
|---|---|
| Phase Partial Fourier | Off |

### Resolution - Filter

| | |
|---|---|
| Raw Filter | Off |
| Elliptical Filter | Off |
| Hamming | Off |
| Distortion Correction | Off |
| Normalize | Off |

### Geometry - Common

| | |
|---|---|
| Slice Group | 1 |
| Slices | 72 |
| Distance Factor | 0 % |
| Position | R4.9 A16.5 H29.7 mm |
| Orientation | T > C-4.3 > S3.3 |
| Phase Encoding Dir. | P >> A |
| Phase Oversampling | 0 % |
| FoV Read | 208 mm |
| FoV Phase | 100.0 % |
| Slice Thickness | 2.0 mm |
| TR | 8000.0 ms |
| Multi-Slice Mode | Interleaved |
| Series | Interleaved |
| Multi-band accel. factor | 1 |

### Geometry - AutoAlign

| | |
|---|---|
| Slice Group | 1 |
| Position | R4.9 A16.5 H29.7 mm |
| Orientation | T > C-4.3 > S3.3 |
| Phase Encoding Dir. | P >> A |
| AutoAlign | Head > Brain |
| Initial Position | R1.4 A2.6 H14.8 |
| R | 1.4 mm |
| A | 2.6 mm |
| H | 14.8 mm |
| Initial Orientation | T > S |
| T > S | -0.40 |
| > C | -0.10 |
| Initial Rotation | -180.00 deg |

### Geometry - Saturation

| | |
|---|---|
| Special Saturation | None |

### Geometry - Tim Planning Suite

| | |
|---|---|
| Set-n-Go Protocol | Off |
| Table Position | 0 mm |
| Table Position | H |
| Inline Composing | Off |

### System - Miscellaneous

| | |
|---|---|
| Coil Selection | ACS All but spine |
| MSMA | S - C - T |
| Sagittal | R >> L |
| Coronal | A >> P |
| Transversal | F >> H |
| Coil Combination | Sum of Squares |
| Matrix Optimization | Off |
| Coil Focus | Flat |

### System - Adjustments

| | |
|---|---|
| Adjustment Strategy | Standard |
| B0 Shim | Standard |
| B1 Shim | TrueForm |
| CoilShim | Off |
| Adjustment Tolerance | Auto |
| Adjust with Body Coil | Off |
| Confirm Frequency | Never |
| Assume Silicone | Off |

### System - Adjust Volume

| | |
|---|---|
| Position | R4.9 A16.5 H29.7 mm |
| Orientation | T > C-4.3 > S3.3 |
| Rotation | 179.42 deg |
| A >> P | 208 mm |
| R >> L | 208 mm |
| F >> H | 144 mm |
| Reset | Off |

### System - pTx

| | |
|---|---|
| B1 Shim | TrueForm |
| Excitation | Standard |

### System - Tx/Rx

| | |
|---|---|
| Frequency 1H | 123.257916 MHz |
| ? Ref. Amplitude 1H | 0.000 V |
| Reset | Off |
| Image Scaling | 1.000 |

### Physio - Signal

| | |
|---|---|
| 1st Signal/Mode | None |
| TR | 8000.0 ms |
| Multi-band accel. factor | 1 |

### BOLD

| | |
|---|---|
| GLM Statistics | Off |
| Ignore Meas. at Start | 0 |
| Ignore After Transition | 0 |
| Model Transition States | Off |
| Temp. Highpass Filter | Off |
| Threshold | 4.00 |
| Paradigm Size | 3 |
| Meas[1] | Active |
| Meas[2] | Active |
| Meas[3] | Ignore |
| Motion Correction | Off |
| Spatial Filter | Off |
| Measurements | 1 |
| Delay in TR | 0.00 ms |

### Sequence - Part 1

| | |
|---|---|
| Sequence Name | epse |
| Dimension | 2D |
| Excitation | Standard |
| RF Pulse Type | Normal |
| Gradient Mode | Performance |
| Bandwidth | 2290 Hz/Px |
| Echo Spacing | 0.58 ms |
| Free Echo Spacing | On |
| EPI Factor | 104 |

### Sequence - Part 2

| | |
|---|---|
| Introduction | Off |

### Sequence - Special

| | |
|---|---|
| Min. prep scans | 0 |
| Delay before PC scans | 0 us |
| SENSE1 coil combine | Off |
| Invert RO/PE polarity | On |
| Disable freq. update | Off |
| Suppress 16-bit DICOM | Off |
| Force equal slice timing | Off |
| FFT scale factor | 1.00 |
| Fat saturation FA | 110.00 deg |
| Fat sat. offset | 0.00 Hz |
| Physio recording | Off |
| Triggering scheme | Standard |

### Sequence - Assistant

| | |
|---|---|
| SAR Assistant | Off |

**\\PHYSICS\XA30 RESEARCH PROT\PARCELLATION\BRAINGATE2 08-26-2024\rfMRI_REST_2_AP**

TA: 6:40 min Coil Selection: Auto Voxel Size: 2.0×2.0×2.0 mm³ Acc:: None Rel. SNR: 1.00

### Properties

| | |
|---|---|
| Start measurement without further preparation | On |
| Wait for User to Start | Off |
| Start measurements | Single Measurement |
| Prio Recon | Off |
| Auto Open Inline Display | Off |
| Auto Close Inline Display | Off |
| Load Images to MR View&GO | On |
| Auto Store Images | On |
| Load Images to Stamp Segments | Off |
| Load Images to Graphic Segments | Off |
| Graphic segment | Default |
| Inline Movie | Off |

### Routine

| | |
|---|---|
| Slice Group | 1 |
| Slices | 72 |
| Distance Factor | 0 % |
| Position | R4.9 A16.5 H29.7 mm |
| Orientation | T > C-4.3 > S3.3 |
| Phase Encoding Dir. | A >> P |
| Phase Oversampling | 0 % |
| FoV Read | 208 mm |
| FoV Phase | 100.0 % |
| Slice Thickness | 2.0 mm |
| TR | 800.0 ms |
| TE | 37.00 ms |
| Averages | 1 |
| Multi-band accel. factor | 8 |
| AutoAlign | Head > Brain |

### Contrast - Common

| | |
|---|---|
| TR | 800.0 ms |
| TE | 37.00 ms |
| MTC | Off |
| Magn. Preparation | None |
| Flip Angle | 52 deg |
| Fat-Water Contrast | Fat Saturation |
| Contrasts | 1 |
| Reconstruction | Magnitude |

### Contrast - Dynamic

| | |
|---|---|
| Dynamic Mode | Standard |
| Measurements | 488 |
| Delay in TR | 0.00 ms |

### Resolution - Common

| | |
|---|---|
| FoV Read | 208 mm |
| FoV Phase | 100.0 % |
| Slice Thickness | 2.0 mm |
| Base Resolution | 104 |
| Phase Resolution | 100 % |
| Interpolation | Off |

### Resolution - Acceleration

| | |
|---|---|
| Acceleration mode | None |
| Advanced Reconstruction | Off |
| Phase Partial Fourier | Off |

### Resolution - Filter

| | |
|---|---|
| Raw Filter | Off |
| Elliptical Filter | Off |
| Hamming | Off |
| Distortion Correction | 2D |
| Normalize | Off |

### Geometry - Common

| | |
|---|---|
| Slice Group | 1 |
| Slices | 72 |
| Distance Factor | 0 % |
| Position | R4.9 A16.5 H29.7 mm |
| Orientation | T > C-4.3 > S3.3 |
| Phase Encoding Dir. | A >> P |
| Phase Oversampling | 0 % |
| FoV Read | 208 mm |
| FoV Phase | 100.0 % |
| Slice Thickness | 2.0 mm |
| TR | 800.0 ms |
| Multi-Slice Mode | Interleaved |
| Series | Interleaved |
| Multi-band accel. factor | 8 |

### Geometry - AutoAlign

| | |
|---|---|
| Slice Group | 1 |
| Position | R4.9 A16.5 H29.7 mm |
| Orientation | T > C-4.3 > S3.3 |
| Phase Encoding Dir. | A >> P |
| AutoAlign | Head > Brain |
| Initial Position | R1.4 A2.6 H14.8 |
| R | 1.4 mm |
| A | 2.6 mm |
| H | 14.8 mm |
| Initial Orientation | T > S |
| T > S | -0.40 |
| > C | -0.10 |
| Initial Rotation | 0.14 deg |

### Geometry - Saturation

| | |
|---|---|
| Special Saturation | None |

### Geometry - Tim Planning Suite

| | |
|---|---|
| Set-n-Go Protocol | Off |
| Table Position | 0 mm |
| Table Position | H |
| Inline Composing | Off |

### System - Miscellaneous

| | |
|---|---|
| Coil Selection | Auto Coil Select |
| MSMA | S - C - T |
| Sagittal | R >> L |
| Coronal | A >> P |
| Transversal | F >> H |
| Coil Combination | Sum of Squares |
| Matrix Optimization | Off |
| Coil Focus | Flat |

### System - Adjustments

| | |
|---|---|
| Adjustment Strategy | Standard |
| B0 Shim | Standard |
| B1 Shim | TrueForm |

### System - Adjustments

| | |
|---|---|
| CoilShim | Off |
| Adjustment Tolerance | Auto |
| Adjust with Body Coil | Off |
| Confirm Frequency | Never |
| Assume Silicone | Off |

### System - Adjust Volume

| | |
|---|---|
| Position | R4.9 A16.5 H29.7 mm |
| Orientation | T > C-4.3 > S3.3 |
| Rotation | -0.44 deg |
| A >> P | 208 mm |
| R >> L | 208 mm |
| F >> H | 144 mm |
| Reset | Off |

### System - pTx

| | |
|---|---|
| B1 Shim | TrueForm |
| Excitation | Standard |

### System - Tx/Rx

| | |
|---|---|
| Frequency 1H | 123.257916 MHz |
| ? Ref. Amplitude 1H | 0.000 V |
| Reset | Off |
| Image Scaling | 1.000 |

### Physio - Signal

| | |
|---|---|
| 1st Signal/Mode | None |
| TR | 800.0 ms |
| Multi-band accel. factor | 8 |

### BOLD

| | |
|---|---|
| GLM Statistics | Off |
| Ignore Meas. at Start | 0 |
| Ignore After Transition | 0 |
| Model Transition States | On |
| Temp. Highpass Filter | On |
| Threshold | 4.00 |
| Paradigm Size | 3 |
| Meas[1] | Active |
| Meas[2] | Active |
| Meas[3] | Ignore |
| Motion Correction | Off |
| Spatial Filter | Off |
| Measurements | 488 |
| Delay in TR | 0.00 ms |

### Sequence - Part 1

| | |
|---|---|
| Sequence Name | epfid |
| Dimension | 2D |
| Excitation | Standard |
| Gradient Mode | Performance |
| Flow Compensation | None |
| Bandwidth | 2290 Hz/Px |
| Echo Spacing | 0.58 ms |
| Free Echo Spacing | On |
| EPI Factor | 104 |

### Sequence - Part 2

| | |
|---|---|
| Introduction | Off |
| RF Spoiling | Off |

### Sequence - Special

| | |
|---|---|
| Excite pulse duration | 6600 us |

### Sequence - Special

| | |
|---|---|
| Min. prep scans | 0 |
| Min. prep scans SB | 0 |
| Delay before PC scans | 0 us |
| Single-band images | Off |
| MB LeakBlock kernel | On |
| MB dual kernel | Off |
| MB RF phase scramble | Off |
| Opt. MB RF pulse BW | Off |
| SENSE1 coil combine | Off |
| Invert RO/PE polarity | Off |
| Disable freq. update | Off |
| Suppress 16-bit DICOM | Off |
| Force equal slice timing | Off |
| Online multi-band recon. | Online |
| FFT scale factor | 1.00 |
| Fat saturation FA | 110.00 deg |
| Fat sat. offset | 0.00 Hz |
| Sinc exc. pulse BWTP | 5.20 |
| Physio recording | Off |
| Triggering scheme | Standard |

### Sequence - Assistant

| | |
|---|---|
| SAR Assistant | Off |

| \\PHYSICS\XA30 RESEARCH PROT\PARCELLATION\BRAINGATE2 08-26-2024\rfMRI_REST_2_PA |
| --- |
| TA: 6:40 min Coil Selection: Auto Voxel Size: 2.0×2.0×2.0 mm³ Acc:: None Rel. SNR: 1.00 |

### Properties

| | |
|---|---|
| Start measurement without further preparation | On |
| Wait for User to Start | Off |
| Start measurements | Single Measurement |
| Prio Recon | Off |
| Auto Open Inline Display | Off |
| Auto Close Inline Display | Off |
| Load Images to MR View&GO | On |
| Auto Store Images | On |
| Load Images to Stamp Segments | Off |
| Load Images to Graphic Segments | Off |
| Graphic segment | Default |
| Inline Movie | Off |

### Routine

| | |
|---|---|
| Slice Group | 1 |
| Slices | 72 |
| Distance Factor | 0 % |
| Position | R4.9 A16.5 H29.7 mm |
| Orientation | T > C-4.3 > S3.3 |
| Phase Encoding Dir. | P >> A |
| Phase Oversampling | 0 % |
| FoV Read | 208 mm |
| FoV Phase | 100.0 % |
| Slice Thickness | 2.0 mm |
| TR | 800.0 ms |
| TE | 37.00 ms |
| Averages | 1 |
| Multi-band accel. factor | 8 |
| AutoAlign | Head > Brain |

### Contrast - Common

| | |
|---|---|
| TR | 800.0 ms |
| TE | 37.00 ms |
| MTC | Off |
| Magn. Preparation | None |
| Flip Angle | 52 deg |
| Fat-Water Contrast | Fat Saturation |
| Contrasts | 1 |
| Reconstruction | Magnitude |

### Contrast - Dynamic

| | |
|---|---|
| Dynamic Mode | Standard |
| Measurements | 488 |
| Delay in TR | 0.00 ms |

### Resolution - Common

| | |
|---|---|
| FoV Read | 208 mm |
| FoV Phase | 100.0 % |
| Slice Thickness | 2.0 mm |
| Base Resolution | 104 |
| Phase Resolution | 100 % |
| Interpolation | Off |

### Resolution - Acceleration

| | |
|---|---|
| Acceleration mode | None |
| Advanced Reconstruction | Off |
| Phase Partial Fourier | Off |

### Resolution - Filter

| | |
|---|---|
| Raw Filter | Off |
| Elliptical Filter | Off |
| Hamming | Off |
| Distortion Correction | 2D |
| Normalize | Off |

### Geometry - Common

| | |
|---|---|
| Slice Group | 1 |
| Slices | 72 |
| Distance Factor | 0 % |
| Position | R4.9 A16.5 H29.7 mm |
| Orientation | T > C-4.3 > S3.3 |
| Phase Encoding Dir. | P >> A |
| Phase Oversampling | 0 % |
| FoV Read | 208 mm |
| FoV Phase | 100.0 % |
| Slice Thickness | 2.0 mm |
| TR | 800.0 ms |
| Multi-Slice Mode | Interleaved |
| Series | Interleaved |
| Multi-band accel. factor | 8 |

### Geometry - AutoAlign

| | |
|---|---|
| Slice Group | 1 |
| Position | R4.9 A16.5 H29.7 mm |
| Orientation | T > C-4.3 > S3.3 |
| Phase Encoding Dir. | P >> A |
| AutoAlign | Head > Brain |
| Initial Position | R1.4 A2.6 H14.8 |
| R | 1.4 mm |
| A | 2.6 mm |
| H | 14.8 mm |
| Initial Orientation | T > S |
| T > S | -0.40 |
| > C | -0.10 |
| Initial Rotation | -180.00 deg |

### Geometry - Saturation

| | |
|---|---|
| Special Saturation | None |

### Geometry - Tim Planning Suite

| | |
|---|---|
| Set-n-Go Protocol | Off |
| Table Position | 0 mm |
| Table Position | H |
| Inline Composing | Off |

### System - Miscellaneous

| | |
|---|---|
| Coil Selection | Auto Coil Select |
| MSMA | S - C - T |
| Sagittal | R >> L |
| Coronal | A >> P |
| Transversal | F >> H |
| Coil Combination | Sum of Squares |
| Matrix Optimization | Off |
| Coil Focus | Flat |

### System - Adjustments

| | |
|---|---|
| Adjustment Strategy | Standard |
| B0 Shim | Standard |
| B1 Shim | TrueForm |

### System - Adjustments

| | |
|---|---|
| CoilShim | Off |
| Adjustment Tolerance | Auto |
| Adjust with Body Coil | Off |
| Confirm Frequency | Never |
| Assume Silicone | Off |

### System - Adjust Volume

| | |
|---|---|
| Position | R4.9 A16.5 H29.7 mm |
| Orientation | T > C-4.3 > S3.3 |
| Rotation | 179.42 deg |
| A >> P | 208 mm |
| R >> L | 208 mm |
| F >> H | 144 mm |
| Reset | Off |

### System - pTx

| | |
|---|---|
| B1 Shim | TrueForm |
| Excitation | Standard |

### System - Tx/Rx

| | |
|---|---|
| Frequency 1H | 123.257916 MHz |
| ? Ref. Amplitude 1H | 0.000 V |
| Reset | Off |
| Image Scaling | 1.000 |

### Physio - Signal

| | |
|---|---|
| 1st Signal/Mode | None |
| TR | 800.0 ms |
| Multi-band accel. factor | 8 |

### BOLD

| | |
|---|---|
| GLM Statistics | Off |
| Ignore Meas. at Start | 0 |
| Ignore After Transition | 0 |
| Model Transition States | On |
| Temp. Highpass Filter | On |
| Threshold | 4.00 |
| Paradigm Size | 3 |
| Meas[1] | Active |
| Meas[2] | Active |
| Meas[3] | Ignore |
| Motion Correction | Off |
| Spatial Filter | Off |
| Measurements | 488 |
| Delay in TR | 0.00 ms |

### Sequence - Part 1

| | |
|---|---|
| Sequence Name | epfid |
| Dimension | 2D |
| Excitation | Standard |
| Gradient Mode | Performance |
| Flow Compensation | None |
| Bandwidth | 2290 Hz/Px |
| Echo Spacing | 0.58 ms |
| Free Echo Spacing | On |
| EPI Factor | 104 |

### Sequence - Part 2

| | |
|---|---|
| Introduction | Off |
| RF Spoiling | Off |

### Sequence - Special

| | |
|---|---|
| Excite pulse duration | 6600 us |
| Min. prep scans | 0 |
| Min. prep scans SB | 0 |
| Delay before PC scans | 0 us |
| Single-band images | Off |
| MB LeakBlock kernel | On |
| MB dual kernel | Off |
| MB RF phase scramble | Off |
| Opt. MB RF pulse BW | Off |
| SENSE1 coil combine | Off |
| Invert RO/PE polarity | Off |
| Disable freq. update | Off |
| Suppress 16-bit DICOM | Off |
| Force equal slice timing | Off |
| Online multi-band recon. | Online |
| FFT scale factor | 1.00 |
| Fat saturation FA | 110.00 deg |
| Fat sat. offset | 0.00 Hz |
| Sinc exc. pulse BWTP | 5.20 |
| Physio recording | Off |
| Triggering scheme | Standard |

### Sequence - Assistant

| | |
|---|---|
| SAR Assistant | Off |

**\\PHYSICS\XA30 RESEARCH PROT\PARCELLATION\BRAINGATE2 08-26-2024\OPEN-CLOSE RT HAND_FMRI**

TA: 4:39 min Coil Selection: Auto Voxel Size: 3.0×3.0×3.0 mm³ Acc:: 2 Rel. SNR: 1.00

### Properties

| | |
|---|---|
| Start measurement without further preparation | On |
| Wait for User to Start | On |
| Start measurements | Single Measurement |
| Prio Recon | Off |
| Auto Open Inline Display | On |
| Auto Close Inline Display | Off |
| Load Images to MR View&GO | On |
| Auto Store Images | On |
| Load Images to Stamp Segments | Off |
| Load Images to Graphic Segments | Off |
| Graphic segment | Default |
| Inline Movie | Off |

### Routine

| | |
|---|---|
| Slice Group | 1 |
| Slices | 41 |
| Distance Factor | 0 % |
| Position | R4.9 A16.5 H29.7 mm |
| Orientation | T > C-4.3 > S3.3 |
| Phase Encoding Dir. | A >> P |
| Phase Oversampling | 0 % |
| FoV Read | 192 mm |
| FoV Phase | 100.0 % |
| Slice Thickness | 3.0 mm |
| TR | 3000.0 ms |
| TE | 30.00 ms |
| Averages | 1 |
| Concatenations | 1 |
| AutoAlign | Head > Brain |

### Contrast - Common

| | |
|---|---|
| TR | 3000.0 ms |
| TE | 30.00 ms |
| MTC | Off |
| Flip Angle | 90 deg |
| Fat-Water Contrast | Fat Saturation |
| Reconstruction | Magnitude |

### Contrast - Dynamic

| | |
|---|---|
| Dynamic Mode | Standard |
| Measurements | 90 |
| Delay in TR | 790.00 ms |

### Resolution - Common

| | |
|---|---|
| FoV Read | 192 mm |
| FoV Phase | 100.0 % |
| Slice Thickness | 3.0 mm |
| Base Resolution | 64 |
| Phase Resolution | 100 % |
| Interpolation | Off |

### Resolution - Acceleration

| | |
|---|---|
| Acceleration mode | GRAPPA |
| Reference Scans | EPI/Separate |
| Acceleration Factor PE | 2 |
| Reference Lines PE | 24 |
| Advanced Reconstruction | Off |
| Phase Partial Fourier | Off |

### Resolution - Filter

| | |
|---|---|
| Raw Filter | Off |
| Elliptical Filter | Off |
| Hamming | Off |
| Distortion Correction | Off |
| Normalize | Prescan |

### Geometry - Common

| | |
|---|---|
| Slice Group | 1 |
| Slices | 41 |
| Distance Factor | 0 % |
| Position | R4.9 A16.5 H29.7 mm |
| Orientation | T > C-4.3 > S3.3 |
| Phase Encoding Dir. | A >> P |
| Phase Oversampling | 0 % |
| FoV Read | 192 mm |
| FoV Phase | 100.0 % |
| Slice Thickness | 3.0 mm |
| TR | 3000.0 ms |
| Multi-Slice Mode | Interleaved |
| Series | Interleaved |
| Concatenations | 1 |

### Geometry - AutoAlign

| | |
|---|---|
| Slice Group | 1 |
| Position | R4.9 A16.5 H29.7 mm |
| Orientation | T > C-4.3 > S3.3 |
| Phase Encoding Dir. | A >> P |
| AutoAlign | Head > Brain |
| Initial Position | R1.4 A2.6 H14.8 |
| R | 1.4 mm |
| A | 2.6 mm |
| H | 14.8 mm |
| Initial Orientation | T > S |
| T > S | -0.40 |
| > C | -0.10 |
| Initial Rotation | 0.14 deg |

### Geometry - Saturation

| | |
|---|---|
| Special Saturation | None |

### Geometry - Tim Planning Suite

| | |
|---|---|
| Set-n-Go Protocol | Off |
| Table Position | 0 mm |
| Table Position | H |
| Inline Composing | Off |

### System - Miscellaneous

| | |
|---|---|
| Coil Selection | ACS All but spine |
| MSMA | S - C - T |
| Sagittal | R >> L |
| Coronal | A >> P |
| Transversal | F >> H |
| Coil Combination | Sum of Squares |
| Matrix Optimization | Off |
| Coil Focus | Flat |

### System - Adjustments

| | |
|---|---|
| Adjustment Strategy | Standard |
| B0 Shim | Standard |

### System - Adjustments

| | |
|---|---|
| B1 Shim | TrueForm |
| CoilShim | Off |
| Adjustment Tolerance | Auto |
| Adjust with Body Coil | Off |
| Confirm Frequency | Never |
| Assume Silicone | Off |

### System - Adjust Volume

| | |
|---|---|
| Position | R4.9 A16.5 H29.7 mm |
| Orientation | T > C-4.3 > S3.3 |
| Rotation | -0.44 deg |
| A >> P | 192 mm |
| R >> L | 192 mm |
| F >> H | 123 mm |
| Reset | Off |

### System - pTx

| | |
|---|---|
| B1 Shim | TrueForm |
| Excitation | Standard |

### System - Tx/Rx

| | |
|---|---|
| Frequency 1H | 123.257916 MHz |
| ? Ref. Amplitude 1H | 0.000 V |
| Reset | Off |
| Image Scaling | 1.000 |

### Physio - Signal

| | |
|---|---|
| 1st Signal/Mode | None |
| TR | 3000.0 ms |
| Concatenations | 1 |

### BOLD

| | |
|---|---|
| GLM Statistics | On |
| Ignore Meas. at Start | 0 |
| Ignore After Transition | 0 |
| Model Transition States | On |
| Temp. Highpass Filter | On |
| Threshold | 4.00 |
| Paradigm Size | 20 |
| Meas[1] | Active |
| Meas[2] | Active |
| Meas[3] | Active |
| Meas[4] | Active |
| Meas[5] | Active |
| Meas[6] | Active |
| Meas[7] | Active |
| Meas[8] | Active |
| Meas[9] | Active |
| Meas[10] | Active |
| Meas[11] | Ignore |
| Meas[12] | Ignore |
| Meas[13] | Ignore |
| Meas[14] | Ignore |
| Meas[15] | Ignore |
| Meas[16] | Ignore |
| Meas[17] | Ignore |
| Meas[18] | Ignore |
| Meas[19] | Ignore |
| Meas[20] | Ignore |
| Motion Correction | Off |
| Spatial Filter | Off |
| Measurements | 90 |
| Delay in TR | 790.00 ms |

### Sequence - Part 1

| | |
|---|---|
| Sequence Name | epfid |
| Excitation | Standard |
| RF Pulse Type | Normal |
| Gradient Mode | Fast |
| Bandwidth | 2442 Hz/Px |
| Echo Spacing | 0.49 ms |
| Free Echo Spacing | Off |
| EPI Factor | 64 |

### Sequence - Part 2

| | |
|---|---|
| Introduction | Off |

**\\PHYSICS\XA30 RESEARCH PROT\PARCELLATION\BRAINGATE2 08-26-2024\OPEN-CLOSE LT HAND_FMRI**

TA: 4:39 min Coil Selection: Auto Voxel Size: 3.0×3.0×3.0 mm³ Acc:: 2 Rel. SNR: 1.00

### Properties

| | |
|---|---|
| Start measurement without further preparation | On |
| Wait for User to Start | On |
| Start measurements | Single Measurement |
| Prio Recon | Off |
| Auto Open Inline Display | On |
| Auto Close Inline Display | Off |
| Load Images to MR View&GO | On |
| Auto Store Images | On |
| Load Images to Stamp Segments | Off |
| Load Images to Graphic Segments | Off |
| Graphic segment | Default |
| Inline Movie | Off |

### Routine

| | |
|---|---|
| Slice Group | 1 |
| Slices | 41 |
| Distance Factor | 0 % |
| Position | R4.9 A16.5 H29.7 mm |
| Orientation | T > C-4.3 > S3.3 |
| Phase Encoding Dir. | A >> P |
| Phase Oversampling | 0 % |
| FoV Read | 192 mm |
| FoV Phase | 100.0 % |
| Slice Thickness | 3.0 mm |
| TR | 3000.0 ms |
| TE | 30.00 ms |
| Averages | 1 |
| Concatenations | 1 |
| AutoAlign | Head > Brain |

### Contrast - Common

| | |
|---|---|
| TR | 3000.0 ms |
| TE | 30.00 ms |
| MTC | Off |
| Flip Angle | 90 deg |
| Fat-Water Contrast | Fat Saturation |
| Reconstruction | Magnitude |

### Contrast - Dynamic

| | |
|---|---|
| Dynamic Mode | Standard |
| Measurements | 90 |
| Delay in TR | 790.00 ms |

### Resolution - Common

| | |
|---|---|
| FoV Read | 192 mm |
| FoV Phase | 100.0 % |
| Slice Thickness | 3.0 mm |
| Base Resolution | 64 |
| Phase Resolution | 100 % |
| Interpolation | Off |

### Resolution - Acceleration

| | |
|---|---|
| Acceleration mode | GRAPPA |
| Reference Scans | EPI/Separate |
| Acceleration Factor PE | 2 |
| Reference Lines PE | 24 |
| Advanced Reconstruction | Off |
| Phase Partial Fourier | Off |

### Resolution - Filter

| | |
|---|---|
| Raw Filter | Off |
| Elliptical Filter | Off |
| Hamming | Off |
| Distortion Correction | Off |
| Normalize | Prescan |

### Geometry - Common

| | |
|---|---|
| Slice Group | 1 |
| Slices | 41 |
| Distance Factor | 0 % |
| Position | R4.9 A16.5 H29.7 mm |
| Orientation | T > C-4.3 > S3.3 |
| Phase Encoding Dir. | A >> P |
| Phase Oversampling | 0 % |
| FoV Read | 192 mm |
| FoV Phase | 100.0 % |
| Slice Thickness | 3.0 mm |
| TR | 3000.0 ms |
| Multi-Slice Mode | Interleaved |
| Series | Interleaved |
| Concatenations | 1 |

### Geometry - AutoAlign

| | |
|---|---|
| Slice Group | 1 |
| Position | R4.9 A16.5 H29.7 mm |
| Orientation | T > C-4.3 > S3.3 |
| Phase Encoding Dir. | A >> P |
| AutoAlign | Head > Brain |
| Initial Position | R1.4 A2.6 H14.8 |
| R | 1.4 mm |
| A | 2.6 mm |
| H | 14.8 mm |
| Initial Orientation | T > S |
| T > S | -0.40 |
| > C | -0.10 |
| Initial Rotation | 0.14 deg |

### Geometry - Saturation

| | |
|---|---|
| Special Saturation | None |

### Geometry - Tim Planning Suite

| | |
|---|---|
| Set-n-Go Protocol | Off |
| Table Position | 0 mm |
| Table Position | H |
| Inline Composing | Off |

### System - Miscellaneous

| | |
|---|---|
| Coil Selection | ACS All but spine |
| MSMA | S - C - T |
| Sagittal | R >> L |
| Coronal | A >> P |
| Transversal | F >> H |
| Coil Combination | Sum of Squares |
| Matrix Optimization | Off |
| Coil Focus | Flat |

### System - Adjustments

| | |
|---|---|
| Adjustment Strategy | Standard |
| B0 Shim | Standard |

### System - Adjustments

| | |
|---|---|
| B1 Shim | TrueForm |
| CoilShim | Off |
| Adjustment Tolerance | Auto |
| Adjust with Body Coil | Off |
| Confirm Frequency | Never |
| Assume Silicone | Off |

### System - Adjust Volume

| | |
|---|---|
| Position | R4.9 A16.5 H29.7 mm |
| Orientation | T > C-4.3 > S3.3 |
| Rotation | -0.44 deg |
| A >> P | 192 mm |
| R >> L | 192 mm |
| F >> H | 123 mm |
| Reset | Off |

### System - pTx

| | |
|---|---|
| B1 Shim | TrueForm |
| Excitation | Standard |

### System - Tx/Rx

| | |
|---|---|
| Frequency 1H | 123.257916 MHz |
| ? Ref. Amplitude 1H | 0.000 V |
| Reset | Off |
| Image Scaling | 1.000 |

### Physio - Signal

| | |
|---|---|
| 1st Signal/Mode | None |
| TR | 3000.0 ms |
| Concatenations | 1 |

### BOLD

| | |
|---|---|
| GLM Statistics | On |
| Ignore Meas. at Start | 0 |
| Ignore After Transition | 0 |
| Model Transition States | On |
| Temp. Highpass Filter | On |
| Threshold | 4.00 |
| Paradigm Size | 20 |
| Meas[1] | Active |
| Meas[2] | Active |
| Meas[3] | Active |
| Meas[4] | Active |
| Meas[5] | Active |
| Meas[6] | Active |
| Meas[7] | Active |
| Meas[8] | Active |
| Meas[9] | Active |
| Meas[10] | Active |
| Meas[11] | Ignore |
| Meas[12] | Ignore |
| Meas[13] | Ignore |
| Meas[14] | Ignore |
| Meas[15] | Ignore |
| Meas[16] | Ignore |
| Meas[17] | Ignore |
| Meas[18] | Ignore |
| Meas[19] | Ignore |
| Meas[20] | Ignore |
| Motion Correction | Off |
| Spatial Filter | Off |
| Measurements | 90 |
| Delay in TR | 790.00 ms |

### Sequence - Part 1

| | |
|---|---|
| Sequence Name | epfid |
| Excitation | Standard |
| RF Pulse Type | Normal |
| Gradient Mode | Fast |
| Bandwidth | 2442 Hz/Px |
| Echo Spacing | 0.49 ms |
| Free Echo Spacing | Off |
| EPI Factor | 64 |

### Sequence - Part 2

| | |
|---|---|
| Introduction | Off |

| \\PHYSICS\XA30 RESEARCH PROT\PARCELLATION\BRAINGATE2 08-26-2024\LIPS_FMRI |
|---|
| TA: 4:39 min Coil Selection: Auto Voxel Size: 3.0×3.0×3.0 mm³ Acc:: 2 Rel. SNR: 1.00 |

### Properties

| | |
|---|---|
| Start measurement without further preparation | On |
| Wait for User to Start | On |
| Start measurements | Single Measurement |
| Prio Recon | Off |
| Auto Open Inline Display | On |
| Auto Close Inline Display | Off |
| Load Images to MR View&GO | On |
| Auto Store Images | On |
| Load Images to Stamp Segments | Off |
| Load Images to Graphic Segments | Off |
| Graphic segment | Default |
| Inline Movie | Off |

### Routine

| | |
|---|---|
| Slice Group | 1 |
| Slices | 41 |
| Distance Factor | 0 % |
| Position | R4.9 A16.5 H29.7 mm |
| Orientation | T > C-4.3 > S3.3 |
| Phase Encoding Dir. | A >> P |
| Phase Oversampling | 0 % |
| FoV Read | 192 mm |
| FoV Phase | 100.0 % |
| Slice Thickness | 3.0 mm |
| TR | 3000.0 ms |
| TE | 30.00 ms |
| Averages | 1 |
| Concatenations | 1 |
| AutoAlign | Head > Brain |

### Contrast - Common

| | |
|---|---|
| TR | 3000.0 ms |
| TE | 30.00 ms |
| MTC | Off |
| Flip Angle | 90 deg |
| Fat-Water Contrast | Fat Saturation |
| Reconstruction | Magnitude |

### Contrast - Dynamic

| | |
|---|---|
| Dynamic Mode | Standard |
| Measurements | 90 |
| Delay in TR | 790.00 ms |

### Resolution - Common

| | |
|---|---|
| FoV Read | 192 mm |
| FoV Phase | 100.0 % |
| Slice Thickness | 3.0 mm |
| Base Resolution | 64 |
| Phase Resolution | 100 % |
| Interpolation | Off |

### Resolution - Acceleration

| | |
|---|---|
| Acceleration mode | GRAPPA |
| Reference Scans | EPI/Separate |
| Acceleration Factor PE | 2 |
| Reference Lines PE | 24 |
| Advanced Reconstruction | Off |
| Phase Partial Fourier | Off |

### Resolution - Filter

| | |
|---|---|
| Raw Filter | Off |
| Elliptical Filter | Off |
| Hamming | Off |
| Distortion Correction | Off |
| Normalize | Prescan |

### Geometry - Common

| | |
|---|---|
| Slice Group | 1 |
| Slices | 41 |
| Distance Factor | 0 % |
| Position | R4.9 A16.5 H29.7 mm |
| Orientation | T > C-4.3 > S3.3 |
| Phase Encoding Dir. | A >> P |
| Phase Oversampling | 0 % |
| FoV Read | 192 mm |
| FoV Phase | 100.0 % |
| Slice Thickness | 3.0 mm |
| TR | 3000.0 ms |
| Multi-Slice Mode | Interleaved |
| Series | Interleaved |
| Concatenations | 1 |

### Geometry - AutoAlign

| | |
|---|---|
| Slice Group | 1 |
| Position | R4.9 A16.5 H29.7 mm |
| Orientation | T > C-4.3 > S3.3 |
| Phase Encoding Dir. | A >> P |
| AutoAlign | Head > Brain |
| Initial Position | R1.4 A2.6 H14.8 |
| R | 1.4 mm |
| A | 2.6 mm |
| H | 14.8 mm |
| Initial Orientation | T > S |
| T > S | -0.40 |
| > C | -0.10 |
| Initial Rotation | 0.14 deg |

### Geometry - Saturation

| | |
|---|---|
| Special Saturation | None |

### Geometry - Tim Planning Suite

| | |
|---|---|
| Set-n-Go Protocol | Off |
| Table Position | 0 mm |
| Table Position | H |
| Inline Composing | Off |

### System - Miscellaneous

| | |
|---|---|
| Coil Selection | ACS All but spine |
| MSMA | S - C - T |
| Sagittal | R >> L |
| Coronal | A >> P |
| Transversal | F >> H |
| Coil Combination | Sum of Squares |
| Matrix Optimization | Off |
| Coil Focus | Flat |

### System - Adjustments

| | |
|---|---|
| Adjustment Strategy | Standard |
| B0 Shim | Standard |
| B1 Shim | TrueForm |

### System - Adjustments

| | |
|---|---|
| CoilShim | Off |
| Adjustment Tolerance | Auto |
| Adjust with Body Coil | Off |
| Confirm Frequency | Never |
| Assume Silicone | Off |

### System - Adjust Volume

| | |
|---|---|
| Position | R4.9 A16.5 H29.7 mm |
| Orientation | T > C-4.3 > S3.3 |
| Rotation | -0.44 deg |
| A >> P | 192 mm |
| R >> L | 192 mm |
| F >> H | 123 mm |
| Reset | Off |

### System - pTx

| | |
|---|---|
| B1 Shim | TrueForm |
| Excitation | Standard |

### System - Tx/Rx

| | |
|---|---|
| Frequency 1H | 123.257916 MHz |
| ? Ref. Amplitude 1H | 0.000 V |
| Reset | Off |
| Image Scaling | 1.000 |

### Physio - Signal

| | |
|---|---|
| 1st Signal/Mode | None |
| TR | 3000.0 ms |
| Concatenations | 1 |

### BOLD

| | |
|---|---|
| GLM Statistics | On |
| Ignore Meas. at Start | 0 |
| Ignore After Transition | 0 |
| Model Transition States | On |
| Temp. Highpass Filter | On |
| Threshold | 4.00 |
| Paradigm Size | 20 |
| Meas[1] | Active |
| Meas[2] | Active |
| Meas[3] | Active |
| Meas[4] | Active |
| Meas[5] | Active |
| Meas[6] | Active |
| Meas[7] | Active |
| Meas[8] | Active |
| Meas[9] | Active |
| Meas[10] | Active |
| Meas[11] | Ignore |
| Meas[12] | Ignore |
| Meas[13] | Ignore |
| Meas[14] | Ignore |
| Meas[15] | Ignore |
| Meas[16] | Ignore |
| Meas[17] | Ignore |
| Meas[18] | Ignore |
| Meas[19] | Ignore |
| Meas[20] | Ignore |
| Motion Correction | Off |
| Spatial Filter | Off |
| Measurements | 90 |
| Delay in TR | 790.00 ms |

### Sequence - Part 1

| | |
|---|---|
| Sequence Name | epfid |
| Excitation | Standard |
| RF Pulse Type | Normal |
| Gradient Mode | Fast |
| Bandwidth | 2442 Hz/Px |
| Echo Spacing | 0.49 ms |
| Free Echo Spacing | Off |
| EPI Factor | 64 |

### Sequence - Part 2

| | |
|---|---|
| Introduction | Off |

**\\PHYSICS\XA30 RESEARCH PROT\PARCELLATION\BRAINGATE2 08-26-2024\TOE TAPPING_ RT FOOT FMRI**

TA: 4:39 min Coil Selection: Auto Voxel Size: 3.0×3.0×3.0 mm³ Acc:: 2 Rel. SNR: 1.00

### Properties

| | |
|---|---|
| Start measurement without further preparation | On |
| Wait for User to Start | On |
| Start measurements | Single Measurement |
| Prio Recon | Off |
| Auto Open Inline Display | On |
| Auto Close Inline Display | Off |
| Load Images to MR View&GO | On |
| Auto Store Images | On |
| Load Images to Stamp Segments | Off |
| Load Images to Graphic Segments | Off |
| Graphic segment | Default |
| Inline Movie | Off |

### Routine

| | |
|---|---|
| Slice Group | 1 |
| Slices | 41 |
| Distance Factor | 0 % |
| Position | R4.9 A16.5 H29.7 mm |
| Orientation | T > C-4.3 > S3.3 |
| Phase Encoding Dir. | A >> P |
| Phase Oversampling | 0 % |
| FoV Read | 192 mm |
| FoV Phase | 100.0 % |
| Slice Thickness | 3.0 mm |
| TR | 3000.0 ms |
| TE | 30.00 ms |
| Averages | 1 |
| Concatenations | 1 |
| AutoAlign | Head > Brain |

### Contrast - Common

| | |
|---|---|
| TR | 3000.0 ms |
| TE | 30.00 ms |
| MTC | Off |
| Flip Angle | 90 deg |
| Fat-Water Contrast | Fat Saturation |
| Reconstruction | Magnitude |

### Contrast - Dynamic

| | |
|---|---|
| Dynamic Mode | Standard |
| Measurements | 90 |
| Delay in TR | 790.00 ms |

### Resolution - Common

| | |
|---|---|
| FoV Read | 192 mm |
| FoV Phase | 100.0 % |
| Slice Thickness | 3.0 mm |
| Base Resolution | 64 |
| Phase Resolution | 100 % |
| Interpolation | Off |

### Resolution - Acceleration

| | |
|---|---|
| Acceleration mode | GRAPPA |
| Reference Scans | EPI/Separate |
| Acceleration Factor PE | 2 |
| Reference Lines PE | 24 |
| Advanced Reconstruction | Off |
| Phase Partial Fourier | Off |

### Resolution - Filter

| | |
|---|---|
| Raw Filter | Off |
| Elliptical Filter | Off |
| Hamming | Off |
| Distortion Correction | Off |
| Normalize | Prescan |

### Geometry - Common

| | |
|---|---|
| Slice Group | 1 |
| Slices | 41 |
| Distance Factor | 0 % |
| Position | R4.9 A16.5 H29.7 mm |
| Orientation | T > C-4.3 > S3.3 |
| Phase Encoding Dir. | A >> P |
| Phase Oversampling | 0 % |
| FoV Read | 192 mm |
| FoV Phase | 100.0 % |
| Slice Thickness | 3.0 mm |
| TR | 3000.0 ms |
| Multi-Slice Mode | Interleaved |
| Series | Interleaved |
| Concatenations | 1 |

### Geometry - AutoAlign

| | |
|---|---|
| Slice Group | 1 |
| Position | R4.9 A16.5 H29.7 mm |
| Orientation | T > C-4.3 > S3.3 |
| Phase Encoding Dir. | A >> P |
| AutoAlign | Head > Brain |
| Initial Position | R1.4 A2.6 H14.8 |
| R | 1.4 mm |
| A | 2.6 mm |
| H | 14.8 mm |
| Initial Orientation | T > S |
| T > S | -0.40 |
| > C | -0.10 |
| Initial Rotation | 0.14 deg |

### Geometry - Saturation

| | |
|---|---|
| Special Saturation | None |

### Geometry - Tim Planning Suite

| | |
|---|---|
| Set-n-Go Protocol | Off |
| Table Position | 0 mm |
| Table Position | H |
| Inline Composing | Off |

### System - Miscellaneous

| | |
|---|---|
| Coil Selection | ACS All but spine |
| MSMA | S - C - T |
| Sagittal | R >> L |
| Coronal | A >> P |
| Transversal | F >> H |
| Coil Combination | Sum of Squares |
| Matrix Optimization | Off |
| Coil Focus | Flat |

### System - Adjustments

| | |
|---|---|
| Adjustment Strategy | Standard |
| B0 Shim | Standard |

### System - Adjustments

| | |
|---|---|
| B1 Shim | TrueForm |
| CoilShim | Off |
| Adjustment Tolerance | Auto |
| Adjust with Body Coil | Off |
| Confirm Frequency | Never |
| Assume Silicone | Off |

### System - Adjust Volume

| | |
|---|---|
| Position | R4.9 A16.5 H29.7 mm |
| Orientation | T > C-4.3 > S3.3 |
| Rotation | -0.44 deg |
| A >> P | 192 mm |
| R >> L | 192 mm |
| F >> H | 123 mm |
| Reset | Off |

### System - pTx

| | |
|---|---|
| B1 Shim | TrueForm |
| Excitation | Standard |

### System - Tx/Rx

| | |
|---|---|
| Frequency 1H | 123.257916 MHz |
| ? Ref. Amplitude 1H | 0.000 V |
| Reset | Off |
| Image Scaling | 1.000 |

### Physio - Signal

| | |
|---|---|
| 1st Signal/Mode | None |
| TR | 3000.0 ms |
| Concatenations | 1 |

### BOLD

| | |
|---|---|
| GLM Statistics | On |
| Ignore Meas. at Start | 0 |
| Ignore After Transition | 0 |
| Model Transition States | On |
| Temp. Highpass Filter | On |
| Threshold | 4.00 |
| Paradigm Size | 20 |
| Meas[1] | Active |
| Meas[2] | Active |
| Meas[3] | Active |
| Meas[4] | Active |
| Meas[5] | Active |
| Meas[6] | Active |
| Meas[7] | Active |
| Meas[8] | Active |
| Meas[9] | Active |
| Meas[10] | Active |
| Meas[11] | Ignore |
| Meas[12] | Ignore |
| Meas[13] | Ignore |
| Meas[14] | Ignore |
| Meas[15] | Ignore |
| Meas[16] | Ignore |
| Meas[17] | Ignore |
| Meas[18] | Ignore |
| Meas[19] | Ignore |
| Meas[20] | Ignore |
| Motion Correction | Off |
| Spatial Filter | Off |
| Measurements | 90 |
| Delay in TR | 790.00 ms |

### Sequence - Part 1

| | |
|---|---|
| Sequence Name | epfid |
| Excitation | Standard |
| RF Pulse Type | Normal |
| Gradient Mode | Fast |
| Bandwidth | 2442 Hz/Px |
| Echo Spacing | 0.49 ms |
| Free Echo Spacing | Off |
| EPI Factor | 64 |

### Sequence - Part 2

| | |
|---|---|
| Introduction | Off |

**\\PHYSICS\XA30 RESEARCH PROT\PARCELLATION\BRAINGATE2 08-26-2024\REACH-GRASP RT H AND_FMRI**

TA: 6:27 min Coil Selection: Auto Voxel Size: 3.0×3.0×3.0 mm³ Acc:: 2 Rel. SNR: 1.00

### Properties

| | |
|---|---|
| Start measurement without further preparation | On |
| Wait for User to Start | On |
| Start measurements | Single Measurement |
| Prio Recon | Off |
| Auto Open Inline Display | On |
| Auto Close Inline Display | Off |
| Load Images to MR View&GO | On |
| Auto Store Images | On |
| Load Images to Stamp Segments | Off |
| Load Images to Graphic Segments | Off |
| Graphic segment | Default |
| Inline Movie | Off |

### Routine

| | |
|---|---|
| Slice Group | 1 |
| Slices | 41 |
| Distance Factor | 0 % |
| Position | R4.9 A16.5 H29.7 mm |
| Orientation | T > C-4.3 > S3.3 |
| Phase Encoding Dir. | A >> P |
| Phase Oversampling | 0 % |
| FoV Read | 192 mm |
| FoV Phase | 100.0 % |
| Slice Thickness | 3.0 mm |
| TR | 3000.0 ms |
| TE | 30.00 ms |
| Averages | 1 |
| Concatenations | 1 |
| AutoAlign | Head > Brain |

### Contrast - Common

| | |
|---|---|
| TR | 3000.0 ms |
| TE | 30.00 ms |
| MTC | Off |
| Flip Angle | 90 deg |
| Fat-Water Contrast | Fat Saturation |
| Reconstruction | Magnitude |

### Contrast - Dynamic

| | |
|---|---|
| Dynamic Mode | Standard |
| Measurements | 126 |
| Delay in TR | 790.00 ms |

### Resolution - Common

| | |
|---|---|
| FoV Read | 192 mm |
| FoV Phase | 100.0 % |
| Slice Thickness | 3.0 mm |
| Base Resolution | 64 |
| Phase Resolution | 100 % |
| Interpolation | Off |

### Resolution - Acceleration

| | |
|---|---|
| Acceleration mode | GRAPPA |
| Reference Scans | EPI/Separate |
| Acceleration Factor PE | 2 |
| Reference Lines PE | 24 |
| Advanced Reconstruction | Off |
| Phase Partial Fourier | Off |

### Resolution - Filter

| | |
|---|---|
| Raw Filter | Off |
| Elliptical Filter | Off |
| Hamming | Off |
| Distortion Correction | Off |
| Normalize | Prescan |

### Geometry - Common

| | |
|---|---|
| Slice Group | 1 |
| Slices | 41 |
| Distance Factor | 0 % |
| Position | R4.9 A16.5 H29.7 mm |
| Orientation | T > C-4.3 > S3.3 |
| Phase Encoding Dir. | A >> P |
| Phase Oversampling | 0 % |
| FoV Read | 192 mm |
| FoV Phase | 100.0 % |
| Slice Thickness | 3.0 mm |
| TR | 3000.0 ms |
| Multi-Slice Mode | Interleaved |
| Series | Interleaved |
| Concatenations | 1 |

### Geometry - AutoAlign

| | |
|---|---|
| Slice Group | 1 |
| Position | R4.9 A16.5 H29.7 mm |
| Orientation | T > C-4.3 > S3.3 |
| Phase Encoding Dir. | A >> P |
| AutoAlign | Head > Brain |
| Initial Position | R1.4 A2.6 H14.8 |
| R | 1.4 mm |
| A | 2.6 mm |
| H | 14.8 mm |
| Initial Orientation | T > S |
| T > S | -0.40 |
| > C | -0.10 |
| Initial Rotation | 0.14 deg |

### Geometry - Saturation

| | |
|---|---|
| Special Saturation | None |

### Geometry - Tim Planning Suite

| | |
|---|---|
| Set-n-Go Protocol | Off |
| Table Position | 0 mm |
| Table Position | H |
| Inline Composing | Off |

### System - Miscellaneous

| | |
|---|---|
| Coil Selection | ACS All but spine |
| MSMA | S - C - T |
| Sagittal | R >> L |
| Coronal | A >> P |
| Transversal | F >> H |
| Coil Combination | Sum of Squares |
| Matrix Optimization | Off |
| Coil Focus | Flat |

### System - Adjustments

| | |
|---|---|
| Adjustment Strategy | Standard |
| B0 Shim | Standard |

### System - Adjustments

| | |
|---|---|
| B1 Shim | TrueForm |
| CoilShim | Off |
| Adjustment Tolerance | Auto |
| Adjust with Body Coil | Off |
| Confirm Frequency | Never |
| Assume Silicone | Off |

### System - Adjust Volume

| | |
|---|---|
| Position | R4.9 A16.5 H29.7 mm |
| Orientation | T > C-4.3 > S3.3 |
| Rotation | -0.44 deg |
| A >> P | 192 mm |
| R >> L | 192 mm |
| F >> H | 123 mm |
| Reset | Off |

### System - pTx

| | |
|---|---|
| B1 Shim | TrueForm |
| Excitation | Standard |

### System - Tx/Rx

| | |
|---|---|
| Frequency 1H | 123.257916 MHz |
| ? Ref. Amplitude 1H | 0.000 V |
| Reset | Off |
| Image Scaling | 1.000 |

### Physio - Signal

| | |
|---|---|
| 1st Signal/Mode | None |
| TR | 3000.0 ms |
| Concatenations | 1 |

### BOLD

| | |
|---|---|
| GLM Statistics | On |
| Ignore Meas. at Start | 0 |
| Ignore After Transition | 0 |
| Model Transition States | On |
| Temp. Highpass Filter | On |
| Threshold | 4.00 |
| Paradigm Size | 20 |
| Meas[1] | Active |
| Meas[2] | Active |
| Meas[3] | Active |
| Meas[4] | Active |
| Meas[5] | Active |
| Meas[6] | Active |
| Meas[7] | Active |
| Meas[8] | Active |
| Meas[9] | Active |
| Meas[10] | Active |
| Meas[11] | Ignore |
| Meas[12] | Ignore |
| Meas[13] | Ignore |
| Meas[14] | Ignore |
| Meas[15] | Ignore |
| Meas[16] | Ignore |
| Meas[17] | Ignore |
| Meas[18] | Ignore |
| Meas[19] | Ignore |
| Meas[20] | Ignore |
| Motion Correction | Off |
| Spatial Filter | Off |
| Measurements | 126 |
| Delay in TR | 790.00 ms |

### Sequence - Part 1

| | |
|---|---|
| Sequence Name | epfid |
| Excitation | Standard |
| RF Pulse Type | Normal |
| Gradient Mode | Fast |
| Bandwidth | 2442 Hz/Px |
| Echo Spacing | 0.49 ms |
| Free Echo Spacing | Off |
| EPI Factor | 64 |

### Sequence - Part 2

| | |
|---|---|
| Introduction | Off |

| \\PHYSICS\XA30 RESEARCH PROT\PARCELLATION\BRAINGATE2 08-26-2024\REACH-GRASP LT HAND_FMRI |
|---|
| TA: 6:27 min Coil Selection: Auto Voxel Size: 3.0×3.0×3.0 mm³ Acc:: 2 Rel. SNR: 1.00 |

### Properties

| | |
|---|---|
| Start measurement without further preparation | On |
| Wait for User to Start | On |
| Start measurements | Single Measurement |
| Prio Recon | Off |
| Auto Open Inline Display | On |
| Auto Close Inline Display | Off |
| Load Images to MR View&GO | On |
| Auto Store Images | On |
| Load Images to Stamp Segments | Off |
| Load Images to Graphic Segments | Off |
| Graphic segment | Default |
| Inline Movie | Off |

### Routine

| | |
|---|---|
| Slice Group | 1 |
| Slices | 41 |
| Distance Factor | 0 % |
| Position | R4.9 A16.5 H29.7 mm |
| Orientation | T > C-4.3 > S3.3 |
| Phase Encoding Dir. | A >> P |
| Phase Oversampling | 0 % |
| FoV Read | 192 mm |
| FoV Phase | 100.0 % |
| Slice Thickness | 3.0 mm |
| TR | 3000.0 ms |
| TE | 30.00 ms |
| Averages | 1 |
| Concatenations | 1 |
| AutoAlign | Head > Brain |

### Contrast - Common

| | |
|---|---|
| TR | 3000.0 ms |
| TE | 30.00 ms |
| MTC | Off |
| Flip Angle | 90 deg |
| Fat-Water Contrast | Fat Saturation |
| Reconstruction | Magnitude |

### Contrast - Dynamic

| | |
|---|---|
| Dynamic Mode | Standard |
| Measurements | 126 |
| Delay in TR | 790.00 ms |

### Resolution - Common

| | |
|---|---|
| FoV Read | 192 mm |
| FoV Phase | 100.0 % |
| Slice Thickness | 3.0 mm |
| Base Resolution | 64 |
| Phase Resolution | 100 % |
| Interpolation | Off |

### Resolution - Acceleration

| | |
|---|---|
| Acceleration mode | GRAPPA |
| Reference Scans | EPI/Separate |
| Acceleration Factor PE | 2 |
| Reference Lines PE | 24 |
| Advanced Reconstruction | Off |
| Phase Partial Fourier | Off |

### Resolution - Filter

| | |
|---|---|
| Raw Filter | Off |
| Elliptical Filter | Off |
| Hamming | Off |
| Distortion Correction | Off |
| Normalize | Prescan |

### Geometry - Common

| | |
|---|---|
| Slice Group | 1 |
| Slices | 41 |
| Distance Factor | 0 % |
| Position | R4.9 A16.5 H29.7 mm |
| Orientation | T > C-4.3 > S3.3 |
| Phase Encoding Dir. | A >> P |
| Phase Oversampling | 0 % |
| FoV Read | 192 mm |
| FoV Phase | 100.0 % |
| Slice Thickness | 3.0 mm |
| TR | 3000.0 ms |
| Multi-Slice Mode | Interleaved |
| Series | Interleaved |
| Concatenations | 1 |

### Geometry - AutoAlign

| | |
|---|---|
| Slice Group | 1 |
| Position | R4.9 A16.5 H29.7 mm |
| Orientation | T > C-4.3 > S3.3 |
| Phase Encoding Dir. | A >> P |
| AutoAlign | Head > Brain |
| Initial Position | R1.4 A2.6 H14.8 |
| R | 1.4 mm |
| A | 2.6 mm |
| H | 14.8 mm |
| Initial Orientation | T > S |
| T > S | -0.40 |
| > C | -0.10 |
| Initial Rotation | 0.14 deg |

### Geometry - Saturation

| | |
|---|---|
| Special Saturation | None |

### Geometry - Tim Planning Suite

| | |
|---|---|
| Set-n-Go Protocol | Off |
| Table Position | 0 mm |
| Table Position | H |
| Inline Composing | Off |

### System - Miscellaneous

| | |
|---|---|
| Coil Selection | ACS All but spine |
| MSMA | S - C - T |
| Sagittal | R >> L |
| Coronal | A >> P |
| Transversal | F >> H |
| Coil Combination | Sum of Squares |
| Matrix Optimization | Off |
| Coil Focus | Flat |

### System - Adjustments

| | |
|---|---|
| Adjustment Strategy | Standard |
| B0 Shim | Standard |

### System - Adjustments

| | |
|---|---|
| B1 Shim | TrueForm |
| CoilShim | Off |
| Adjustment Tolerance | Auto |
| Adjust with Body Coil | Off |
| Confirm Frequency | Never |
| Assume Silicone | Off |

### System - Adjust Volume

| | |
|---|---|
| Position | R4.9 A16.5 H29.7 mm |
| Orientation | T > C-4.3 > S3.3 |
| Rotation | -0.44 deg |
| A >> P | 192 mm |
| R >> L | 192 mm |
| F >> H | 123 mm |
| Reset | Off |

### System - pTx

| | |
|---|---|
| B1 Shim | TrueForm |
| Excitation | Standard |

### System - Tx/Rx

| | |
|---|---|
| Frequency 1H | 123.257916 MHz |
| ? Ref. Amplitude 1H | 0.000 V |
| Reset | Off |
| Image Scaling | 1.000 |

### Physio - Signal

| | |
|---|---|
| 1st Signal/Mode | None |
| TR | 3000.0 ms |
| Concatenations | 1 |

### BOLD

| | |
|---|---|
| GLM Statistics | On |
| Ignore Meas. at Start | 0 |
| Ignore After Transition | 0 |
| Model Transition States | On |
| Temp. Highpass Filter | On |
| Threshold | 4.00 |
| Paradigm Size | 20 |
| Meas[1] | Active |
| Meas[2] | Active |
| Meas[3] | Active |
| Meas[4] | Active |
| Meas[5] | Active |
| Meas[6] | Active |
| Meas[7] | Active |
| Meas[8] | Active |
| Meas[9] | Active |
| Meas[10] | Active |
| Meas[11] | Ignore |
| Meas[12] | Ignore |
| Meas[13] | Ignore |
| Meas[14] | Ignore |
| Meas[15] | Ignore |
| Meas[16] | Ignore |
| Meas[17] | Ignore |
| Meas[18] | Ignore |
| Meas[19] | Ignore |
| Meas[20] | Ignore |
| Motion Correction | Off |
| Spatial Filter | Off |
| Measurements | 126 |
| Delay in TR | 790.00 ms |

### Sequence - Part 1

| | |
|---|---|
| Sequence Name | epfid |
| Excitation | Standard |
| RF Pulse Type | Normal |
| Gradient Mode | Fast |
| Bandwidth | 2442 Hz/Px |
| Echo Spacing | 0.49 ms |
| Free Echo Spacing | Off |
| EPI Factor | 64 |

### Sequence - Part 2

| | |
|---|---|
| Introduction | Off |

| \\PHYSICS\XA30 RESEARCH PROT\PARCELLATION\BRAINGATE2 08-26-2024\Ax_Flair |
|---|
| TA: 1:50 min Coil Selection: Auto Voxel Size: 0.4×0.4×5.0 mm³ Acc:: 3 Rel. SNR: 1.00 |

### Properties

| | |
|---|---|
| Start measurement without further preparation | On |
| Wait for User to Start | Off |
| Start measurements | Single Measurement |
| Prio Recon | Off |
| Auto Open Inline Display | Off |
| Auto Close Inline Display | Off |
| Load Images to MR View&GO | On |
| Auto Store Images | On |
| Load Images to Stamp Segments | Off |
| Load Images to Graphic Segments | Off |
| Graphic segment | Default |
| Inline Movie | Off |

### Routine

| | |
|---|---|
| Slice Group | 1 |
| Slices | 28 |
| Distance Factor | 20 % |
| Position | R2.6 A15.0 H14.9 mm |
| Orientation | T > C-4.2 > S3.7 |
| Phase Encoding Dir. | R >> L |
| Phase Oversampling | 0 % |
| FoV Read | 220 mm |
| FoV Phase | 100.0 % |
| Slice Thickness | 5.0 mm |
| TR | 9000.0 ms |
| TE | 88.00 ms |
| Averages | 1 |
| Concatenations | 2 |
| AutoAlign | Head > Brain |

### Contrast - Common

| | |
|---|---|
| TR | 9000.0 ms |
| TE | 88.00 ms |
| TD | 0.00 ms |
| MTC | Off |
| Magn. Preparation | Slice-sel. IR |
| TI | 2500 ms |
| Freeze Suppr. Tissue | On |
| Flip Angle | 150 deg |
| Fat-Water Contrast | Fat Saturation |
| Fat Saturation | Strong |
| Dark Blood | Off |
| Contrasts | 1 |
| Wrap-up Magn. | None |
| Reconstruction | Magnitude |

### Contrast - Dynamic

| | |
|---|---|
| Dynamic Mode | Standard |
| Measurements | 1 |
| Multiple Series | Each Measurement |

### Resolution - Common

| | |
|---|---|
| FoV Read | 220 mm |
| FoV Phase | 100.0 % |
| Slice Thickness | 5.0 mm |
| Base Resolution | 256 |
| Phase Resolution | 75 % |
| Trajectory | Cartesian |
| Interpolation | On |

### Resolution - Acceleration

| | |
|---|---|
| Acceleration mode | GRAPPA |
| Reference Scans | Integrated |
| Acceleration Factor PE | 3 |
| Reference Lines PE | 32 |
| Advanced Reconstruction | Off |
| Phase Partial Fourier | Off |

### Resolution - Filter

| | |
|---|---|
| Raw Filter | Off |
| Elliptical Filter | On |
| Distortion Correction | 2D |
| Normalize | Prescan |
| Image Filter | Off |

### Geometry - Common

| | |
|---|---|
| Slice Group | 1 |
| Slices | 28 |
| Distance Factor | 20 % |
| Position | R2.6 A15.0 H14.9 mm |
| Orientation | T > C-4.2 > S3.7 |
| Phase Encoding Dir. | R >> L |
| Phase Oversampling | 0 % |
| FoV Read | 220 mm |
| FoV Phase | 100.0 % |
| Slice Thickness | 5.0 mm |
| TR | 9000.0 ms |
| Multi-Slice Mode | Interleaved |
| Series | Interleaved |
| Concatenations | 2 |

### Geometry - AutoAlign

| | |
|---|---|
| Slice Group | 1 |
| Position | R2.6 A15.0 H14.9 mm |
| Orientation | T > C-4.2 > S3.7 |
| Phase Encoding Dir. | R >> L |
| AutoAlign | Head > Brain |
| Initial Position | Isocenter |
| R | 0.0 mm |
| P | 0.0 mm |
| H | 0.0 mm |
| Initial Orientation | Transversal |
| Initial Rotation | 90.00 deg |

### Geometry - Navigator

### Geometry - Saturation

| | |
|---|---|
| Special Saturation | None |

### Geometry - Tim Planning Suite

| | |
|---|---|
| Set-n-Go Protocol | Off |
| Table Position | 0 mm |
| Table Position | H |
| Inline Composing | Off |

### System - Miscellaneous

| | |
|---|---|
| Coil Selection | ACS All but spine |
| MSMA | S - C - T |
| Sagittal | R >> L |
| Coronal | A >> P |
| Transversal | F >> H |

### System - Miscellaneous

| | |
|---|---|
| Coil Combination | Adaptive Combine |
| Matrix Optimization | Off |
| Coil Focus | Flat |

### System - Adjustments

| | |
|---|---|
| Adjustment Strategy | Standard |
| B0 Shim | Standard |
| B1 Shim | TrueForm |
| CoilShim | Off |
| Adjustment Tolerance | Auto |
| Adjust with Body Coil | Off |
| Confirm Frequency | Never |
| Assume Silicone | Off |

### System - Adjust Volume

| | |
|---|---|
| Position | R2.6 A15.0 H14.9 mm |
| Orientation | T > C-4.2 > S3.7 |
| Rotation | 89.41 deg |
| R >> L | 220 mm |
| A >> P | 220 mm |
| F >> H | 167 mm |
| Reset | Off |

### System - pTx

| | |
|---|---|
| B1 Shim | TrueForm |
| LR Balancing | Off |

### System - Tx/Rx

| | |
|---|---|
| Frequency 1H | 123.257916 MHz |
| ? Ref. Amplitude 1H | 0.000 V |
| Reset | Off |
| Image Scaling | 1.000 |

### Physio - Signal

| | |
|---|---|
| 1st Signal/Mode | None |
| TR | 9000.0 ms |
| Concatenations | 2 |

### Physio - Cardiac

| | |
|---|---|
| Fat-Water Contrast | Fat Saturation |
| Magn. Preparation | Slice-sel. IR |
| TI | 2500 ms |
| Dark Blood | Off |
| FoV Read | 220 mm |
| FoV Phase | 100.0 % |
| Phase Resolution | 75 % |
| Trajectory | Cartesian |
| Dynamic Mode | Standard |

### Physio - PACE

| | |
|---|---|
| Resp. Control | Off |
| Concatenations | 2 |

### Inline - Subtraction

| | |
|---|---|
| Subtract | Off |
| Measurements | 1 |
| StdDev | Off |
| Save Original Images | On |

### Inline - Cardiac

| | |
|---|---|
| Magn. Preparation | Slice-sel. IR |
| Save Original Images | On |
| Contrasts | 1 |

### Inline - Cardiac

| | |
|---|---|
| TE | 88.00 ms |
| TR | 9000.0 ms |

### Inline - MIP

| | |
|---|---|
| MIP Sag | Off |
| MIP Cor | Off |
| MIP Tra | Off |
| MIP Time | Off |
| Radial MIP | Off |
| Save Original Images | On |
| MPR Sag | Off |
| MPR Cor | Off |
| MPR Tra | Off |

### Inline - Composing

| | |
|---|---|
| Inline Composing | Off |

### Sequence - Part 1

| | |
|---|---|
| Sequence Name | tir |
| Dimension | 2D |
| RF Pulse Type | Normal |
| Gradient Mode | Performance |
| Flow Compensation | None |
| Bandwidth | 271 Hz/Px |
| Echo Spacing | 8.02 ms |
| Free Echo Spacing | Off |
| Define | Turbo Factor |
| Turbo Factor | 17 |
| Echo Trains per Slice | 5 |

### Sequence - Part 2

| | |
|---|---|
| Introduction | On |
| Phase Correction | Automatic |
| Compensate T2 Decay | Off |
| Hyperecho | Off |
| WARP | Off |
| Red. EC Sensitivity | Off |
| Acoustic noise reduction | Off |
| Reduce Motion Sens. | On |

### Sequence - Assistant

| | |
|---|---|
| SAR Assistant | Off |
| Allowed Delay | 60 s |

**\\PHYSICS\XA30 RESEARCH PROT\PARCELLATION\BRAINGATE2 08-26-2024\Sag_Flair_3D_Space_Wave**

TA: 2:54 min Coil Selection: Auto Voxel Size: 1.0×1.0×1.0 mm³ Acc:: 4 Rel. SNR: 1.00

### Properties

| | |
|---|---|
| Start measurement without further preparation | On |
| Wait for User to Start | Off |
| Start measurements | Single Measurement |
| Prio Recon | Off |
| Auto Open Inline Display | Off |
| Auto Close Inline Display | Off |
| Load Images to MR View&GO | On |
| Auto Store Images | On |
| Load Images to Stamp Segments | Off |
| Load Images to Graphic Segments | Off |
| Graphic segment | Default |
| Inline Movie | Off |

### Routine

| | |
|---|---|
| Slab Group | 1 |
| Slabs | 1 |
| Position | R1.6 A29.4 F10.9 mm |
| Orientation | S > T-2.8 > C-0.8 |
| Phase Encoding Dir. | A >> P |
| Slices per Slab | 192 |
| Phase Oversampling | 0 % |
| Slice Oversampling | 0.0 % |
| FoV Read | 256 mm |
| FoV Phase | 100.0 % |
| Slice Thickness | 1.00 mm |
| TR | 5000.0 ms |
| TE | 388.00 ms |
| Averages | 1.0 |
| Concatenations | 1 |
| AutoAlign | Head > Basis |

### Contrast - Common

| | |
|---|---|
| TR | 5000.0 ms |
| TE | 388.00 ms |
| MTC | Off |
| Magn. Preparation | Non-sel. T2-IR |
| TI 1 | 1800 ms |
| Flip Angle Mode | T2 Var |
| Fat-Water Contrast | Standard |
| Dark Blood | Off |
| Blood Suppression | Off |
| Wrap-up Magn. | None |
| Reconstruction | Magnitude |

### Contrast - Dynamic

| | |
|---|---|
| Dynamic Mode | Standard |
| Measurements | 1 |
| Multiple Series | Each Measurement |
| Reordering | Linear |

### Resolution - Common

| | |
|---|---|
| FoV Read | 256 mm |
| FoV Phase | 100.0 % |
| Slice Thickness | 1.00 mm |
| Base Resolution | 256 |
| Phase Resolution | 100 % |
| Slice Resolution | 90 % |
| Interpolation | Off |

### Resolution - Acceleration

| | |
|---|---|
| Acceleration mode | CAIPIRINHA |
| Total Factor | 4 |
| Reference Scans | GRE/Separate |
| Acceleration Factor PE | 2 |
| Reference Lines PE | 24 |
| Acceleration Factor 3D | 2 |
| Reference Lines 3D | 48 |
| Reordering Shift 3D | 0 |
| Phase Partial Fourier | Allowed |
| Slice Partial Fourier | Off |
| Elliptical Scanning | On |

### Resolution - Filter

| | |
|---|---|
| Distortion Correction | 3D |
| Normalize | Prescan |
| Image Filter | Off |

### Geometry - Common

| | |
|---|---|
| Slab Group | 1 |
| Slabs | 1 |
| Position | R1.6 A29.4 F10.9 mm |
| Orientation | S > T-2.8 > C-0.8 |
| Phase Encoding Dir. | A >> P |
| Slices per Slab | 192 |
| Phase Oversampling | 0 % |
| Slice Oversampling | 0.0 % |
| FoV Read | 256 mm |
| FoV Phase | 100.0 % |
| Slice Thickness | 1.00 mm |
| TR | 5000.0 ms |
| Concatenations | 1 |

### Geometry - AutoAlign

| | |
|---|---|
| Slab Group | 1 |
| Position | R1.6 A29.4 F10.9 mm |
| Orientation | S > T-2.8 > C-0.8 |
| Phase Encoding Dir. | A >> P |
| AutoAlign | Head > Basis |
| Initial Position | L0.4 P0.4 H3.5 |
| L | 0.4 mm |
| P | 0.4 mm |
| H | 3.5 mm |
| Initial Orientation | S > T |
| S > T | 0.80 |
| > C | 0.10 |
| Initial Rotation | 6.56 deg |

### Geometry - Navigator

### Geometry - Saturation

| | |
|---|---|
| Special Saturation | None |

### Geometry - Tim Planning Suite

| | |
|---|---|
| Set-n-Go Protocol | Off |
| Table Position | 0 mm |
| Table Position | H |
| Inline Composing | Off |

### System - Miscellaneous

| | |
|---|---|
| Coil Selection | Auto Coil Select |
| MSMA | S - C - T |
| Sagittal | R >> L |
| Coronal | A >> P |
| Transversal | F >> H |
| Coil Combination | Sum of Squares |
| Matrix Optimization | Performance |
| Coil Focus | Flat |

### System - Adjustments

| | |
|---|---|
| Adjustment Strategy | Standard |
| B0 Shim | Tune up |
| B1 Shim | TrueForm |
| CoilShim | Off |
| Adjustment Tolerance | Auto |
| Adjust with Body Coil | Off |
| Confirm Frequency | Never |
| Assume Silicone | Off |

### System - Adjust Volume

| | |
|---|---|
| Position | Isocenter |
| Orientation | Transversal |
| Rotation | 0.00 deg |
| A >> P | 263 mm |
| R >> L | 350 mm |
| F >> H | 350 mm |
| Reset | Off |

### System - pTx

| | |
|---|---|
| B1 Shim | TrueForm |
| Excitation | Non-sel. |

### System - Tx/Rx

| | |
|---|---|
| Frequency 1H | 123.257916 MHz |
| ? Ref. Amplitude 1H | 0.000 V |
| Reset | Off |
| Image Scaling | 1.000 |

### Physio - Signal

| | |
|---|---|
| 1st Signal/Mode | None |
| Trigger Delay | 0 ms |
| TR | 5000.0 ms |
| Concatenations | 1 |

### Physio - Cardiac

| | |
|---|---|
| Fat-Water Contrast | Standard |
| Magn. Preparation | Non-sel. T2-IR |
| TI 1 | 1800 ms |
| Dark Blood | Off |
| FoV Read | 256 mm |
| FoV Phase | 100.0 % |
| Phase Resolution | 100 % |
| Dynamic Mode | Standard |

### Physio - PACE

| | |
|---|---|
| Resp. Control | Off |
| Concatenations | 1 |

### Inline - Subtraction

| | |
|---|---|
| Subtract | Off |
| Measurements | 1 |
| StdDev | Off |
| Save Original Images | On |

### Inline - Cardiac

| | |
|---|---|
| Magn. Preparation | Non-sel. T2-IR |
| Save Original Images | On |
| TE | 388.00 ms |
| TR | 5000.0 ms |

### Inline - MIP

| | |
|---|---|
| MIP Sag | Off |
| MIP Cor | Off |
| MIP Tra | Off |
| MIP Time | Off |
| Radial MIP | Off |
| Save Original Images | On |
| MPR Sag | Off |
| MPR Cor | Off |
| MPR Tra | Off |

### Inline - Composing

| | |
|---|---|
| Inline Composing | Off |

### Sequence - Part 1

| | |
|---|---|
| Sequence Name | WIP_spcir |
| Dimension | 3D |
| Excitation | Non-sel. |
| RF Pulse Type | Normal |
| Gradient Mode | Fast |
| Flow Compensation | None |
| Reordering | Linear |
| Bandwidth | 651 Hz/Px |
| Echo Spacing | 3.84 ms |
| Turbo Factor | 278 |
| Echo Train Duration | 918 ms |

### Sequence - Part 2

| | |
|---|---|
| Introduction | On |

### Sequence - Special

| | |
|---|---|
| Play Blip | Yes |
| Wave | Both |
| Coil Mixing | On |
| ROFilt Pre | 10 perc |
| ROFilt Post | 0 perc |
| LINFilt Pre | 0 perc |
| LINFilt Post | 0 perc |
| PARFilt Pre | 10 perc |
| PARFilt Post | 10 perc |
| Reg. Beta | 0.20 |
| Reg. Lambda | 0.20 |
| MAX Slew wave | 160.0 mT/m/ms |
| MAX Ampl wave | 20.0 mT/m |
| NumOfCycles Wave | 5.0 |

### Sequence - Assistant

| | |
|---|---|
| SAR Assistant | Off |
| Allowed Delay | 0 s |

**\\PHYSICS\XA30 RESEARCH PROT\PARCELLATION\BRAINGATE2 08-26-2024\Ax_DWI**

TA: 1:06 min Coil Selection: Auto Voxel Size: 1.7×1.7×5.0 mm³ Acc:: 4 Rel. SNR: 1.00

### Properties

| | |
|---|---|
| Start measurement without further preparation | Off |
| Wait for User to Start | Off |
| Start measurements | Single Measurement |
| Prio Recon | Off |
| Auto Open Inline Display | Off |
| Auto Close Inline Display | Off |
| Load Images to MR View&GO | On |
| Auto Store Images | On |
| Load Images to Stamp Segments | Off |
| Load Images to Graphic Segments | Off |
| Graphic segment | Default |
| Inline Movie | Off |

### Routine

| | |
|---|---|
| Slice Group | 1 |
| Slices | 28 |
| Distance Factor | 20 % |
| Position | R2.6 A15.0 H14.9 mm |
| Orientation | T > C-4.2 > S3.7 |
| Phase Encoding Dir. | A >> P |
| Phase Oversampling | 0 % |
| FoV Read | 272 mm |
| FoV Phase | 100.0 % |
| Slice Thickness | 5.0 mm |
| TR | 2000.0 ms |
| TE | 82.00 ms |
| Concatenations | 1 |
| AutoAlign | Head > Brain |

### Contrast - Common

| | |
|---|---|
| TR | 2000.0 ms |
| TE | 82.00 ms |
| MTC | Off |
| Magn. Preparation | None |
| Fat-Water Contrast | Fat Saturation |
| Fat Saturation | Weak |
| Reconstruction | Magnitude |

### Contrast - Dynamic

| | |
|---|---|
| Dynamic Mode | Standard |
| Multiple Series | Off |
| Delay in TR | 0.00 ms |

### Resolution - Common

| | |
|---|---|
| FoV Read | 272 mm |
| FoV Phase | 100.0 % |
| Slice Thickness | 5.0 mm |
| Base Resolution | 160 |
| Phase Resolution | 100 % |
| Interpolation | Off |

### Resolution - Acceleration

| | |
|---|---|
| Acceleration mode | SMS |
| Reference Scans | EPI/Separate |
| Acceleration Factor PE | 2 |
| Reference Lines PE | 40 |
| SMS Factor | 2 |
| Advanced Reconstruction | Off |
| Phase Partial Fourier | 6/8 |

### Resolution - Filter

| | |
|---|---|
| Raw Filter | Off |
| Elliptical Filter | Off |
| Distortion Correction | Off |
| Normalize | Prescan |
| Noise Masking | Off |

### Geometry - Common

| | |
|---|---|
| Slice Group | 1 |
| Slices | 28 |
| Distance Factor | 20 % |
| Position | R2.6 A15.0 H14.9 mm |
| Orientation | T > C-4.2 > S3.7 |
| Phase Encoding Dir. | A >> P |
| Phase Oversampling | 0 % |
| FoV Read | 272 mm |
| FoV Phase | 100.0 % |
| Slice Thickness | 5.0 mm |
| TR | 2000.0 ms |
| Multi-Slice Mode | Interleaved |
| Series | Interleaved |
| Concatenations | 1 |

### Geometry - AutoAlign

| | |
|---|---|
| Slice Group | 1 |
| Position | R2.6 A15.0 H14.9 mm |
| Orientation | T > C-4.2 > S3.7 |
| Phase Encoding Dir. | A >> P |
| AutoAlign | Head > Brain |
| Initial Position | Isocenter |
| R | 0.0 mm |
| P | 0.0 mm |
| H | 0.0 mm |
| Initial Orientation | Transversal |
| Initial Rotation | 0.00 deg |

### Geometry - Navigator

### Geometry - Saturation

| | |
|---|---|
| Special Saturation | None |

### Geometry - Tim Planning Suite

| | |
|---|---|
| Set-n-Go Protocol | Off |
| Table Position | 0 mm |
| Table Position | H |
| Inline Composing | Off |

### System - Miscellaneous

| | |
|---|---|
| Coil Selection | Auto Coil Select |
| MSMA | S - C - T |
| Sagittal | R >> L |
| Coronal | A >> P |
| Transversal | F >> H |
| Coil Combination | Adaptive Combine |
| Matrix Optimization | Off |
| Coil Focus | Flat |

### System - Adjustments

| | |
|---|---|
| Adjustment Strategy | Standard |
| B0 Shim | Standard |
| B1 Shim | TrueForm |

### System - Adjustments

| | |
|---|---|
| CoilShim | Off |
| Adjustment Tolerance | Auto |
| Adjust with Body Coil | Off |
| Confirm Frequency | Never |
| Assume Silicone | Off |

### System - Adjust Volume

| | |
|---|---|
| Position | R2.6 A15.0 H14.9 mm |
| Orientation | T > C-4.2 > S3.7 |
| Rotation | -0.59 deg |
| A >> P | 272 mm |
| R >> L | 272 mm |
| F >> H | 167 mm |
| Reset | Off |

### System - pTx

| | |
|---|---|
| B1 Shim | TrueForm |
| Excitation | Standard |

### System - Tx/Rx

| | |
|---|---|
| Frequency 1H | 123.257916 MHz |
| ? Ref. Amplitude 1H | 0.000 V |
| Reset | Off |
| Image Scaling | 1.000 |

### Physio - Signal

| | |
|---|---|
| 1st Signal/Mode | None |
| TR | 2000.0 ms |
| Concatenations | 1 |

### Physio - PACE

| | |
|---|---|
| Resp. Control | Off |
| Concatenations | 1 |

### Diff

| | |
|---|---|
| Diffusion Mode | Free |
| Diff. Directions | 25 |
| Diffusion Scheme | Bipolar |
| Diff. Weightings | 2 |
| b-value 1 | 0 s/mm² |
| b-value 2 | 1000 s/mm² |
| Averages 1 | 3 |
| Averages 2 | 1 |
| Dynamic Field Correction | Off |
| Invert Gray Scale | Off |
| Diff. Weighted Images | On |
| Trace Weighted Images | On |
| Tensor | Off |
| FA Maps | On |
| ADC Maps | On |
| Exponential ADC Maps | On |
| ADC Noise Threshold | 20 |
| Noise Masking | Off |
| Calculated Image | Off |

### Sequence - Part 1

| | |
|---|---|
| Sequence Name | epse |
| Excitation | Standard |
| RF Pulse Type | Normal |
| Gradient Mode | Fast |
| Bandwidth | 1736 Hz/Px |
| Echo Spacing | 0.93 ms |
| Free Echo Spacing | Off |

### Sequence - Part 1

| | |
|---|---|
| Optimization | None |
| EPI Factor | 160 |

### Sequence - Part 2

| | |
|---|---|
| Introduction | Off |
| Phase Correction | Internal |

| \\PHYSICS\XA30 RESEARCH PROT\PARCELLATION\BRAINGATE2 08-26-2024\Sag 3D PC MRV Head |
|---|
| TA: 9:03 min Coil Selection: Auto Voxel Size: 0.4×0.4×1.1 mm³ Acc:: 2 Rel. SNR: 1.00 |

### Properties

| | |
|---|---|
| Start measurement without further preparation | On |
| Wait for User to Start | Off |
| Start measurements | Single Measurement |
| Prio Recon | Off |
| Auto Open Inline Display | Off |
| Auto Close Inline Display | Off |
| Load Images to MR View&GO | On |
| Auto Store Images | On |
| Load Images to Stamp Segments | Off |
| Load Images to Graphic Segments | Off |
| Graphic segment | Default |
| Inline Movie | Off |

### Routine

| | |
|---|---|
| Slab Group | 1 |
| Slabs | 1 |
| Distance Factor | 20 % |
| Position | R2.5 A23.2 F4.0 mm |
| Orientation | S > C-3.8 > T-3.4 |
| Phase Encoding Dir. | A >> P |
| Slices per Slab | 144 |
| Phase Oversampling | 0 % |
| Slice Oversampling | 0.0 % |
| FoV Read | 230 mm |
| FoV Phase | 100.0 % |
| Slice Thickness | 1.1 mm |
| TR | 77.0 ms |
| TE | 8.65 ms |
| Averages | 1 |
| Concatenations | 1 |
| AutoAlign | Head > Basis |

### Contrast - Common

| | |
|---|---|
| TR | 77.0 ms |
| TE | 8.65 ms |
| Flip Angle | 15 deg |
| Contrasts | 1 |
| Reconstruction | Magnitude |

### Contrast - Dynamic

| | |
|---|---|
| Dynamic Mode | Standard |
| Measurements | 1 |
| Multiple Series | Each Measurement |

### Resolution - Common

| | |
|---|---|
| FoV Read | 230 mm |
| FoV Phase | 100.0 % |
| Slice Thickness | 1.1 mm |
| Base Resolution | 256 |
| Phase Resolution | 67 % |
| Slice Resolution | 50 % |
| Interpolation | On |

### Resolution - Acceleration

| | |
|---|---|
| Acceleration mode | GRAPPA |
| Reference Scans | Integrated |
| Acceleration Factor PE | 2 |
| Reference Lines PE | 24 |
| Acceleration Factor 3D | 1 |

### Resolution - Acceleration

| | |
|---|---|
| Phase Partial Fourier | Off |
| Asymmetric Echo | Weak |
| Elliptical Scanning | Off |

### Resolution - Filter

| | |
|---|---|
| Raw Filter | Off |
| Elliptical Filter | On |
| Distortion Correction | 2D |
| Normalize | Off |
| Image Filter | Off |

### Geometry - Common

| | |
|---|---|
| Slab Group | 1 |
| Slabs | 1 |
| Distance Factor | 20 % |
| Position | R2.5 A23.2 F4.0 mm |
| Orientation | S > C-3.8 > T-3.4 |
| Phase Encoding Dir. | A >> P |
| Slices per Slab | 144 |
| Phase Oversampling | 0 % |
| Slice Oversampling | 0.0 % |
| FoV Read | 230 mm |
| FoV Phase | 100.0 % |
| Slice Thickness | 1.1 mm |
| TR | 77.0 ms |
| Multi-Slice Mode | Sequential |
| Series | Interleaved |
| Concatenations | 1 |

### Geometry - AutoAlign

| | |
|---|---|
| Slab Group | 1 |
| Position | R2.5 A23.2 F4.0 mm |
| Orientation | S > C-3.8 > T-3.4 |
| Phase Encoding Dir. | A >> P |
| AutoAlign | Head > Basis |
| Initial Position | L0.0 P6.9 H10.0 |
| R | 0.0 mm |
| P | 6.9 mm |
| H | 10.0 mm |
| Initial Orientation | S > C |
| S > C | -2.90 |
| > T | 0.30 |
| Initial Rotation | 7.92 deg |

### Geometry - Saturation

| | |
|---|---|
| Saturation Region | 1 |
| Thickness | 50.00 mm |
| Position | L4.2 A27.8 F89.3 mm |
| Orientation | T > C-12.4 > S2.9 |
| Special Saturation | None |

### Geometry - Tim Planning Suite

| | |
|---|---|
| Set-n-Go Protocol | Off |
| Table Position | 0 mm |
| Table Position | H |
| Inline Composing | Off |

### System - Miscellaneous

| | |
|---|---|
| Coil Selection | Auto Coil Select |
| MSMA | S - C - T |

### System - Miscellaneous

| | |
|---|---|
| Sagittal | R >> L |
| Coronal | A >> P |
| Transversal | F >> H |
| Coil Combination | Adaptive Combine |
| Matrix Optimization | Off |
| Coil Focus | Flat |

### System - Adjustments

| | |
|---|---|
| Adjustment Strategy | Standard |
| B0 Shim | Tune up |
| B1 Shim | TrueForm |
| CoilShim | Off |
| Adjustment Tolerance | Auto |
| Adjust with Body Coil | Off |
| Confirm Frequency | Never |
| Assume Silicone | Off |

### System - Adjust Volume

| | |
|---|---|
| Position | Isocenter |
| Orientation | Transversal |
| Rotation | 0.00 deg |
| A >> P | 263 mm |
| R >> L | 350 mm |
| F >> H | 350 mm |
| Reset | Off |

### System - pTx

| | |
|---|---|
| B1 Shim | TrueForm |

### System - Tx/Rx

| | |
|---|---|
| Frequency 1H | 123.257916 MHz |
| ? Ref. Amplitude 1H | 0.000 V |
| Reset | Off |
| Image Scaling | 1.000 |

### Physio - Signal

| | |
|---|---|
| 1st Signal/Mode | None |
| TR | 77.0 ms |
| Segments | 1 |
| Concatenations | 1 |

### Angio - Common

| | |
|---|---|
| Flow Mode | Single Vel. |
| Encodings | 3 |
| Velocity Enc. | 15 cm/s |
| Direction 1 | Through Plane |
| Direction 2 | A >> P |
| Direction 3 | F >> H |
| Rephased Images | On |
| Magnitude Images | On |
| Magnitude Sum | On |
| Phase Images | On |

### Angio - Subtraction

| | |
|---|---|
| Subtract | Off |
| Measurements | 1 |
| StdDev | Off |
| Save Original Images | On |

### Angio - Cardiac

| | |
|---|---|
| Save Original Images | On |
| Contrasts | 1 |
| TE | 8.65 ms |

### Angio - Cardiac

| | |
|---|---|
| TR | 77.0 ms |

### Angio - MIP

| | |
|---|---|
| MIP Sag | On |
| MIP Cor | On |
| MIP Tra | On |
| MIP Time | Off |
| Radial MIP | Off |
| Save Original Images | On |
| MPR Sag | Off |
| MPR Cor | Off |
| MPR Tra | Off |

### Angio - Composing

| | |
|---|---|
| Inline Composing | Off |

### Sequence - Part 1

| | |
|---|---|
| Sequence Name | fi |
| Dimension | 3D |
| RF Pulse Type | Normal |
| Gradient Mode | Fast |
| Flow Compensation | None |
| Bandwidth | 300 Hz/Px |
| Asymmetric Echo | Weak |
| Segments | 1 |

### Sequence - Part 2

| | |
|---|---|
| Introduction | Off |
| RF Spoiling | Off |

### Sequence - Assistant

| | |
|---|---|
| SAR Assistant | Off |

| \\PHYSICS\XA30 RESEARCH PROT\PARCELLATION\BRAINGATE2 08-26-2024\Ax_SWI_Wave |
| --- |
| TA: 2:06 min Coil Selection: Auto Voxel Size: 0.9×0.9×1.8 mm³ Acc:: 6 Rel. SNR: 1.00 |

### Properties

| | |
| --- | --- |
| Start measurement without further preparation | On |
| Wait for User to Start | Off |
| Start measurements | Single Measurement |
| Prio Recon | Off |
| Auto Open Inline Display | Off |
| Auto Close Inline Display | Off |
| Load Images to MR View&GO | On |
| Auto Store Images | On |
| Load Images to Stamp Segments | Off |
| Load Images to Graphic Segments | Off |
| Graphic segment | Default |
| Inline Movie | Off |

### Routine

| | |
| --- | --- |
| Slab Group | 1 |
| Slabs | 1 |
| Distance Factor | 20 % |
| Position | R2.6 A15.0 H14.9 mm |
| Orientation | T > C-4.2 > S3.7 |
| Phase Encoding Dir. | R >> L |
| Slices per Slab | 96 |
| Phase Oversampling | 0 % |
| Slice Oversampling | 0.0 % |
| FoV Read | 250 mm |
| FoV Phase | 87.5 % |
| Slice Thickness | 1.8 mm |
| TR | 41.0 ms |
| TE 1 | 13.00 ms |
| TE 2 | 31.00 ms |
| Averages | 1 |
| Concatenations | 1 |
| AutoAlign | Head > Brain |

### Contrast - Common

| | |
| --- | --- |
| TR | 41.0 ms |
| TE 1 | 13.00 ms |
| TE 2 | 31.00 ms |
| MTC | Off |
| Magn. Preparation | None |
| Flip Angle | 15 deg |
| Fat-Water Contrast | Standard |
| Dark Blood | Off |
| Contrasts | 2 |
| SWI | On |
| Reconstruction | Magn./Phase |

### Contrast - Dynamic

| | |
| --- | --- |
| Dynamic Mode | Standard |
| Multiple Series | Each Measurement |

### Resolution - Common

| | |
| --- | --- |
| FoV Read | 250 mm |
| FoV Phase | 87.5 % |
| Slice Thickness | 1.8 mm |
| Base Resolution | 288 |
| Phase Resolution | 69 % |
| Slice Resolution | 100 % |
| Interpolation | Off |

### Resolution - Acceleration

| | |
| --- | --- |
| Acceleration mode | GRAPPA |
| Reference Scans | GRE/Separate |
| Acceleration Factor PE | 3 |
| Reference Lines PE | 30 |
| Acceleration Factor 3D | 2 |
| Reference Lines 3D | 48 |
| Advanced Reconstruction | Off |
| Phase Partial Fourier | Off |
| Slice Partial Fourier | Off |
| Asymmetric Echo | Off |
| Elliptical Scanning | Off |

### Resolution - Filter

| | |
| --- | --- |
| Distortion Correction | 2D |
| Normalize | Prescan |
| Image Filter | Off |

### Geometry - Common

| | |
| --- | --- |
| Slab Group | 1 |
| Slabs | 1 |
| Distance Factor | 20 % |
| Position | R2.6 A15.0 H14.9 mm |
| Orientation | T > C-4.2 > S3.7 |
| Phase Encoding Dir. | R >> L |
| Slices per Slab | 96 |
| Phase Oversampling | 0 % |
| Slice Oversampling | 0.0 % |
| FoV Read | 250 mm |
| FoV Phase | 87.5 % |
| Slice Thickness | 1.8 mm |
| TR | 41.0 ms |
| Multi-Slice Mode | Sequential |
| Series | Interleaved |
| Concatenations | 1 |

### Geometry - AutoAlign

| | |
| --- | --- |
| Slab Group | 1 |
| Position | R2.6 A15.0 H14.9 mm |
| Orientation | T > C-4.2 > S3.7 |
| Phase Encoding Dir. | R >> L |
| AutoAlign | Head > Brain |
| Initial Position | Isocenter |
| R | 0.0 mm |
| P | 0.0 mm |
| H | 0.0 mm |
| Initial Orientation | Transversal |
| Initial Rotation | 90.00 deg |

### Geometry - Saturation

| | |
| --- | --- |
| Saturation Mode | Standard |
| Special Saturation | None |

### Geometry - Tim Planning Suite

| | |
| --- | --- |
| Set-n-Go Protocol | Off |
| Table Position | 0 mm |
| Table Position | H |
| Inline Composing | Off |

### System - Miscellaneous

| | |
| --- | --- |
| Coil Selection | Auto Coil Select |

### System - Miscellaneous

| | |
|---|---|
| MSMA | S - C - T |
| Sagittal | R >> L |
| Coronal | A >> P |
| Transversal | F >> H |
| Coil Combination | Sum of Squares |
| Matrix Optimization | Performance |
| Coil Focus | Flat |

### System - Adjustments

| | |
|---|---|
| Adjustment Strategy | Standard |
| B0 Shim | Standard |
| B1 Shim | TrueForm |
| CoilShim | Off |
| Adjustment Tolerance | Auto |
| Adjust with Body Coil | Off |
| Confirm Frequency | Never |
| Assume Silicone | Off |

### System - Adjust Volume

| | |
|---|---|
| Position | R2.6 A15.0 H14.9 mm |
| Orientation | T > C-4.2 > S3.7 |
| Rotation | 89.41 deg |
| R >> L | 219 mm |
| A >> P | 250 mm |
| F >> H | 173 mm |
| Reset | Off |

### System - pTx

| | |
|---|---|
| B1 Shim | TrueForm |
| Excitation | Slab-sel. |
| LR Balancing | Off |

### System - Tx/Rx

| | |
|---|---|
| Frequency 1H | 123.257916 MHz |
| ? Ref. Amplitude 1H | 0.000 V |
| Reset | Off |
| Image Scaling | 1.000 |

### Physio - Cardiac

| | |
|---|---|
| Tagging | None |
| Fat-Water Contrast | Standard |
| Magn. Preparation | None |
| Dark Blood | Off |
| FoV Read | 250 mm |
| FoV Phase | 87.5 % |
| Phase Resolution | 69 % |
| Dynamic Mode | Standard |

### Physio - PACE

| | |
|---|---|
| Resp. Control | Off |
| Concatenations | 1 |

### Inline - Subtraction

### Inline - Cardiac

| | |
|---|---|
| Magn. Preparation | None |
| Contrasts | 2 |
| TE 1 | 13.00 ms |
| TE 2 | 31.00 ms |
| TR | 41.0 ms |

### Inline - Composing

| | |
|---|---|
| Inline Composing | Off |

### Sequence - Part 1

| | |
|---|---|
| Sequence Name | WIP_swi_r |
| Dimension | 3D |
| Excitation | Slab-sel. |
| RF Pulse Type | Normal |
| Readout Mode | Monopolar |
| Gradient Mode | Fast |
| Flow Compensation 1 | On |
| Flow Compensation 2 | On |
| Bandwidth 1 | 100 Hz/Px |
| Bandwidth 2 | 100 Hz/Px |
| Asymmetric Echo | Off |
| Segments | 1 |

### Sequence - Part 2

| | |
|---|---|
| Introduction | On |
| RF Spoiling | On |
| Acoustic noise reduction | Off |

### Sequence - Special

| | |
|---|---|
| Play Blip | Yes |
| Wave | Both |
| Coil Mixing | Off |
| Reg. Beta | 0.20 |
| Reg. Lambda | 0.20 |
| # of Dummies | 5 |
| Add OSF ACSz | 20 perc |
| MAX Slew wave | 50.0 mT/m/ms |
| MAX Ampl wave | 8.0 mT/m |
| NumOfCycles Wave | 31.0 |

### Sequence - Assistant

| | |
|---|---|
| SAR Assistant | Off |
| Allowed Delay | 0 s |

**\\PHYSICS\XA30 RESEARCH PROT\PARCELLATION\BRAINGATE2 08-26-2024\dMRI_dir95_AP**

TA: 5:28 min Coil Selection: Auto Voxel Size: 1.5×1.5×1.5 mm³ Acc:: None Rel. SNR: 1.00

### Properties

| | |
|---|---|
| Start measurement without further preparation | On |
| Wait for User to Start | Off |
| Start measurements | Single Measurement |
| Prio Recon | Off |
| Auto Open Inline Display | Off |
| Auto Close Inline Display | Off |
| Load Images to MR View&GO | On |
| Auto Store Images | On |
| Load Images to Stamp Segments | Off |
| Load Images to Graphic Segments | Off |
| Graphic segment | Default |
| Inline Movie | Off |

### Routine

| | |
|---|---|
| Slice Group | 1 |
| Slices | 92 |
| Distance Factor | 0 % |
| Position | R1.5 A2.6 H5.8 mm |
| Orientation | T > S-0.4 > C-0.1 |
| Phase Encoding Dir. | A >> P |
| Phase Oversampling | 0 % |
| FoV Read | 210 mm |
| FoV Phase | 100.0 % |
| Slice Thickness | 1.5 mm |
| TR | 3230.0 ms |
| TE | 89.20 ms |
| Multi-band accel. factor | 4 |
| AutoAlign | Head > Brain |

### Contrast - Common

| | |
|---|---|
| TR | 3230.0 ms |
| TE | 89.20 ms |
| MTC | Off |
| Magn. Preparation | None |
| Flip Angle | 78 deg |
| Refocus flip angle | 160 deg |
| Fat-Water Contrast | Standard |
| Grad. rev. fat suppr. | Enabled |
| Reconstruction | Magnitude |

### Contrast - Dynamic

| | |
|---|---|
| Dynamic Mode | Standard |
| Multiple Series | Off |
| Delay in TR | 0.00 ms |

### Resolution - Common

| | |
|---|---|
| FoV Read | 210 mm |
| FoV Phase | 100.0 % |
| Slice Thickness | 1.5 mm |
| Base Resolution | 140 |
| Phase Resolution | 100 % |
| Interpolation | Off |

### Resolution - Acceleration

| | |
|---|---|
| Acceleration mode | None |
| Advanced Reconstruction | Off |
| Phase Partial Fourier | 6/8 |

### Resolution - Filter

| | |
|---|---|
| Raw Filter | Off |
| Elliptical Filter | Off |
| Distortion Correction | Off |
| Normalize | Off |

### Geometry - Common

| | |
|---|---|
| Slice Group | 1 |
| Slices | 92 |
| Distance Factor | 0 % |
| Position | R1.5 A2.6 H5.8 mm |
| Orientation | T > S-0.4 > C-0.1 |
| Phase Encoding Dir. | A >> P |
| Phase Oversampling | 0 % |
| FoV Read | 210 mm |
| FoV Phase | 100.0 % |
| Slice Thickness | 1.5 mm |
| TR | 3230.0 ms |
| Multi-Slice Mode | Interleaved |
| Series | Interleaved |
| Multi-band accel. factor | 4 |

### Geometry - AutoAlign

| | |
|---|---|
| Slice Group | 1 |
| Position | R1.5 A2.6 H5.8 mm |
| Orientation | T > S-0.4 > C-0.1 |
| Phase Encoding Dir. | A >> P |
| AutoAlign | Head > Brain |
| Initial Position | R1.5 A2.6 H5.8 |
| R | 1.5 mm |
| A | 2.6 mm |
| H | 5.8 mm |
| Initial Orientation | T > S |
| T > S | -0.40 |
| > C | -0.10 |
| Initial Rotation | 0.14 deg |

### Geometry - Navigator

### Geometry - Saturation

| | |
|---|---|
| Special Saturation | None |

### Geometry - Tim Planning Suite

| | |
|---|---|
| Set-n-Go Protocol | Off |
| Table Position | 6 mm |
| Table Position | H |
| Inline Composing | Off |

### System - Miscellaneous

| | |
|---|---|
| Coil Selection | Auto Coil Select |
| MSMA | S - C - T |
| Sagittal | R >> L |
| Coronal | A >> P |
| Transversal | F >> H |
| Coil Combination | Sum of Squares |
| Matrix Optimization | Off |
| Coil Focus | Flat |

### System - Adjustments

| | |
|---|---|
| Adjustment Strategy | Standard |
| B0 Shim | Standard |

### System - Adjustments

| | |
|---|---|
| B1 Shim | TrueForm |
| CoilShim | Off |
| Adjustment Tolerance | Auto |
| Adjust with Body Coil | Off |
| Confirm Frequency | Never |
| Assume Silicone | Off |

### System - Adjust Volume

| | |
|---|---|
| Position | R1.5 A2.6 H5.8 mm |
| Orientation | T > S-0.4 > C-0.1 |
| Rotation | 0.14 deg |
| A >> P | 210 mm |
| R >> L | 210 mm |
| F >> H | 138 mm |
| Reset | Off |

### System - pTx

| | |
|---|---|
| B1 Shim | TrueForm |
| Excitation | Standard |

### System - Tx/Rx

| | |
|---|---|
| Frequency 1H | 123.257916 MHz |
| ? Ref. Amplitude 1H | 0.000 V |
| Reset | Off |
| Image Scaling | 1.000 |

### Physio - Signal

| | |
|---|---|
| 1st Signal/Mode | None |
| TR | 3230.0 ms |
| Multi-band accel. factor | 4 |

### Physio - PACE

| | |
|---|---|
| Resp. Control | Off |
| Multi-band accel. factor | 4 |

### Diff

| | |
|---|---|
| Diffusion Mode | Free |
| Diff. Directions | 95 |
| Diffusion Scheme | Monopolar |
| Diff. Weightings | 2 |
| b-value 1 | 0 s/mm² |
| b-value 2 | 1000 s/mm² |
| Averages 1 | 1 |
| Averages 2 | 1 |
| Dynamic Field Correction | Off |
| Invert Gray Scale | Off |
| Diff. Weighted Images | On |
| Trace Weighted Images | On |
| Tensor | On |
| FA Maps | On |
| ADC Maps | On |
| Exponential ADC Maps | Off |
| ADC Noise Threshold | 40 |
| Calculated Image | Off |

### Sequence - Part 1

| | |
|---|---|
| Sequence Name | epse |
| Dimension | 2D |
| Excitation | Standard |
| Gradient Mode | Performance |
| Bandwidth | 1786 Hz/Px |
| Echo Spacing | 0.66 ms |
| Free Echo Spacing | Off |

### Sequence - Part 1

| | |
|---|---|
| EPI Factor | 140 |

### Sequence - Part 2

| | |
|---|---|
| Introduction | On |
| RF Spoiling | Off |
| Phase Correction | Internal |

### Sequence - Special

| | |
|---|---|
| Excite pulse duration | 4480 us |
| Refocus pulse duration | 8320 us |
| Min. prep scans | 0 |
| Min. prep scans SB | 0 |
| Delay before PC scans | 0 us |
| Single-band images | On |
| MB LeakBlock kernel | On |
| MB dual kernel | Off |
| MB RF phase scramble | Off |
| Opt. MB RF pulse BW | Off |
| Time-shifted MB RF | Off |
| SENSE1 coil combine | On |
| Invert RO/PE polarity | Off |
| PF omits higher k-space | Off |
| Disable freq. update | Off |
| Suppress 16-bit DICOM | Off |
| Force equal slice timing | Off |
| Online multi-band recon. | Online |
| FFT scale factor | 1.00 |
| Physio recording | DICOM |

### Sequence - Assistant

| | |
|---|---|
| SAR Assistant | Off |

| \\PHYSICS\XA30 RESEARCH PROT\PARCELLATION\BRAINGATE2 08-26-2024\dMRI_dir95_PA |
|---|
| TA: 5:28 min Coil Selection: Auto Voxel Size: 1.5×1.5×1.5 mm³ Acc:: None Rel. SNR: 1.00 |

### Properties

| | |
|---|---|
| Start measurement without further preparation | On |
| Wait for User to Start | Off |
| Start measurements | Single Measurement |
| Prio Recon | Off |
| Auto Open Inline Display | Off |
| Auto Close Inline Display | Off |
| Load Images to MR View&GO | On |
| Auto Store Images | On |
| Load Images to Stamp Segments | Off |
| Load Images to Graphic Segments | Off |
| Graphic segment | Default |
| Inline Movie | Off |

### Routine

| | |
|---|---|
| Slice Group | 1 |
| Slices | 92 |
| Distance Factor | 0 % |
| Position | R1.5 A2.6 H5.8 mm |
| Orientation | T > S-0.4 > C-0.1 |
| Phase Encoding Dir. | A >> P |
| Phase Oversampling | 0 % |
| FoV Read | 210 mm |
| FoV Phase | 100.0 % |
| Slice Thickness | 1.5 mm |
| TR | 3230.0 ms |
| TE | 89.20 ms |
| Multi-band accel. factor | 4 |
| AutoAlign | Head > Brain |

### Contrast - Common

| | |
|---|---|
| TR | 3230.0 ms |
| TE | 89.20 ms |
| MTC | Off |
| Magn. Preparation | None |
| Flip Angle | 78 deg |
| Refocus flip angle | 160 deg |
| Fat-Water Contrast | Standard |
| Grad. rev. fat suppr. | Enabled |
| Reconstruction | Magnitude |

### Contrast - Dynamic

| | |
|---|---|
| Dynamic Mode | Standard |
| Multiple Series | Off |
| Delay in TR | 0.00 ms |

### Resolution - Common

| | |
|---|---|
| FoV Read | 210 mm |
| FoV Phase | 100.0 % |
| Slice Thickness | 1.5 mm |
| Base Resolution | 140 |
| Phase Resolution | 100 % |
| Interpolation | Off |

### Resolution - Acceleration

| | |
|---|---|
| Acceleration mode | None |
| Advanced Reconstruction | Off |
| Phase Partial Fourier | 6/8 |

### Resolution - Filter

| | |
|---|---|
| Raw Filter | Off |
| Elliptical Filter | Off |
| Distortion Correction | Off |
| Normalize | Off |

### Geometry - Common

| | |
|---|---|
| Slice Group | 1 |
| Slices | 92 |
| Distance Factor | 0 % |
| Position | R1.5 A2.6 H5.8 mm |
| Orientation | T > S-0.4 > C-0.1 |
| Phase Encoding Dir. | A >> P |
| Phase Oversampling | 0 % |
| FoV Read | 210 mm |
| FoV Phase | 100.0 % |
| Slice Thickness | 1.5 mm |
| TR | 3230.0 ms |
| Multi-Slice Mode | Interleaved |
| Series | Interleaved |
| Multi-band accel. factor | 4 |

### Geometry - AutoAlign

| | |
|---|---|
| Slice Group | 1 |
| Position | R1.5 A2.6 H5.8 mm |
| Orientation | T > S-0.4 > C-0.1 |
| Phase Encoding Dir. | A >> P |
| AutoAlign | Head > Brain |
| Initial Position | R1.5 A2.6 H5.8 |
| R | 1.5 mm |
| A | 2.6 mm |
| H | 5.8 mm |
| Initial Orientation | T > S |
| T > S | -0.40 |
| > C | -0.10 |
| Initial Rotation | 0.14 deg |

### Geometry - Navigator

### Geometry - Saturation

| | |
|---|---|
| Special Saturation | None |

### Geometry - Tim Planning Suite

| | |
|---|---|
| Set-n-Go Protocol | Off |
| Table Position | 6 mm |
| Table Position | H |
| Inline Composing | Off |

### System - Miscellaneous

| | |
|---|---|
| Coil Selection | Auto Coil Select |
| MSMA | S - C - T |
| Sagittal | R >> L |
| Coronal | A >> P |
| Transversal | F >> H |
| Coil Combination | Sum of Squares |
| Matrix Optimization | Off |
| Coil Focus | Flat |

### System - Adjustments

| | |
|---|---|
| Adjustment Strategy | Standard |
| B0 Shim | Standard |

### System - Adjustments

| | |
|---|---|
| B1 Shim | TrueForm |
| CoilShim | Off |
| Adjustment Tolerance | Auto |
| Adjust with Body Coil | Off |
| Confirm Frequency | Never |
| Assume Silicone | Off |

### System - Adjust Volume

| | |
|---|---|
| Position | R1.5 A2.6 H5.8 mm |
| Orientation | T > S-0.4 > C-0.1 |
| Rotation | 0.14 deg |
| A >> P | 210 mm |
| R >> L | 210 mm |
| F >> H | 138 mm |
| Reset | Off |

### System - pTx

| | |
|---|---|
| B1 Shim | TrueForm |
| Excitation | Standard |

### System - Tx/Rx

| | |
|---|---|
| Frequency 1H | 123.257916 MHz |
| ? Ref. Amplitude 1H | 0.000 V |
| Reset | Off |
| Image Scaling | 1.000 |

### Physio - Signal

| | |
|---|---|
| 1st Signal/Mode | None |
| TR | 3230.0 ms |
| Multi-band accel. factor | 4 |

### Physio - PACE

| | |
|---|---|
| Resp. Control | Off |
| Multi-band accel. factor | 4 |

### Diff

| | |
|---|---|
| Diffusion Mode | Free |
| Diff. Directions | 95 |
| Diffusion Scheme | Monopolar |
| Diff. Weightings | 2 |
| b-value 1 | 0 s/mm² |
| b-value 2 | 1000 s/mm² |
| Averages 1 | 1 |
| Averages 2 | 1 |
| Dynamic Field Correction | Off |
| Invert Gray Scale | Off |
| Diff. Weighted Images | On |
| Trace Weighted Images | On |
| Tensor | On |
| FA Maps | On |
| ADC Maps | On |
| Exponential ADC Maps | Off |
| ADC Noise Threshold | 40 |
| Calculated Image | Off |

### Sequence - Part 1

| | |
|---|---|
| Sequence Name | epse |
| Dimension | 2D |
| Excitation | Standard |
| Gradient Mode | Performance |
| Bandwidth | 1786 Hz/Px |
| Echo Spacing | 0.66 ms |
| Free Echo Spacing | Off |

### Sequence - Part 1

| | |
|---|---|
| EPI Factor | 140 |

### Sequence - Part 2

| | |
|---|---|
| Introduction | On |
| RF Spoiling | Off |
| Phase Correction | Internal |

### Sequence - Special

| | |
|---|---|
| Excite pulse duration | 4480 us |
| Refocus pulse duration | 8320 us |
| Min. prep scans | 0 |
| Min. prep scans SB | 0 |
| Delay before PC scans | 0 us |
| Single-band images | On |
| MB LeakBlock kernel | On |
| MB dual kernel | Off |
| MB RF phase scramble | Off |
| Opt. MB RF pulse BW | Off |
| Time-shifted MB RF | Off |
| SENSE1 coil combine | On |
| Invert RO/PE polarity | On |
| PF omits higher k-space | Off |
| Disable freq. update | Off |
| Suppress 16-bit DICOM | Off |
| Force equal slice timing | Off |
| Online multi-band recon. | Online |
| FFT scale factor | 1.00 |
| Physio recording | DICOM |

### Sequence - Assistant

| | |
|---|---|
| SAR Assistant | Off |

| \\PHYSICS\XA30 RESEARCH PROT\PARCELLATION\BRAINGATE2 08-26-2024\dMRI_dir96_AP |
|---|
| TA: 5:31 min Coil Selection: Auto Voxel Size: 1.5×1.5×1.5 mm³ Acc:: None Rel. SNR: 1.00 |

### Properties

| | |
|---|---|
| Start measurement without further preparation | On |
| Wait for User to Start | Off |
| Start measurements | Single Measurement |
| Prio Recon | Off |
| Auto Open Inline Display | Off |
| Auto Close Inline Display | Off |
| Load Images to MR View&GO | On |
| Auto Store Images | On |
| Load Images to Stamp Segments | Off |
| Load Images to Graphic Segments | Off |
| Graphic segment | Default |
| Inline Movie | Off |

### Routine

| | |
|---|---|
| Slice Group | 1 |
| Slices | 92 |
| Distance Factor | 0 % |
| Position | R1.5 A2.6 H5.8 mm |
| Orientation | T > S-0.4 > C-0.1 |
| Phase Encoding Dir. | A >> P |
| Phase Oversampling | 0 % |
| FoV Read | 210 mm |
| FoV Phase | 100.0 % |
| Slice Thickness | 1.5 mm |
| TR | 3230.0 ms |
| TE | 89.20 ms |
| Multi-band accel. factor | 4 |
| AutoAlign | Head > Brain |

### Contrast - Common

| | |
|---|---|
| TR | 3230.0 ms |
| TE | 89.20 ms |
| MTC | Off |
| Magn. Preparation | None |
| Flip Angle | 78 deg |
| Refocus flip angle | 160 deg |
| Fat-Water Contrast | Standard |
| Grad. rev. fat suppr. | Enabled |
| Reconstruction | Magnitude |

### Contrast - Dynamic

| | |
|---|---|
| Dynamic Mode | Standard |
| Multiple Series | Off |
| Delay in TR | 0.00 ms |

### Resolution - Common

| | |
|---|---|
| FoV Read | 210 mm |
| FoV Phase | 100.0 % |
| Slice Thickness | 1.5 mm |
| Base Resolution | 140 |
| Phase Resolution | 100 % |
| Interpolation | Off |

### Resolution - Acceleration

| | |
|---|---|
| Acceleration mode | None |
| Advanced Reconstruction | Off |
| Phase Partial Fourier | 6/8 |

### Resolution - Filter

| | |
|---|---|
| Raw Filter | Off |
| Elliptical Filter | Off |
| Distortion Correction | Off |
| Normalize | Off |

### Geometry - Common

| | |
|---|---|
| Slice Group | 1 |
| Slices | 92 |
| Distance Factor | 0 % |
| Position | R1.5 A2.6 H5.8 mm |
| Orientation | T > S-0.4 > C-0.1 |
| Phase Encoding Dir. | A >> P |
| Phase Oversampling | 0 % |
| FoV Read | 210 mm |
| FoV Phase | 100.0 % |
| Slice Thickness | 1.5 mm |
| TR | 3230.0 ms |
| Multi-Slice Mode | Interleaved |
| Series | Interleaved |
| Multi-band accel. factor | 4 |

### Geometry - AutoAlign

| | |
|---|---|
| Slice Group | 1 |
| Position | R1.5 A2.6 H5.8 mm |
| Orientation | T > S-0.4 > C-0.1 |
| Phase Encoding Dir. | A >> P |
| AutoAlign | Head > Brain |
| Initial Position | R1.5 A2.6 H5.8 |
| R | 1.5 mm |
| A | 2.6 mm |
| H | 5.8 mm |
| Initial Orientation | T > S |
| T > S | -0.40 |
| > C | -0.10 |
| Initial Rotation | 0.14 deg |

### Geometry - Navigator

### Geometry - Saturation

| | |
|---|---|
| Special Saturation | None |

### Geometry - Tim Planning Suite

| | |
|---|---|
| Set-n-Go Protocol | Off |
| Table Position | 6 mm |
| Table Position | H |
| Inline Composing | Off |

### System - Miscellaneous

| | |
|---|---|
| Coil Selection | Auto Coil Select |
| MSMA | S - C - T |
| Sagittal | R >> L |
| Coronal | A >> P |
| Transversal | F >> H |
| Coil Combination | Sum of Squares |
| Matrix Optimization | Off |
| Coil Focus | Flat |

### System - Adjustments

| | |
|---|---|
| Adjustment Strategy | Standard |
| B0 Shim | Standard |

### System - Adjustments

| | |
|---|---|
| B1 Shim | TrueForm |
| CoilShim | Off |
| Adjustment Tolerance | Auto |
| Adjust with Body Coil | Off |
| Confirm Frequency | Never |
| Assume Silicone | Off |

### System - Adjust Volume

| | |
|---|---|
| Position | R1.5 A2.6 H5.8 mm |
| Orientation | T > S-0.4 > C-0.1 |
| Rotation | 0.14 deg |
| A >> P | 210 mm |
| R >> L | 210 mm |
| F >> H | 138 mm |
| Reset | Off |

### System - pTx

| | |
|---|---|
| B1 Shim | TrueForm |
| Excitation | Standard |

### System - Tx/Rx

| | |
|---|---|
| Frequency 1H | 123.257916 MHz |
| ? Ref. Amplitude 1H | 0.000 V |
| Reset | Off |
| Image Scaling | 1.000 |

### Physio - Signal

| | |
|---|---|
| 1st Signal/Mode | None |
| TR | 3230.0 ms |
| Multi-band accel. factor | 4 |

### Physio - PACE

| | |
|---|---|
| Resp. Control | Off |
| Multi-band accel. factor | 4 |

### Diff

| | |
|---|---|
| Diffusion Mode | Free |
| Diff. Directions | 96 |
| Diffusion Scheme | Monopolar |
| Diff. Weightings | 2 |
| b-value 1 | 0 s/mm² |
| b-value 2 | 2000 s/mm² |
| Averages 1 | 1 |
| Averages 2 | 1 |
| Dynamic Field Correction | Off |
| Invert Gray Scale | Off |
| Diff. Weighted Images | On |
| Trace Weighted Images | On |
| Tensor | On |
| FA Maps | On |
| ADC Maps | On |
| Exponential ADC Maps | Off |
| ADC Noise Threshold | 40 |
| Calculated Image | Off |

### Sequence - Part 1

| | |
|---|---|
| Sequence Name | epse |
| Dimension | 2D |
| Excitation | Standard |
| Gradient Mode | Performance |
| Bandwidth | 1786 Hz/Px |
| Echo Spacing | 0.66 ms |
| Free Echo Spacing | Off |

### Sequence - Part 1

| | |
|---|---|
| EPI Factor | 140 |

### Sequence - Part 2

| | |
|---|---|
| Introduction | On |
| RF Spoiling | Off |
| Phase Correction | Internal |

### Sequence - Special

| | |
|---|---|
| Excite pulse duration | 4480 us |
| Refocus pulse duration | 8320 us |
| Min. prep scans | 0 |
| Min. prep scans SB | 0 |
| Delay before PC scans | 0 us |
| Single-band images | On |
| MB LeakBlock kernel | On |
| MB dual kernel | Off |
| MB RF phase scramble | Off |
| Opt. MB RF pulse BW | Off |
| Time-shifted MB RF | Off |
| SENSE1 coil combine | On |
| Invert RO/PE polarity | Off |
| PF omits higher k-space | Off |
| Disable freq. update | Off |
| Suppress 16-bit DICOM | Off |
| Force equal slice timing | Off |
| Online multi-band recon. | Online |
| FFT scale factor | 1.00 |
| Physio recording | DICOM |

### Sequence - Assistant

| | |
|---|---|
| SAR Assistant | Off |

| \\PHYSICS\XA30 RESEARCH PROT\PARCELLATION\BRAINGATE2 08-26-2024\dMRI_dir96_PA |
|---|
| TA: 5:31 min Coil Selection: Auto Voxel Size: 1.5×1.5×1.5 mm³ Acc:: None Rel. SNR: 1.00 |

### Properties

| | |
|---|---|
| Start measurement without further preparation | On |
| Wait for User to Start | Off |
| Start measurements | Single Measurement |
| Prio Recon | Off |
| Auto Open Inline Display | Off |
| Auto Close Inline Display | Off |
| Load Images to MR View&GO | On |
| Auto Store Images | On |
| Load Images to Stamp Segments | Off |
| Load Images to Graphic Segments | Off |
| Graphic segment | Default |
| Inline Movie | Off |

### Routine

| | |
|---|---|
| Slice Group | 1 |
| Slices | 92 |
| Distance Factor | 0 % |
| Position | R1.5 A2.6 H5.8 mm |
| Orientation | T > S-0.4 > C-0.1 |
| Phase Encoding Dir. | A >> P |
| Phase Oversampling | 0 % |
| FoV Read | 210 mm |
| FoV Phase | 100.0 % |
| Slice Thickness | 1.5 mm |
| TR | 3230.0 ms |
| TE | 89.20 ms |
| Multi-band accel. factor | 4 |
| AutoAlign | Head > Brain |

### Contrast - Common

| | |
|---|---|
| TR | 3230.0 ms |
| TE | 89.20 ms |
| MTC | Off |
| Magn. Preparation | None |
| Flip Angle | 78 deg |
| Refocus flip angle | 160 deg |
| Fat-Water Contrast | Standard |
| Grad. rev. fat suppr. | Enabled |
| Reconstruction | Magnitude |

### Contrast - Dynamic

| | |
|---|---|
| Dynamic Mode | Standard |
| Multiple Series | Off |
| Delay in TR | 0.00 ms |

### Resolution - Common

| | |
|---|---|
| FoV Read | 210 mm |
| FoV Phase | 100.0 % |
| Slice Thickness | 1.5 mm |
| Base Resolution | 140 |
| Phase Resolution | 100 % |
| Interpolation | Off |

### Resolution - Acceleration

| | |
|---|---|
| Acceleration mode | None |
| Advanced Reconstruction | Off |
| Phase Partial Fourier | 6/8 |

### Resolution - Filter

| | |
|---|---|
| Raw Filter | Off |
| Elliptical Filter | Off |
| Distortion Correction | Off |
| Normalize | Off |

### Geometry - Common

| | |
|---|---|
| Slice Group | 1 |
| Slices | 92 |
| Distance Factor | 0 % |
| Position | R1.5 A2.6 H5.8 mm |
| Orientation | T > S-0.4 > C-0.1 |
| Phase Encoding Dir. | A >> P |
| Phase Oversampling | 0 % |
| FoV Read | 210 mm |
| FoV Phase | 100.0 % |
| Slice Thickness | 1.5 mm |
| TR | 3230.0 ms |
| Multi-Slice Mode | Interleaved |
| Series | Interleaved |
| Multi-band accel. factor | 4 |

### Geometry - AutoAlign

| | |
|---|---|
| Slice Group | 1 |
| Position | R1.5 A2.6 H5.8 mm |
| Orientation | T > S-0.4 > C-0.1 |
| Phase Encoding Dir. | A >> P |
| AutoAlign | Head > Brain |
| Initial Position | R1.5 A2.6 H5.8 |
| R | 1.5 mm |
| A | 2.6 mm |
| H | 5.8 mm |
| Initial Orientation | T > S |
| T > S | -0.40 |
| > C | -0.10 |
| Initial Rotation | 0.14 deg |

### Geometry - Navigator

### Geometry - Saturation

| | |
|---|---|
| Special Saturation | None |

### Geometry - Tim Planning Suite

| | |
|---|---|
| Set-n-Go Protocol | Off |
| Table Position | 6 mm |
| Table Position | H |
| Inline Composing | Off |

### System - Miscellaneous

| | |
|---|---|
| Coil Selection | Auto Coil Select |
| MSMA | S - C - T |
| Sagittal | R >> L |
| Coronal | A >> P |
| Transversal | F >> H |
| Coil Combination | Sum of Squares |
| Matrix Optimization | Off |
| Coil Focus | Flat |

### System - Adjustments

| | |
|---|---|
| Adjustment Strategy | Standard |
| B0 Shim | Standard |

#### System - Adjustments

| | |
|---|---|
| B1 Shim | TrueForm |
| CoilShim | Off |
| Adjustment Tolerance | Auto |
| Adjust with Body Coil | Off |
| Confirm Frequency | Never |
| Assume Silicone | Off |

#### System - Adjust Volume

| | |
|---|---|
| Position | R1.5 A2.6 H5.8 mm |
| Orientation | T > S-0.4 > C-0.1 |
| Rotation | 0.14 deg |
| A >> P | 210 mm |
| R >> L | 210 mm |
| F >> H | 138 mm |
| Reset | Off |

#### System - pTx

| | |
|---|---|
| B1 Shim | TrueForm |
| Excitation | Standard |

#### System - Tx/Rx

| | |
|---|---|
| Frequency 1H | 123.257916 MHz |
| ? Ref. Amplitude 1H | 0.000 V |
| Reset | Off |
| Image Scaling | 1.000 |

#### Physio - Signal

| | |
|---|---|
| 1st Signal/Mode | None |
| TR | 3230.0 ms |
| Multi-band accel. factor | 4 |

#### Physio - PACE

| | |
|---|---|
| Resp. Control | Off |
| Multi-band accel. factor | 4 |

#### Diff

| | |
|---|---|
| Diffusion Mode | Free |
| Diff. Directions | 96 |
| Diffusion Scheme | Monopolar |
| Diff. Weightings | 2 |
| b-value 1 | 0 s/mm² |
| b-value 2 | 2000 s/mm² |
| Averages 1 | 1 |
| Averages 2 | 1 |
| Dynamic Field Correction | Off |
| Invert Gray Scale | Off |
| Diff. Weighted Images | On |
| Trace Weighted Images | On |
| Tensor | On |
| FA Maps | On |
| ADC Maps | On |
| Exponential ADC Maps | Off |
| ADC Noise Threshold | 40 |
| Calculated Image | Off |

#### Sequence - Part 1

| | |
|---|---|
| Sequence Name | epse |
| Dimension | 2D |
| Excitation | Standard |
| Gradient Mode | Performance |
| Bandwidth | 1786 Hz/Px |
| Echo Spacing | 0.66 ms |
| Free Echo Spacing | Off |

#### Sequence - Part 1

| | |
|---|---|
| EPI Factor | 140 |

#### Sequence - Part 2

| | |
|---|---|
| Introduction | On |
| RF Spoiling | Off |
| Phase Correction | Internal |

#### Sequence - Special

| | |
|---|---|
| Excite pulse duration | 4480 us |
| Refocus pulse duration | 8320 us |
| Min. prep scans | 0 |
| Min. prep scans SB | 0 |
| Delay before PC scans | 0 us |
| Single-band images | On |
| MB LeakBlock kernel | On |
| MB dual kernel | Off |
| MB RF phase scramble | Off |
| Opt. MB RF pulse BW | Off |
| Time-shifted MB RF | Off |
| SENSE1 coil combine | On |
| Invert RO/PE polarity | On |
| PF omits higher k-space | Off |
| Disable freq. update | Off |
| Suppress 16-bit DICOM | Off |
| Force equal slice timing | Off |
| Online multi-band recon. | Online |
| FFT scale factor | 1.00 |
| Physio recording | DICOM |

#### Sequence - Assistant

| | |
|---|---|
| SAR Assistant | Off |

**\\PHYSICS\XA30 RESEARCH PROT\PARCELLATION\BRAINGATE2 08-26-2024\dMRI_dir97_AP**

TA: 5:34 min Coil Selection: Auto Voxel Size: 1.5×1.5×1.5 mm³ Acc:: None Rel. SNR: 1.00

### Properties

| | |
|---|---|
| Start measurement without further preparation | On |
| Wait for User to Start | Off |
| Start measurements | Single Measurement |
| Prio Recon | Off |
| Auto Open Inline Display | Off |
| Auto Close Inline Display | Off |
| Load Images to MR View&GO | On |
| Auto Store Images | On |
| Load Images to Stamp Segments | Off |
| Load Images to Graphic Segments | Off |
| Graphic segment | Default |
| Inline Movie | Off |

### Routine

| | |
|---|---|
| Slice Group | 1 |
| Slices | 92 |
| Distance Factor | 0 % |
| Position | R1.5 A2.6 H5.8 mm |
| Orientation | T > S-0.4 > C-0.1 |
| Phase Encoding Dir. | A >> P |
| Phase Oversampling | 0 % |
| FoV Read | 210 mm |
| FoV Phase | 100.0 % |
| Slice Thickness | 1.5 mm |
| TR | 3230.0 ms |
| TE | 89.20 ms |
| Multi-band accel. factor | 4 |
| AutoAlign | Head > Brain |

### Contrast - Common

| | |
|---|---|
| TR | 3230.0 ms |
| TE | 89.20 ms |
| MTC | Off |
| Magn. Preparation | None |
| Flip Angle | 78 deg |
| Refocus flip angle | 160 deg |
| Fat-Water Contrast | Standard |
| Grad. rev. fat suppr. | Enabled |
| Reconstruction | Magnitude |

### Contrast - Dynamic

| | |
|---|---|
| Dynamic Mode | Standard |
| Multiple Series | Off |
| Delay in TR | 0.00 ms |

### Resolution - Common

| | |
|---|---|
| FoV Read | 210 mm |
| FoV Phase | 100.0 % |
| Slice Thickness | 1.5 mm |
| Base Resolution | 140 |
| Phase Resolution | 100 % |
| Interpolation | Off |

### Resolution - Acceleration

| | |
|---|---|
| Acceleration mode | None |
| Advanced Reconstruction | Off |
| Phase Partial Fourier | 6/8 |

### Resolution - Filter

| | |
|---|---|
| Raw Filter | Off |
| Elliptical Filter | Off |
| Distortion Correction | Off |
| Normalize | Off |

### Geometry - Common

| | |
|---|---|
| Slice Group | 1 |
| Slices | 92 |
| Distance Factor | 0 % |
| Position | R1.5 A2.6 H5.8 mm |
| Orientation | T > S-0.4 > C-0.1 |
| Phase Encoding Dir. | A >> P |
| Phase Oversampling | 0 % |
| FoV Read | 210 mm |
| FoV Phase | 100.0 % |
| Slice Thickness | 1.5 mm |
| TR | 3230.0 ms |
| Multi-Slice Mode | Interleaved |
| Series | Interleaved |
| Multi-band accel. factor | 4 |

### Geometry - AutoAlign

| | |
|---|---|
| Slice Group | 1 |
| Position | R1.5 A2.6 H5.8 mm |
| Orientation | T > S-0.4 > C-0.1 |
| Phase Encoding Dir. | A >> P |
| AutoAlign | Head > Brain |
| Initial Position | R1.5 A2.6 H5.8 |
| R | 1.5 mm |
| A | 2.6 mm |
| H | 5.8 mm |
| Initial Orientation | T > S |
| T > S | -0.40 |
| > C | -0.10 |
| Initial Rotation | 0.14 deg |

### Geometry - Navigator

### Geometry - Saturation

| | |
|---|---|
| Special Saturation | None |

### Geometry - Tim Planning Suite

| | |
|---|---|
| Set-n-Go Protocol | Off |
| Table Position | 6 mm |
| Table Position | H |
| Inline Composing | Off |

### System - Miscellaneous

| | |
|---|---|
| Coil Selection | Auto Coil Select |
| MSMA | S - C - T |
| Sagittal | R >> L |
| Coronal | A >> P |
| Transversal | F >> H |
| Coil Combination | Sum of Squares |
| Matrix Optimization | Off |
| Coil Focus | Flat |

### System - Adjustments

| | |
|---|---|
| Adjustment Strategy | Standard |
| B0 Shim | Standard |

### System - Adjustments

| | |
|---|---|
| B1 Shim | TrueForm |
| CoilShim | Off |
| Adjustment Tolerance | Auto |
| Adjust with Body Coil | Off |
| Confirm Frequency | Never |
| Assume Silicone | Off |

### System - Adjust Volume

| | |
|---|---|
| Position | R1.5 A2.6 H5.8 mm |
| Orientation | T > S-0.4 > C-0.1 |
| Rotation | 0.14 deg |
| A >> P | 210 mm |
| R >> L | 210 mm |
| F >> H | 138 mm |
| Reset | Off |

### System - pTx

| | |
|---|---|
| B1 Shim | TrueForm |
| Excitation | Standard |

### System - Tx/Rx

| | |
|---|---|
| Frequency 1H | 123.257916 MHz |
| ? Ref. Amplitude 1H | 0.000 V |
| Reset | Off |
| Image Scaling | 1.000 |

### Physio - Signal

| | |
|---|---|
| 1st Signal/Mode | None |
| TR | 3230.0 ms |
| Multi-band accel. factor | 4 |

### Physio - PACE

| | |
|---|---|
| Resp. Control | Off |
| Multi-band accel. factor | 4 |

### Diff

| | |
|---|---|
| Diffusion Mode | Free |
| Diff. Directions | 97 |
| Diffusion Scheme | Monopolar |
| Diff. Weightings | 2 |
| b-value 1 | 0 s/mm² |
| b-value 2 | 3000 s/mm² |
| Averages 1 | 1 |
| Averages 2 | 1 |
| Dynamic Field Correction | Off |
| Invert Gray Scale | Off |
| Diff. Weighted Images | On |
| Trace Weighted Images | On |
| Tensor | On |
| FA Maps | On |
| ADC Maps | On |
| Exponential ADC Maps | Off |
| ADC Noise Threshold | 40 |
| Calculated Image | Off |

### Sequence - Part 1

| | |
|---|---|
| Sequence Name | epse |
| Dimension | 2D |
| Excitation | Standard |
| Gradient Mode | Performance |
| Bandwidth | 1786 Hz/Px |
| Echo Spacing | 0.66 ms |
| Free Echo Spacing | Off |

### Sequence - Part 1

| | |
|---|---|
| EPI Factor | 140 |

### Sequence - Part 2

| | |
|---|---|
| Introduction | On |
| RF Spoiling | Off |
| Phase Correction | Internal |

### Sequence - Special

| | |
|---|---|
| Excite pulse duration | 4480 us |
| Refocus pulse duration | 8320 us |
| Min. prep scans | 0 |
| Min. prep scans SB | 0 |
| Delay before PC scans | 0 us |
| Single-band images | On |
| MB LeakBlock kernel | On |
| MB dual kernel | Off |
| MB RF phase scramble | Off |
| Opt. MB RF pulse BW | Off |
| Time-shifted MB RF | Off |
| SENSE1 coil combine | On |
| Invert RO/PE polarity | Off |
| PF omits higher k-space | Off |
| Disable freq. update | Off |
| Suppress 16-bit DICOM | Off |
| Force equal slice timing | Off |
| Online multi-band recon. | Online |
| FFT scale factor | 1.00 |
| Physio recording | DICOM |

### Sequence - Assistant

| | |
|---|---|
| SAR Assistant | Off |

**\\PHYSICS\XA30 RESEARCH PROT\PARCELLATION\BRAINGATE2 08-26-2024\dMRI_dir97_PA**

TA: 5:34 min Coil Selection: Auto Voxel Size: 1.5×1.5×1.5 mm³ Acc:: None Rel. SNR: 1.00

### Properties

| | |
|---|---|
| Start measurement without further preparation | On |
| Wait for User to Start | Off |
| Start measurements | Single Measurement |
| Prio Recon | Off |
| Auto Open Inline Display | Off |
| Auto Close Inline Display | Off |
| Load Images to MR View&GO | On |
| Auto Store Images | On |
| Load Images to Stamp Segments | Off |
| Load Images to Graphic Segments | Off |
| Graphic segment | Default |
| Inline Movie | Off |

### Routine

| | |
|---|---|
| Slice Group | 1 |
| Slices | 92 |
| Distance Factor | 0 % |
| Position | R1.5 A2.6 H5.8 mm |
| Orientation | T > S-0.4 > C-0.1 |
| Phase Encoding Dir. | A >> P |
| Phase Oversampling | 0 % |
| FoV Read | 210 mm |
| FoV Phase | 100.0 % |
| Slice Thickness | 1.5 mm |
| TR | 3230.0 ms |
| TE | 89.20 ms |
| Multi-band accel. factor | 4 |
| AutoAlign | Head > Brain |

### Contrast - Common

| | |
|---|---|
| TR | 3230.0 ms |
| TE | 89.20 ms |
| MTC | Off |
| Magn. Preparation | None |
| Flip Angle | 78 deg |
| Refocus flip angle | 160 deg |
| Fat-Water Contrast | Standard |
| Grad. rev. fat suppr. | Enabled |
| Reconstruction | Magnitude |

### Contrast - Dynamic

| | |
|---|---|
| Dynamic Mode | Standard |
| Multiple Series | Off |
| Delay in TR | 0.00 ms |

### Resolution - Common

| | |
|---|---|
| FoV Read | 210 mm |
| FoV Phase | 100.0 % |
| Slice Thickness | 1.5 mm |
| Base Resolution | 140 |
| Phase Resolution | 100 % |
| Interpolation | Off |

### Resolution - Acceleration

| | |
|---|---|
| Acceleration mode | None |
| Advanced Reconstruction | Off |
| Phase Partial Fourier | 6/8 |

### Resolution - Filter

| | |
|---|---|
| Raw Filter | Off |
| Elliptical Filter | Off |
| Distortion Correction | Off |
| Normalize | Off |

### Geometry - Common

| | |
|---|---|
| Slice Group | 1 |
| Slices | 92 |
| Distance Factor | 0 % |
| Position | R1.5 A2.6 H5.8 mm |
| Orientation | T > S-0.4 > C-0.1 |
| Phase Encoding Dir. | A >> P |
| Phase Oversampling | 0 % |
| FoV Read | 210 mm |
| FoV Phase | 100.0 % |
| Slice Thickness | 1.5 mm |
| TR | 3230.0 ms |
| Multi-Slice Mode | Interleaved |
| Series | Interleaved |
| Multi-band accel. factor | 4 |

### Geometry - AutoAlign

| | |
|---|---|
| Slice Group | 1 |
| Position | R1.5 A2.6 H5.8 mm |
| Orientation | T > S-0.4 > C-0.1 |
| Phase Encoding Dir. | A >> P |
| AutoAlign | Head > Brain |
| Initial Position | R1.5 A2.6 H5.8 |
| R | 1.5 mm |
| A | 2.6 mm |
| H | 5.8 mm |
| Initial Orientation | T > S |
| T > S | -0.40 |
| > C | -0.10 |
| Initial Rotation | 0.14 deg |

### Geometry - Navigator

### Geometry - Saturation

| | |
|---|---|
| Special Saturation | None |

### Geometry - Tim Planning Suite

| | |
|---|---|
| Set-n-Go Protocol | Off |
| Table Position | 6 mm |
| Table Position | H |
| Inline Composing | Off |

### System - Miscellaneous

| | |
|---|---|
| Coil Selection | Auto Coil Select |
| MSMA | S - C - T |
| Sagittal | R >> L |
| Coronal | A >> P |
| Transversal | F >> H |
| Coil Combination | Sum of Squares |
| Matrix Optimization | Off |
| Coil Focus | Flat |

### System - Adjustments

| | |
|---|---|
| Adjustment Strategy | Standard |
| B0 Shim | Standard |

### System - Adjustments

| | |
|---|---|
| B1 Shim | TrueForm |
| CoilShim | Off |
| Adjustment Tolerance | Auto |
| Adjust with Body Coil | Off |
| Confirm Frequency | Never |
| Assume Silicone | Off |

### System - Adjust Volume

| | |
|---|---|
| Position | R1.5 A2.6 H5.8 mm |
| Orientation | T > S-0.4 > C-0.1 |
| Rotation | 0.14 deg |
| A >> P | 210 mm |
| R >> L | 210 mm |
| F >> H | 138 mm |
| Reset | Off |

### System - pTx

| | |
|---|---|
| B1 Shim | TrueForm |
| Excitation | Standard |

### System - Tx/Rx

| | |
|---|---|
| Frequency 1H | 123.257916 MHz |
| ? Ref. Amplitude 1H | 0.000 V |
| Reset | Off |
| Image Scaling | 1.000 |

### Physio - Signal

| | |
|---|---|
| 1st Signal/Mode | None |
| TR | 3230.0 ms |
| Multi-band accel. factor | 4 |

### Physio - PACE

| | |
|---|---|
| Resp. Control | Off |
| Multi-band accel. factor | 4 |

### Diff

| | |
|---|---|
| Diffusion Mode | Free |
| Diff. Directions | 97 |
| Diffusion Scheme | Monopolar |
| Diff. Weightings | 2 |
| b-value 1 | 0 s/mm² |
| b-value 2 | 3000 s/mm² |
| Averages 1 | 1 |
| Averages 2 | 1 |
| Dynamic Field Correction | Off |
| Invert Gray Scale | Off |
| Diff. Weighted Images | On |
| Trace Weighted Images | On |
| Tensor | On |
| FA Maps | On |
| ADC Maps | On |
| Exponential ADC Maps | Off |
| ADC Noise Threshold | 40 |
| Calculated Image | Off |

### Sequence - Part 1

| | |
|---|---|
| Sequence Name | epse |
| Dimension | 2D |
| Excitation | Standard |
| Gradient Mode | Performance |
| Bandwidth | 1786 Hz/Px |
| Echo Spacing | 0.66 ms |
| Free Echo Spacing | Off |

### Sequence - Part 1

| | |
|---|---|
| EPI Factor | 140 |

### Sequence - Part 2

| | |
|---|---|
| Introduction | On |
| RF Spoiling | Off |
| Phase Correction | Internal |

### Sequence - Special

| | |
|---|---|
| Excite pulse duration | 4480 us |
| Refocus pulse duration | 8320 us |
| Min. prep scans | 0 |
| Min. prep scans SB | 0 |
| Delay before PC scans | 0 us |
| Single-band images | On |
| MB LeakBlock kernel | On |
| MB dual kernel | Off |
| MB RF phase scramble | Off |
| Opt. MB RF pulse BW | Off |
| Time-shifted MB RF | Off |
| SENSE1 coil combine | On |
| Invert RO/PE polarity | On |
| PF omits higher k-space | Off |
| Disable freq. update | Off |
| Suppress 16-bit DICOM | Off |
| Force equal slice timing | Off |
| Online multi-band recon. | Online |
| FFT scale factor | 1.00 |
| Physio recording | DICOM |

### Sequence - Assistant

| | |
|---|---|
| SAR Assistant | Off |

**\\PHYSICS\XA30 RESEARCH PROT\PARCELLATION\BRAINGATE2 08-26-2024\Post_Sag_T2_3D_Space_Wave**

TA: 1:38 min Coil Selection: Auto Voxel Size: 1.0×1.0×1.0 mm³ Acc:: 6 Rel. SNR: 1.00

### Properties

| | |
|---|---|
| Start measurement without further preparation | On |
| Wait for User to Start | Off |
| Start measurements | Single Measurement |
| Prio Recon | Off |
| Auto Open Inline Display | Off |
| Auto Close Inline Display | Off |
| Load Images to MR View&GO | On |
| Auto Store Images | On |
| Load Images to Stamp Segments | Off |
| Load Images to Graphic Segments | Off |
| Graphic segment | Default |
| Inline Movie | Off |

### Routine

| | |
|---|---|
| Slab Group | 1 |
| Slabs | 1 |
| Position | R1.6 A29.4 F10.9 mm |
| Orientation | S > T-2.8 > C-0.8 |
| Phase Encoding Dir. | A >> P |
| Slices per Slab | 224 |
| Phase Oversampling | 0 % |
| Slice Oversampling | 0.0 % |
| FoV Read | 256 mm |
| FoV Phase | 100.0 % |
| Slice Thickness | 1.00 mm |
| TR | 3200.0 ms |
| TE | 407.00 ms |
| Averages | 1.0 |
| Concatenations | 1 |
| AutoAlign | Head > Basis |

### Contrast - Common

| | |
|---|---|
| TR | 3200.0 ms |
| TE | 407.00 ms |
| MTC | Off |
| Magn. Preparation | None |
| Flip Angle Mode | T2 Var |
| Fat-Water Contrast | Standard |
| Dark Blood | Off |
| Blood Suppression | Off |
| Wrap-up Magn. | Restore |
| Reconstruction | Magnitude |

### Contrast - Dynamic

| | |
|---|---|
| Dynamic Mode | Standard |
| Measurements | 1 |
| Multiple Series | Each Measurement |
| Reordering | Linear |

### Resolution - Common

| | |
|---|---|
| FoV Read | 256 mm |
| FoV Phase | 100.0 % |
| Slice Thickness | 1.00 mm |
| Base Resolution | 256 |
| Phase Resolution | 100 % |
| Slice Resolution | 100 % |
| Interpolation | Off |

### Resolution - Acceleration

| | |
|---|---|
| Acceleration mode | CAIPIRINHA |
| Total Factor | 6 |
| Reference Scans | GRE/Separate |
| Acceleration Factor PE | 3 |
| Reference Lines PE | 24 |
| Acceleration Factor 3D | 2 |
| Reference Lines 3D | 48 |
| Reordering Shift 3D | 0 |
| Phase Partial Fourier | Allowed |
| Slice Partial Fourier | Off |
| Elliptical Scanning | On |

### Resolution - Filter

| | |
|---|---|
| Distortion Correction | 3D |
| Normalize | Prescan |
| Image Filter | Off |

### Geometry - Common

| | |
|---|---|
| Slab Group | 1 |
| Slabs | 1 |
| Position | R1.6 A29.4 F10.9 mm |
| Orientation | S > T-2.8 > C-0.8 |
| Phase Encoding Dir. | A >> P |
| Slices per Slab | 224 |
| Phase Oversampling | 0 % |
| Slice Oversampling | 0.0 % |
| FoV Read | 256 mm |
| FoV Phase | 100.0 % |
| Slice Thickness | 1.00 mm |
| TR | 3200.0 ms |
| Concatenations | 1 |

### Geometry - AutoAlign

| | |
|---|---|
| Slab Group | 1 |
| Position | R1.6 A29.4 F10.9 mm |
| Orientation | S > T-2.8 > C-0.8 |
| Phase Encoding Dir. | A >> P |
| AutoAlign | Head > Basis |
| Initial Position | L0.4 P0.4 H3.5 |
| L | 0.4 mm |
| P | 0.4 mm |
| H | 3.5 mm |
| Initial Orientation | S > T |
| S > T | 0.80 |
| > C | 0.10 |
| Initial Rotation | 6.56 deg |

### Geometry - Navigator

### Geometry - Saturation

| | |
|---|---|
| Special Saturation | None |

### Geometry - Tim Planning Suite

| | |
|---|---|
| Set-n-Go Protocol | Off |
| Table Position | 0 mm |
| Table Position | H |
| Inline Composing | Off |

### System - Miscellaneous

| | |
|---|---|
| Coil Selection | Auto Coil Select |
| MSMA | S - C - T |
| Sagittal | R >> L |
| Coronal | A >> P |
| Transversal | F >> H |
| Coil Combination | Sum of Squares |
| Matrix Optimization | Performance |
| Coil Focus | Flat |

### System - Adjustments

| | |
|---|---|
| Adjustment Strategy | Standard |
| B0 Shim | Tune up |
| B1 Shim | TrueForm |
| CoilShim | Off |
| Adjustment Tolerance | Auto |
| Adjust with Body Coil | Off |
| Confirm Frequency | Never |
| Assume Silicone | Off |

### System - Adjust Volume

| | |
|---|---|
| Position | Isocenter |
| Orientation | Transversal |
| Rotation | 0.00 deg |
| A >> P | 263 mm |
| R >> L | 350 mm |
| F >> H | 350 mm |
| Reset | Off |

### System - pTx

| | |
|---|---|
| B1 Shim | TrueForm |
| Excitation | Non-sel. |

### System - Tx/Rx

| | |
|---|---|
| Frequency 1H | 123.257916 MHz |
| ? Ref. Amplitude 1H | 0.000 V |
| Reset | Off |
| Image Scaling | 1.000 |

### Physio - Signal

| | |
|---|---|
| 1st Signal/Mode | None |
| Trigger Delay | 0 ms |
| TR | 3200.0 ms |
| Concatenations | 1 |

### Physio - Cardiac

| | |
|---|---|
| Fat-Water Contrast | Standard |
| Magn. Preparation | None |
| Dark Blood | Off |
| FoV Read | 256 mm |
| FoV Phase | 100.0 % |
| Phase Resolution | 100 % |
| Dynamic Mode | Standard |

### Physio - PACE

| | |
|---|---|
| Resp. Control | Off |
| Concatenations | 1 |

### Inline - Subtraction

| | |
|---|---|
| Subtract | Off |
| Measurements | 1 |
| StdDev | Off |
| Save Original Images | On |

### Inline - Cardiac

| | |
|---|---|
| Magn. Preparation | None |
| Save Original Images | On |
| TE | 407.00 ms |
| TR | 3200.0 ms |

### Inline - MIP

| | |
|---|---|
| MIP Sag | Off |
| MIP Cor | Off |
| MIP Tra | Off |
| MIP Time | Off |
| Radial MIP | Off |
| Save Original Images | On |
| MPR Sag | Off |
| MPR Cor | Off |
| MPR Tra | Off |

### Inline - Composing

| | |
|---|---|
| Inline Composing | Off |

### Sequence - Part 1

| | |
|---|---|
| Sequence Name | WIP_spcR |
| Dimension | 3D |
| Excitation | Non-sel. |
| RF Pulse Type | Normal |
| Gradient Mode | Fast |
| Flow Compensation | None |
| Reordering | Linear |
| Bandwidth | 651 Hz/Px |
| Echo Spacing | 3.84 ms |
| Turbo Factor | 282 |
| Echo Train Duration | 952 ms |

### Sequence - Part 2

| | |
|---|---|
| Introduction | On |

### Sequence - Special

| | |
|---|---|
| Play Blip | Yes |
| Wave | Both |
| Coil Mixing | On |
| ROFilt Pre | 10 perc |
| ROFilt Post | 0 perc |
| LINFilt Pre | 0 perc |
| LINFilt Post | 0 perc |
| PARFilt Pre | 10 perc |
| PARFilt Post | 10 perc |
| Reg. Beta | 0.20 |
| Reg. Lambda | 0.20 |
| MAX Slew wave | 160.0 mT/m/ms |
| MAX Ampl wave | 20.0 mT/m |
| NumOfCycles Wave | 5.0 |

### Sequence - Assistant

| | |
|---|---|
| SAR Assistant | Off |
| Allowed Delay | 0 s |

**\\PHYSICS\XA30 RESEARCH PROT\PARCELLATION\BRAINGATE2 08-26-2024\Post Sag_T1_3D_Mprage**

TA: 5:21 min Coil Selection: Auto Voxel Size: 1.0×1.0×0.9 mm³ Acc:: 2 Rel. SNR: 1.00

### Properties

| | |
|---|---|
| Start measurement without further preparation | On |
| Wait for User to Start | Off |
| Start measurements | Single Measurement |
| Prio Recon | Off |
| Auto Open Inline Display | Off |
| Auto Close Inline Display | Off |
| Load Images to MR View&GO | On |
| Auto Store Images | On |
| Load Images to Stamp Segments | Off |
| Load Images to Graphic Segments | Off |
| Graphic segment | Default |
| Inline Movie | Off |

### Routine

| | |
|---|---|
| Slab Group | 1 |
| Slabs | 1 |
| Distance Factor | 50 % |
| Position | R1.6 A29.4 F10.9 mm |
| Orientation | S > T-2.8 > C-0.8 |
| Phase Encoding Dir. | A >> P |
| Slices per Slab | 192 |
| Phase Oversampling | 0 % |
| Slice Oversampling | 16.7 % |
| FoV Read | 260 mm |
| FoV Phase | 100.0 % |
| Slice Thickness | 0.9 mm |
| TR | 2300.0 ms |
| TE | 2.32 ms |
| Averages | 1 |
| Concatenations | 1 |
| AutoAlign | Head > Basis |

### Contrast - Common

| | |
|---|---|
| TR | 2300.0 ms |
| TE | 2.32 ms |
| Magn. Preparation | Non-sel. IR |
| TI | 900 ms |
| Flip Angle | 8 deg |
| Fat-Water Contrast | Standard |
| Dark Blood | Off |
| Reconstruction | Magnitude |

### Contrast - Dynamic

| | |
|---|---|
| Dynamic Mode | Standard |
| Measurements | 1 |
| Multiple Series | Each Measurement |
| Reordering | Linear |

### Resolution - Common

| | |
|---|---|
| FoV Read | 260 mm |
| FoV Phase | 100.0 % |
| Slice Thickness | 0.9 mm |
| Base Resolution | 256 |
| Phase Resolution | 100 % |
| Slice Resolution | 100 % |
| Interpolation | Off |

### Resolution - Acceleration

| | |
|---|---|
| Acceleration mode | GRAPPA |
| Reference Scans | Integrated |
| Acceleration Factor PE | 2 |
| Reference Lines PE | 24 |
| Acceleration Factor 3D | 1 |
| Phase Partial Fourier | Off |
| Slice Partial Fourier | Off |
| Asymmetric Echo | Allowed |
| Elliptical Scanning | Off |

### Resolution - Filter

| | |
|---|---|
| Raw Filter | Off |
| Elliptical Filter | Off |
| Distortion Correction | 2D |
| Normalize | Prescan |
| Image Filter | Off |

### Geometry - Common

| | |
|---|---|
| Slab Group | 1 |
| Slabs | 1 |
| Distance Factor | 50 % |
| Position | R1.6 A29.4 F10.9 mm |
| Orientation | S > T-2.8 > C-0.8 |
| Phase Encoding Dir. | A >> P |
| Slices per Slab | 192 |
| Phase Oversampling | 0 % |
| Slice Oversampling | 16.7 % |
| FoV Read | 260 mm |
| FoV Phase | 100.0 % |
| Slice Thickness | 0.9 mm |
| TR | 2300.0 ms |
| Multi-Slice Mode | Single Shot |
| Series | Ascending |
| Concatenations | 1 |

### Geometry - AutoAlign

| | |
|---|---|
| Slab Group | 1 |
| Position | R1.6 A29.4 F10.9 mm |
| Orientation | S > T-2.8 > C-0.8 |
| Phase Encoding Dir. | A >> P |
| AutoAlign | Head > Basis |
| Initial Position | L0.4 P0.4 H3.5 |
| L | 0.4 mm |
| P | 0.4 mm |
| H | 3.5 mm |
| Initial Orientation | S > T |
| S > T | 0.80 |
| > C | 0.10 |
| Initial Rotation | 6.56 deg |

### Geometry - Navigator

### Geometry - Tim Planning Suite

| | |
|---|---|
| Set-n-Go Protocol | Off |
| Table Position | 0 mm |
| Table Position | H |
| Inline Composing | Off |

### System - Miscellaneous

| | |
|---|---|
| Coil Selection | ACS All but spine |
| MSMA | S - C - T |
| Sagittal | R >> L |
| Coronal | A >> P |
| Transversal | F >> H |
| Coil Combination | Adaptive Combine |
| Matrix Optimization | Off |
| Coil Focus | Flat |

### System - Adjustments

| | |
|---|---|
| Adjustment Strategy | Standard |
| B0 Shim | Tune up |
| B1 Shim | TrueForm |
| CoilShim | Off |
| Adjustment Tolerance | Auto |
| Adjust with Body Coil | Off |
| Confirm Frequency | Never |
| Assume Silicone | Off |

### System - Adjust Volume

| | |
|---|---|
| Position | Isocenter |
| Orientation | Transversal |
| Rotation | 0.00 deg |
| A >> P | 263 mm |
| R >> L | 350 mm |
| F >> H | 350 mm |
| Reset | Off |

### System - pTx

| | |
|---|---|
| B1 Shim | TrueForm |
| Excitation | Non-sel. |

### System - Tx/Rx

| | |
|---|---|
| Frequency 1H | 123.257916 MHz |
| ? Ref. Amplitude 1H | 0.000 V |
| Reset | Off |
| Image Scaling | 1.000 |

### Physio - Signal

| | |
|---|---|
| 1st Signal/Mode | None |
| TR | 2300.0 ms |
| Concatenations | 1 |

### Physio - Cardiac

| | |
|---|---|
| Fat-Water Contrast | Standard |
| Magn. Preparation | Non-sel. IR |
| TI | 900 ms |
| Dark Blood | Off |
| FoV Read | 260 mm |
| FoV Phase | 100.0 % |
| Phase Resolution | 100 % |
| Dynamic Mode | Standard |

### Physio - PACE

| | |
|---|---|
| Resp. Control | Off |
| Concatenations | 1 |

### Inline - Subtraction

| | |
|---|---|
| Subtract | Off |
| Measurements | 1 |
| StdDev | Off |
| Save Original Images | On |

### Inline - Cardiac

| | |
|---|---|
| Magn. Preparation | Non-sel. IR |
| Save Original Images | On |
| TE | 2.32 ms |
| TR | 2300.0 ms |

### Inline - MIP

| | |
|---|---|
| MIP Sag | Off |
| MIP Cor | Off |
| MIP Tra | Off |
| MIP Time | Off |
| Radial MIP | Off |
| Save Original Images | On |
| MPR Sag | Off |
| MPR Cor | Off |
| MPR Tra | Off |

### Inline - Composing

| | |
|---|---|
| Inline Composing | Off |

### Inline - MapIt

| | |
|---|---|
| MapIt | None |
| Flip Angle | 8 deg |
| Measurements | 1 |
| TE | 2.32 ms |
| TR | 2300.0 ms |
| Save Original Images | On |

### Sequence - Part 1

| | |
|---|---|
| Sequence Name | tfl |
| Dimension | 3D |
| Excitation | Non-sel. |
| RF Pulse Type | Normal |
| Gradient Mode | Normal |
| Flow Compensation | None |
| Reordering | Linear |
| Bandwidth | 200 Hz/Px |
| Echo Spacing | 6.86 ms |
| Asymmetric Echo | Allowed |
| Turbo Factor | 224 |

### Sequence - Part 2

| | |
|---|---|
| Introduction | On |
| RF Spoiling | On |
| Incr. Gradient Spoiling | Off |

### Sequence - Assistant

| | |
|---|---|
| SAR Assistant | Off |
